# Gaps in lipid management after diabetes diagnosis and associated cardiovascular outcomes in a cohort of US adults

**DOI:** 10.64898/2026.05.24.26354000

**Authors:** Alexander M. Heilman, Theodore Warsavage, Wenhui Liu, Peter W.F. Wilson, Lawrence S. Phillips, Jane E.B. Reusch, Sridharan Raghavan

**Author notes:** <u>Address for Correspondence:</u> Sridharan Raghavan, VA Eastern Colorado Health Care System 1700 North Wheeling Street, Aurora, CO 80045.

## Abstract

**Importance:** Despite the benefits of statin therapy in individuals with diabetes, fewer than 70% of adults with diabetes meet contemporary guidelines for statin therapy and reducing low-density lipoprotein cholesterol (LDL) to <100 mg/dL. Evidence describing delays in statin initiation after diabetes diagnosis and associated clinical outcomes may motivate process of care interventions to improve guideline recommended care in individuals newly diagnosed with type 2 diabetes mellitus (T2D).

**Objective:** To examine the timing of statin initiation and achievement of LDL <100 mg/dL after diabetes diagnosis, and to determine the association of early LDL reduction among statin initiators with incident atherosclerotic cardiovascular disease (ASCVD).

**Design:** Retrospective observational cohort study using data from 2005-2021

**Setting:** Veterans Affairs Health Care System (VA)

**Participants:** Individuals with newly diagnosed T2D

**Exposure:** Primary exposure was ASCVD risk based on ACC/AHA Pooled Cohort Equations; secondary exposure was LDL <100 mg/dL in the first year after T2D diagnosis among statin initiators

**Main Outcomes and Measures:** Co-primary outcomes were initiation of statin therapy and achievement of LDL <100 mg/dL within 5 years of diabetes diagnosis; incident 5-year ASCVD was a secondary outcome.

**Results:** Among 100,406 individuals with newly diagnosed T2D, 59,615 were prescribed statin therapy within five years (59.4%), and 44,783 (57.5%) of those with LDL above goal achieved LDL <100 mg/dL within 5 years. Relative to those at low (<7.5%) 10-year ASCVD risk, individuals at intermediate (7.5-20%) and high (>20%) risk were more likely to be initiated on a statin (intermediate: Hazard Ratio [HR] 1.14 [95% CI 1.11, 1.17]; high: HR 1.16 [95% CI 1.13, 1.19]) and to achieve LDL <100 mg/dL (intermediate: HR 1.23 [95% CI 1.19, 1.26]; high: HR 1.34 [95% CI 1.30, 1.38]). Among those prescribed a statin within one year of diabetes diagnosis, achieving LDL <100 mg/dL in the first year after diabetes diagnosis was associated with lower risk of 5-year incident ASCVD (HR 0.84 [95% CI 0.77, 0.92]).

**Conclusions and Relevance:** Gaps in guideline-directed primary prevention of ASCVD arise early following initial diabetes diagnosis. Guideline recommended early LDL lowering among statin initiators was associated with improved clinical outcomes.

## Introduction

Type 2 diabetes mellitus (T2D) affects approximately 10% of United States (US) adults, and individuals with T2D are at increased risk of atherosclerotic cardiovascular disease (ASCVD) and cardiovascular mortality.^1–5^ Accordingly, professional society guidelines for T2D management recommend aggressive ASCVD risk factor modification, specifically lipid-lowering with statin therapy and blood pressure control, in addition to early optimization of glycemic control.^6–9^ However, numerous studies have found substantial gaps in achieving the multiple evidence-based treatment goals recommended by professional society guidelines for individuals with diabetes, including low rates of statin prescribing and LDL reduction to goal levels.^10–13^

This poor adherence to guideline recommendations reflects a chronic disease management challenge for individuals with newly diagnosed T2D: initiating multiple lines of treatment (statin therapy, glucose-lowering therapy, possibly blood pressure treatment) and surveillance for treatment response and incident complications. There is extensive evidence describing clinical inertia and outcomes associated with early glycemic treatment in those with newly diagnosed T2D.^14–21^ Analogous studies describing the timing of statin initiation and LDL lowering after diabetes diagnosis and associated clinical outcomes may inform prioritization of the multiple interventions recommended for individuals newly diagnosed with T2D.

To address this evidence gap, we examined the timing of statin initiation and lipid lowering in individuals for whom a new T2D diagnosis represented a new indication for cholesterol management with statins in the Veterans Affairs Health Care System (VA), the largest integrated health system in the US. In addition, to determine if early lipid lowering warrants prioritization among the multiple recommended interventions after T2D diagnosis, we assessed the association of early LDL reduction in statin-treated individuals with 5-year ASCVD incidence in individuals with newly diagnosed T2D.

## Methods

### Study Population

This study included individuals who received routine primary care within the VA and were newly diagnosed with T2D between 2005 and 2016. Follow-up continued through the end of 2022 to ensure that all individuals were eligible for at least 5 years of follow-up. T2D status was defined as use of an outpatient diabetes mellitus medication combined with either ≥2 uses of *International Classification of Diseases*, Ninth Revision, Clinical Modification (ICD-9) diagnosis codes 250.xx or ≥1 use of 250.xx in conjunction with a primary care physician visit. The first occurrence satisfying these criteria was deemed the date of diagnosis. To be included, individuals must have had at least one VA primary care visit in the two years prior to the first use of a diabetes diagnosis code without any use of a diabetes-related diagnosis code or a diabetes medication prescription, and they had to have filled at least one non-diabetes medication prescription in the VA in the two years prior to the date of diabetes diagnosis. We did not include individuals who met laboratory criteria for diabetes without an accompanying diagnosis code and medication use as the laboratory criteria used in the VA varied over the period of the study, and the absence of a diagnosis code for diabetes in VA records may reflect deferral of diabetes management to a non-VA provider. As we were interested in individuals for whom diabetes diagnosis represented a new indication for statin therapy, we excluded those with prevalent ASCVD, defined as a history of coronary artery disease, myocardial infarction, ischemic stroke, heart failure, peripheral artery disease (PAD), or who had previously undergone percutaneous coronary intervention (PCI) or coronary artery bypass grafting (CABG), and those with a statin prescription at the time of diabetes diagnosis. In analyses for which LDL <100 mg/dL was the outcome, we further restricted our analytic sample to those with LDL >100 mg/dL at baseline. The Colorado Multiple Institutional Review Board and the Research & Development Committee of the VA Eastern Colorado Healthcare System provided human subjects oversight and approval of this work.

### Outcomes

The two co-primary outcomes for this study were a) initiation of a statin medication and b) achievement of LDL <100 mg/dL, both within five years of initial diabetes diagnosis. Five-year incident ASCVD in those started on statin therapy within one year of diabetes diagnosis was a secondary outcome. All statins approved for use in the US were identified according to their generic or trade name. A target LDL of 100 mg/dL was chosen as this was the guideline specified LDL-C goal during the observation analysis period.^9,22^ To satisfy the requirement of achieving LDL <100 mg/dL, individuals must have had two consecutive LDL measurements <100 mg/dL, with the first occurrence being deemed the date that target LDL was achieved. ASCVD diagnoses were identified by ICD-9/10 codes within the electronic health record (EHR) (**Supplement A**).

### Predictor Variables and Subgroup Analyses

For analyses of statin initiation and LDL reduction, the primary predictor variable was predicted ASCVD risk. We estimated 10-year ASCVD risk based on the American College of Cardiology/American Heart Association Pooled Cohorts Equations (PCE),^9^ categorized as low-(<7.5%), intermediate- (7.5-20%), and high-risk (>20%). We examined temporal trends in statin initiation and target LDL reduction using the year of diabetes diagnosis as a predictor variable, with 2005 as the reference group. Finally, subgroup analyses were performed to evaluate for differences in statin initiation and achieving LDL <100 mg/dL by sex and by self-reported race/ethnicity. For analyses of incident ASCVD events (secondary outcome), the LDL value nearest to one-year and occurring between 6 and 18 months after diabetes diagnosis was the primary predictor variable (categorized as <100 versus ≥100 mg/dL or as <70, 70-100, and >100 mg/dL).

### Covariates

All multivariable models were adjusted for age, sex, race/ethnicity, body mass index, baseline laboratory parameters (total cholesterol (TC), LDL, high-density lipoprotein cholesterol (HDL), hemoglobin A1c (HbA1c)), comorbidities associated with ASCVD (hypertension, atrial fibrillation, obstructive sleep apnea, liver disease, chronic kidney disease, and smoking status), and medications commonly used for primary prevention of ASCVD (aspirin, antihypertensives, and non-statin lipid-lowering medications). Psychiatric diagnoses (anxiety, depression, substance abuse, and schizophrenia) and aging-specific variables (dementia, Alzheimer’s disease, arthritis, fatigue, and walking difficulty) were included to account for potential reasons why individuals may not have been prescribed or been adherent to statin treatment.

### Statistical Analysis

We plotted unadjusted cumulative incidence curves to describe time to the co-primary and secondary outcomes, using a log-rank test to compare individuals stratified by ASCVD risk, race/ethnicity, sex, and LDL achieved in the first year after T2D diagnosis. We used multivariable Cox proportional hazards models to examine associations of ASCVD risk, year of diabetes diagnosis, race/ethnicity, and sex with the co-primary outcomes, censoring individuals who died prior to statin initiation or achieving LDL <100 mg/dL. The covariates listed above were included in the multivariable analyses with one exception. When ASCVD risk was the predictor/independent variable of interest, the variables included in the PCE (age, sex, race/ethnicity, TC, HDL, hypertension, smoking status, and use of antihypertensives) were omitted as covariates. End of follow-up was considered as whichever occurred last of the following dates: last HbA1c or LDL measurement, first statin prescription, or death (all restricted to within six years of initial diagnosis). Those who received an ASCVD diagnosis prior to occurrence of the primary outcome were censored.

To evaluate the relationship between early LDL reduction in those receiving statin therapy with cardiovascular outcomes, we used multivariable Cox proportional hazards models to examine the association of one-year LDL values (defined as above) with the occurrence of an incident ASCVD diagnosis within five years. This analysis was limited to individuals who were newly prescribed a statin within one year of T2D diagnosis. We performed this secondary analysis in two ways. First, for consistency with the primary analyses above, we examined LDL at one year after diabetes diagnosis categorized as <100 mg/dL versus ≥100 mg/dL. Second, in consideration of contemporary diabetes treatment guidelines,^23^ we repeated the secondary analysis using a three-category specification of one-year LDL (<70, 70-100, and >100 mg/dL). For the latter analysis, we also stratified by risk categories using the PCE (10-year risk <7.5%, 7.5-20%, and >20%). To account for intraindividual variability in LDL, we performed sensitivity analyses excluding those with baseline LDL below 100 mg/dL and below 120 mg/dL. Those who experienced an incident ASCVD event prior to initiation of statin therapy or within one year of initial diabetes diagnosis were censored.

A *p*-value of 0.05 was used to denote significance. All analyses were performed using *R* version 4.0.3 (R Foundation for Statistical Computing, Vienna, Austria). Statistical code for all analyses is available upon request.

## Results

### Baseline Characteristics

There were 375,186 individuals who met criteria for newly diagnosed T2D and started an oral T2D medication between 2005 and 2016. Of these, 23,649 were excluded due to missing demographic or covariate data, 143,246 were excluded due to prevalent ASCVD, and 107,885 were excluded due to use of statin therapy at time of T2D diagnosis. This resulted in 100,406 individuals with the diabetes diagnosis representing a new indication for statin initiation. Of these, 77,893 had baseline LDL >100 mg/dL. The baseline characteristics of included participants are shown in **Table 1**.

**Table 1.** Study participant characteristics.

|  |  |
| --- | --- |
| **Baseline characteristics** |  |
| Total population | 100,406 |
| Age, median (IQR) | 58.5 (50.6-65.0) |
| Male sex, n (%) | 94,634 (94.3) |
| Race/ethnicity, n (%) |  |
| *White* | 58,743 (58.5) |
| *Black* | 24,989 (24.9) |
| *Hispanic* | 7,884 (7.9) |
| *Other* | 8,790 (8.8) |
| Body mass index, median (IQR) | 32.6 (28.8-37.1) |
| Total cholesterol, median (IQR) | 182 (157-208) |
| *High-density lipoprotein* | 32 (37-45) |
| *Low-density lipoprotein* | 105 (84-128) |
| **Comorbidities, n (%)** |  |
| Hypertension | 71,470 (71.2) |
| Hyperlipidemia | 38,344 (38.2) |
| Atrial fibrillation | 2,409 (2.4) |
| Obstructive sleep apnea | 17,731 (17.7) |
| Liver disease | 12,936 (12.9) |
| Chronic kidney disease | 11,582 (11.5) |
| Smoking status |  |
| *Current* | 32,836 (32.7) |
| *Former* | 41,687 (41.5) |
| **Medications, n (%)** |  |
| Aspirin | 6,058 (6.0) |
| Antihypertensive agent | 59,531 (59.3) |
| Other lipid-lowering medication | 6,734 (6.7) |
| **Psychiatric diagnoses, n (%)** |  |
| Anxiety/depression | 40,366 (40.2) |
| Schizophrenia | 3,917 (3.9) |
| Substance use | 12,011 (12.0) |
| **Aging-specific variables, n (%)** |  |
| Dementia | 1,509 (1.5) |
| Alzheimer's disease | 185 (0.2) |
| Arthritis | 40,083 (39.9) |
| Fatigue | 2,921 (2.9) |
| Walking difficulty | 3,067 (3.1) |
| **Predicted 10-year ASCVD risk, n (%)** |  |
| Low-risk (<7.5%) | 15,420 (15.4) |
| Intermediate-risk (7.5-20%) | 31,896 (31.8) |
| High-risk (>20%) | 53,090 (52.9) |

### Co-primary Outcomes

A total of 59,615 individuals (59.4%) were started on statin therapy within 5 years of initial diabetes diagnosis. ASCVD risk based on the PCE was associated with statin initiation (**Figure 1**). In adjusted analyses, individuals at intermediate risk (10-year ASCVD risk of 7.5-20%) and high risk (10-year ASCVD risk of >20%) had higher likelihood of statin initiation within 5 years of diabetes diagnosis than those at low risk (Hazard Ratio [HR] 1.14 (95% Confidence Interval [CI] 1.11-1.17) for intermediate-risk and HR 1.16 [95% CI 1.13-1.19] for high-risk, both p<0.001). Only 19,606 individuals (19.5%) received a statin prescription within 4 weeks of initial T2D treatment, and the association of ASCVD risk with 5-year statin initiation was similar when these early initiators were excluded (**eFigure 1**). Over 35% of individuals were not initiated on statin therapy within 5 years of initial diabetes diagnosis irrespective of estimated ASCVD risk (**Table 2**).

**Figure 1.**
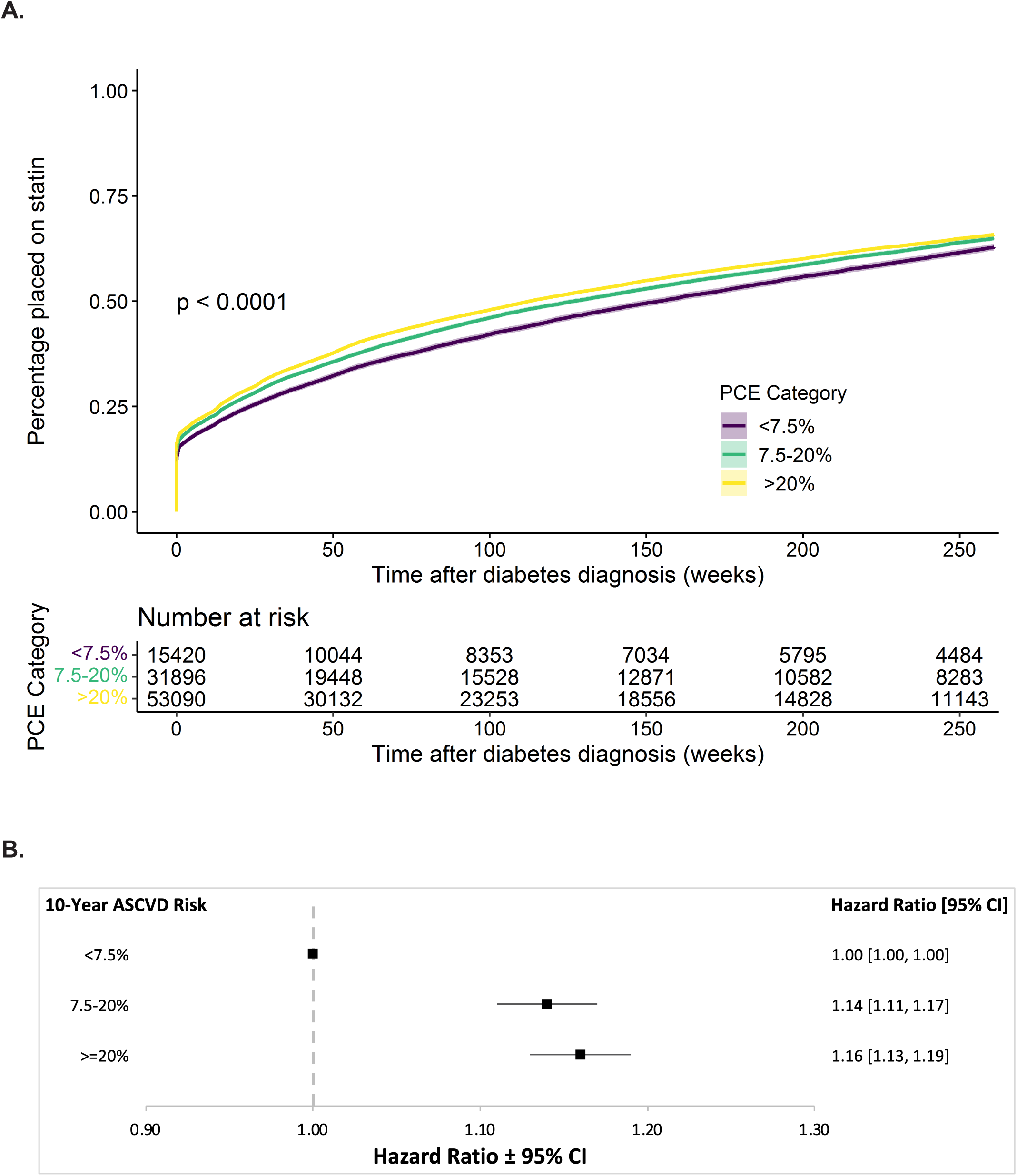
(A) Crude cumulative incidence of initiation of statin therapy following initial diabetes diagnosis, stratified by 10-year ASCVD risk as predicted by the ACC/AHA Pooled Cohorts Equation (PCE, low risk <7.5%, intermediate risk 7.5-20%, and high risk >20%). (B) Forest plot of HR for statin initiation within 5 years for intermediate- and high-risk individuals compared to low-risk individuals after covariate adjustment.

**Table 2.**
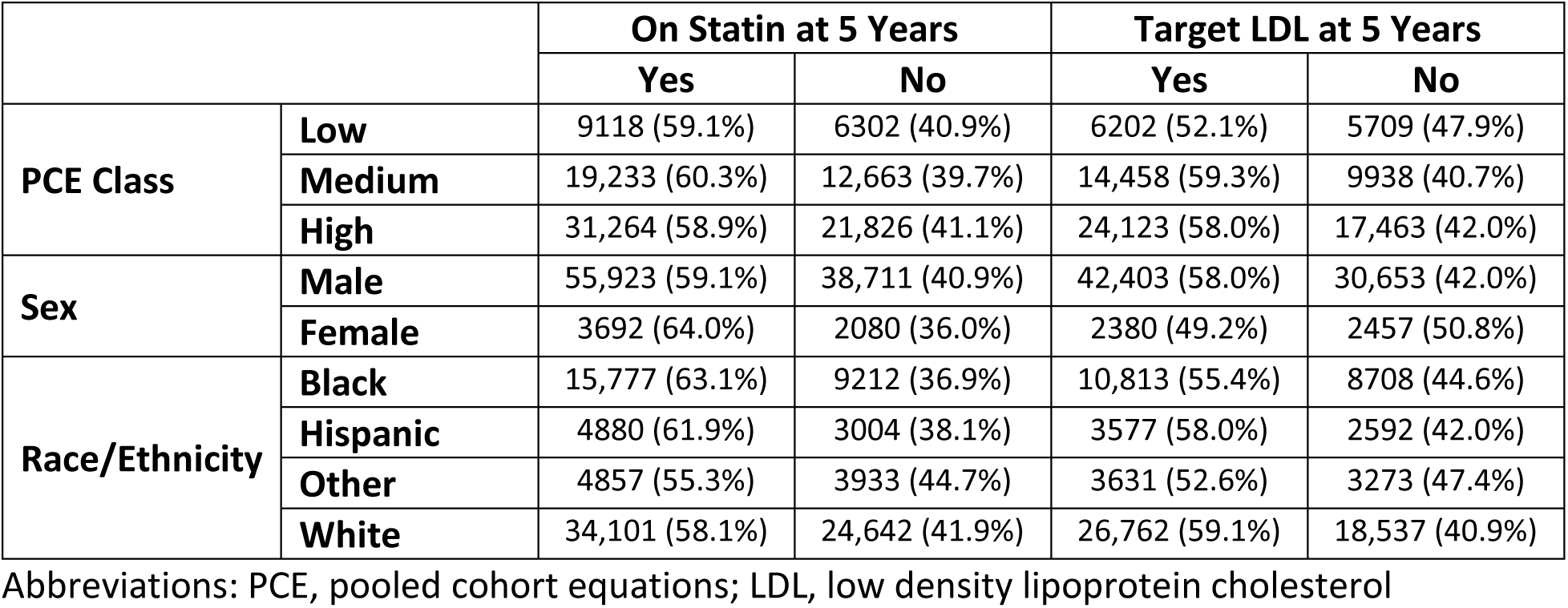
Crude rates of statin initiation and achievement of LDL <100 mg/dL.

A total of 44,783 individuals (57.5%) who had baseline LDL values >100 mg/dL achieved LDL <100 mg/dL within 5 years of initial T2D diagnosis. Individuals who were deemed at intermediate or high ASCVD risk were more likely to achieve target LDL than those at low risk (**Figure 2**; adjusted HR 1.23 [95% CI 1.19-1.26] in intermediate-risk and 1.34 [95% CI 1.30-1.38] in high-risk individuals relative to those at low risk; both p<0.001). The proportional reduction in LDL from baseline at end of follow-up increased as ASCVD risk increased (low: 2.9% [SD 0.37], intermediate: 8.5% [SD 0.37], high: 13.5% [SD 0.36]; **eTable 1**). As with statin initiation, over 35% of individuals with LDL >100 mg/dL at the time of T2D diagnosis did not achieve LDL levels <100 mg/dL within 5 years of initial diabetes diagnosis irrespective of ASCVD risk category (**Table 2**). We found a similar trend for achievement of LDL <70 mg/dL in those with baseline values above this (N=97,767), with only 21,069 (21.6%) achieving this goal within 5 years of initial T2D diagnosis **(eFigure 2).**

**Figure 2.**
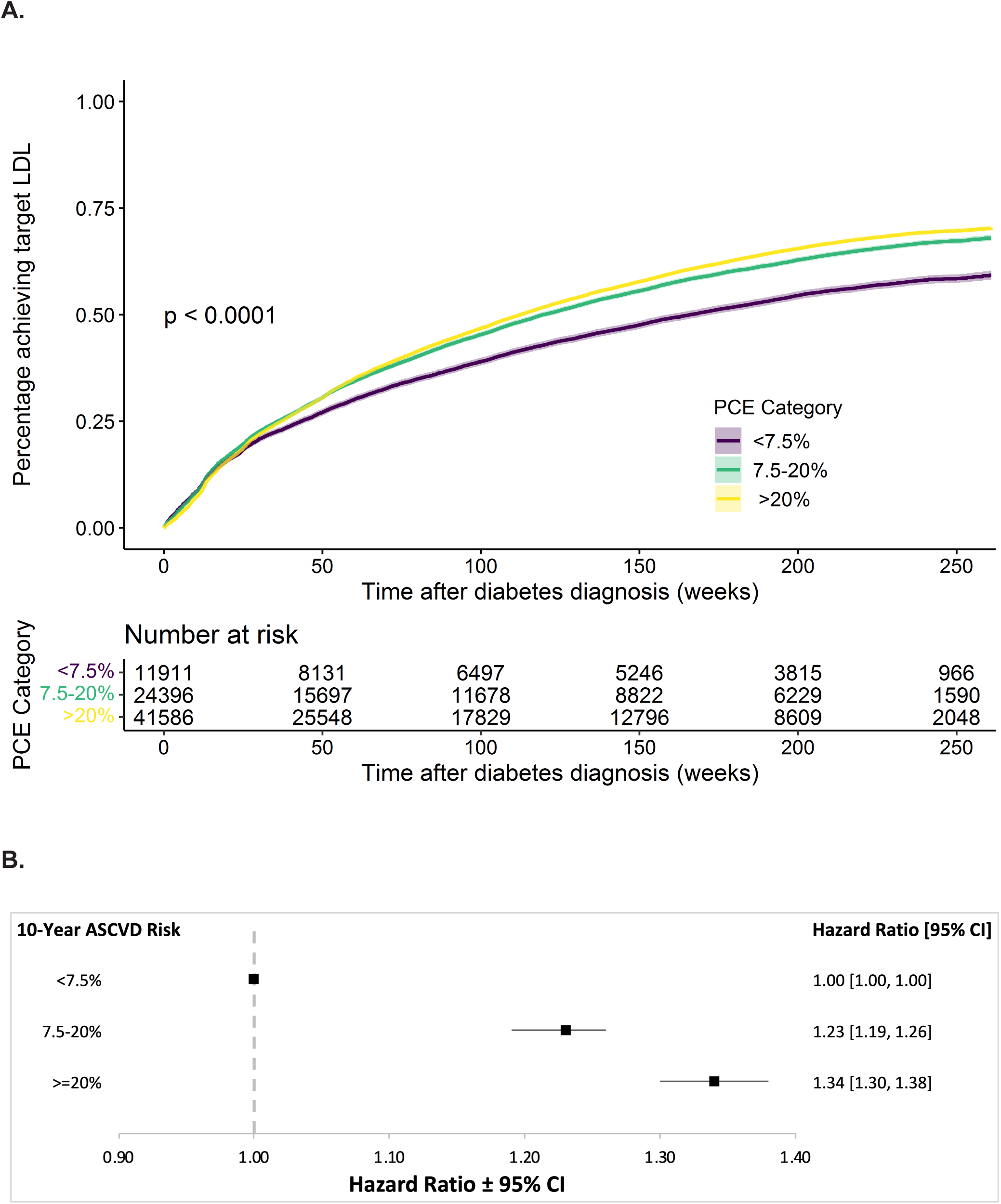
(A) Crude cumulative incidence of time to target LDL reduction following initial diabetes diagnosis, stratified by 10-year ASCVD risk as predicted by the ACC/AHA Pooled Cohorts Equation (PCE, low risk <7.5%, intermediate risk 7.5-20%, and high risk >20%). (B) Forest plot of HR for target LDL reduction within 5 years for intermediate- and high-risk individuals compared to low-risk individuals after covariate adjustment.

Compared to those diagnosed with T2D in 2005, statin initiation and target LDL reduction within 5 years of diabetes diagnosis did not vary substantially across consecutive annual cohorts of newly diagnosed individuals with T2D (**eTable 2)**. After multivariable adjustment, we observed significant year-to-year variation in 5-year statin initiation and target LDL reduction associated with year of T2D diagnosis but without a consistent trend for improvement in either outcome (**eFigure 3)**. We observed similar rates of statin initiation irrespective of sex or race (**Table 2, eFigure 4-5).** Despite similar rates of 5-year statin initiation, women were less likely than men to achieve target LDL levels within 5 years of T2D diagnosis (HR 0.83 [95% CI 0.80-0.87], p<0.001), and Black individuals were less likely than white individuals to achieve target LDL reduction within 5 years of T2D diagnosis (HR 0.91 [95% CI 0.88-0.94], p<0.001).

### Secondary Outcome

A total of 37,008 (36.9%) individuals were started on a statin within one year of initial diabetes diagnosis. We excluded 361 (1.0%) individuals who were diagnosed with ASCVD in the first year after T2D diagnosis prior to initiation of statin therapy and 6185 (16.7%) who did not have an LDL measurement between 6 and 18 months after T2D diagnosis. The baseline characteristics of those who did and did not have one-year LDL measurements were similar (**eTable 3**). The distribution of predicted cardiovascular risk and one-year LDL categories is shown in **eTable 4**. Among those prescribed a statin within one year of diabetes diagnosis, achieving LDL <100 mg/dL in the first year after diabetes diagnosis was associated with lower risk of 5-year incident ASCVD (HR 0.84 [95% CI 0.77, 0.92], **Figure 3, eTable 5)**. In analysis including three target LDL categories (<70, 70-100, and >100 mg/dL), individuals with one-year LDL <70 mg/dL experienced fewer incident ASCVD event within 5 years of initial diabetes diagnosis than those with one-year LDL >100 mg/dL (adjusted HR 0.73, 95% CI 0.64-0.83 for LDL <70 mg/dL, p<0.001; HR 0.90, 95% CI 0.82-0.99 for LDL 70-100 mg/dL, p=0.032; **Figure 3, eTable 5**). In those predicted to be at low ASCVD risk, neither one-year LDL <70 mg/dL (HR 0.59, 95% CI 0.32-1.09, p=0.092) nor 70-100 mg/dL (HR 0.75, 95% CI 0.49-1.13, p=0.163) were associated with five-year ASCVD compared to those with one-year LDL >100 mg/dL. In individuals predicted to be at intermediate or high risk, those with one-year LDL <70 mg/dL experienced fewer 5-year ASCVD events when compared to those with one-year LDL >100 mg/dL (HR 0.75, 95% CI 0.59-0.95, p=0.019 for LDL <70 mg/dL, intermediate risk; HR 0.71, 95% CI 0.61-0.84, p<0.001 for LDL <70 mg/dL, high risk; **Figure 3, eTable 5**). Among individuals predicted to be at intermediate or high risk, LDL 70-100 mg/dL at one year was not significantly associated with 5-year ASCVD events compared to those with LDL >100 mg/dL (HR 0.83, 95% CI 0.69-1.00, p=0.055 for intermediate risk; HR 0.93, 95% CI 0.83-1.05, p=0.237 for high risk; **Figure 3, eTable 5**). Sensitivity analyses excluding individuals with baseline LDL values less than 100 mg/dL (n=6559) and less than 120 mg/dL (n=13,901) had similar findings (**eTable 6**).

**Figure 3.**
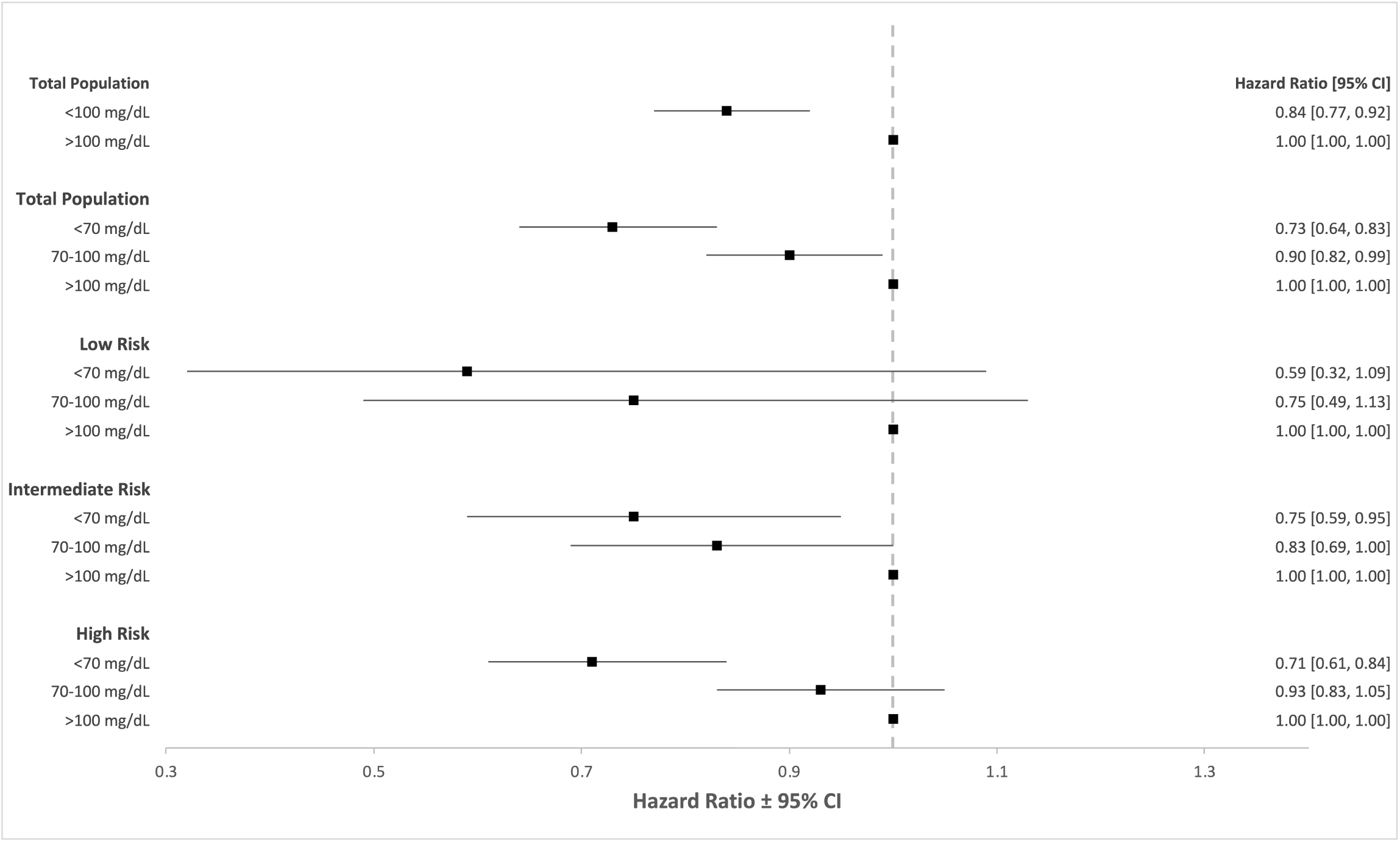
Forest plot of covariate-adjusted HR for incident ASCVD diagnosis within 5 years, stratified by one-year LDL values and ASCVD risk class.

## Discussion

We found that gaps in primary prevention of ASCVD through statin initiation and LDL reduction arise early after diabetes diagnosis within the VA. Guidelines recommend that all individuals aged 40-75 diagnosed with diabetes be placed on at minimum moderate-intensity statin therapy with a goal LDL <100 mg/dL.^6,24^ Despite this, nearly 40% of individuals in our study had not been started on guideline-directed statin therapy and nearly 40% of those with baseline LDL values >100 mg/dL failed to achieve target LDL reduction within five years of initial T2D diagnosis. Serial cross-sectional studies utilizing data from the National Health and Nutrition Examination Survey (NHANES) have shown similar proportions of individuals with diabetes not on statin therapy with LDL levels above target.^10,11,25–27^ Our study suggests that these gaps arise early and are at least partially due to a lack of statin initiation and intensification rather than inadequate maintenance of recommended therapy. This widespread failure to initiate guideline-directed treatment early after diabetes diagnosis may lead to persistent undertreatment of cardiovascular risk and may contribute to adverse ASCVD outcomes.

Stratification of our population into categories based on ASCVD risk showed that individuals who were predicted to be at higher risk were prioritized to receive statin therapy and were more likely to achieve LDL values <100 mg/dL within 5 years of diagnosis. This prioritization demonstrates that there is some appropriate focus on risk-based treatment; however, differences between groups are small and not clearly due to systematic differences in management approaches. Over 35% of individuals in all risk groups were not treated with statin therapy and failed to achieve target LDL reduction. These results are similar to that seen in NHANES, which showed that statin use in individuals with diabetes ranged from 43% to 55% from low-risk to high-risk individuals (those with prior ASCVD)^25^. While the analyses are not identical or directly comparable, the results are similar in that the gradient of lipid-lowering across risk categories in those with diabetes is modest.

The results of this study suggest clinical inertia in diabetes care that may have worsened over time. We found that approximately 20% of individuals are started on statin therapy within the first month of diabetes diagnosis, irrespective of ASCVD risk. Of those not started around the time of initial diagnosis, the rate of new starts is gradual and roughly linear over the subsequent years. In addition, despite the strengthening of guidelines in recent years, there were not significant improvements in either statin prescribing or target LDL reduction over time. There are several explanations for this temporal trend. First, this may reflect worsening competing demands during medical visits over time. The number of electronic reminders for clinical care issued to VA primary care providers is substantial and has grown with time^28^, potentially contributing to the adverse temporal trend in lipid management for Veterans with diabetes. Second, VA quality measures may have inadvertently prioritized certain aspects of diabetes care. For example, for several years during our study period, the VA primary care quality metrics emphasized achieving a goal HbA1c <9%, inherently prioritizing glycemic control over lipid lowering for Veterans with diabetes. The temporal trends observed for statin initiation and achieving target LDL levels mirror prior work in Veterans showing that diabetes treatment intensification for glycemic control occurred later after initial diabetes diagnosis and at progressively higher HbA1c levels over time from 2005 to 2013^14^. Third, the characteristics of Veterans with diabetes changed over the study period with increasing proportions of individuals newly diagnosed with diabetes of Black race, female sex, with comorbid sleep apnea or with comorbid psychiatric diagnoses **(eTable 7)**. It is possible that these changing patient characteristics could impact treatment preferences related to statin use and intensity of lipid lowering.

Prior studies have shown a benefit of early glycemic and hypertension control in the reduction of adverse cardiovascular events in those newly diagnosed with diabetes^19–21^. Similar studies evaluating the benefit of early lipid control after diabetes diagnosis for reducing ASCVD risk are lacking despite evidence of elevated ASCVD risk in individuals with diabetes^1,29–37^ and that statin therapy can reduce this risk^38–42^. Our study found that of individuals started on statin therapy within one year of diabetes diagnosis, early achievement of LDL <100 mg/dL was associated with 16% reduction in incident ASCVD diagnoses within 5 years of initial diabetes diagnosis compared to those who did not achieve early LDL reduction. Moreover, we observed that lower thresholds of LDL reduction were necessary for significant associations with ASCVD risk reduction as ASCVD risk increased. This is supportive of the 2023 update to the ADA Standards of Care in Diabetes, which recommends a graded approach to lipid lowering in individuals with diabetes based on predicted risk of ASCVD^23^. In sum, the results suggest potential ASCVD risk reduction associated with early LDL lowering with statin treatment – observational evidence that LDL reduction should be prioritized among the initial steps in care for individuals newly diagnosed with diabetes.

### Limitations

First, the veteran patient population differs from the general adult diabetes population in the US, limiting generalizability. Our study was primarily composed of white male veterans 40 years and older with T2D, and prior work has shown that veterans have more diabetes complications, greater ASCVD risk, and were more likely to be treated with lipid lowering medications to goal LDL than the general US population.^43^ Second, the specific dose of statin prescribed was not reliably available in our data, so we were unable to determine if individuals were placed on moderate versus high intensity statin therapy based on ASCVD risk and/or initial LDL level. Third, our study did not evaluate medication adherence, which is one mechanism by which goal LDL levels might not be achieved despite initiating statin therapy. Our aim was focused on describing guideline directed treatment and associated outcomes early after diabetes diagnosis; future studies will explore mechanisms and possible interventions for care improvement. Fourth, we cannot determine from the EHR data used for this study if there were specific reasons for avoiding statin use in certain patients. As with any treatment decision, statin initiation requires a discussion with patients and is subject to shared decision making. For many patients, initiation of a daily medication is a considerable step, especially when they are otherwise at low cardiovascular risk and they are concurrently being initiated on one or more medications for diabetes treatment. That said, contraindications and patient preference are unlikely to explain the nearly 40% gap in statin use in our sample. Finally, the study design precludes causal inference regarding one-year LDL levels and ASCVD outcomes. That is, achievement of low LDL levels at one year may not be causally associated with ASCVD risk reduction if LDL lowering is correlated with other ASCVD risk-reducing characteristics.

### Conclusion

Gaps in statin initiation and LDL reduction arise early following diabetes diagnosis within the VA. Gaps are present regardless of predicted ASCVD risk, sex, and race/ethnicity. Early LDL lowering among statin initiators was associated with a lower incidence of early ASCVD diagnoses following diabetes diagnosis. Delay in statin initiation and LDL reduction places diabetes patients at increased risk for potentially preventable adverse cardiovascular outcomes.

## Author Contributions

Study design was conceived by AMH, SR, and JEBR. Data collection and organization were performed by AMH, SR, TW, and WGL. Analyses were performed by AMH. All authors participated in interpreting results, manuscript writing and critical revision; all authors approve of the manuscript submission in its current form. SR is the guarantor of the manuscript, including study design, analyses, and drafting, submission, and publication of the manuscript.

## Supporting information

Supplemental Results

## Data Availability

Statistical code for all analyses is available upon request. A deidentified, anonymized limited data set derived from the datasets used for the analysis can be made available upon reasonable request from researchers with necessary human subjects research oversight and in accordance with VA data sharing policies.

