## Supplemental Methods for "Gaps in lipid management after diabetes diagnosis and associated cardiovascular outcomes in a cohort of US adults"

| ICD9 Code | ICD9 Proc Code | ICD10 Code | ICD10 Proc Code | CPT Code |
|---|---|---|---|---|
| 346.60 | 00.55 | E08.51 | 0210083 | 0001T |
| 346.61 | 00.63 | E08.52 | 0210088 | 0002T |
| 346.62 | 00.66 | E09.51 | 0210089 | 0075T |
| 346.63 | 17.55 | E09.52 | 021008C | 0076T |
| 362.34 | 17.56 | E10.51 | 021008F | 0078T |
| 410.0 | 36.03 | E10.52 | 021008W | 0079T |
| 410.00 | 36.04 | E11.51 | 0210093 | 0234T |
| 410.01 | 36.06 | E11.52 | 0210098 | 0235T |
| 410.02 | 36.07 | E13.51 | 0210099 | 0236T |
| 410.1 | 36.09 | E13.52 | 021009C | 0237T |
| 410.10 | 36.10 | G43.601 | 021009F | 0238T |
| 410.11 | 36.11 | G43.609 | 021009W | 33510 |
| 410.12 | 36.12 | G43.611 | 02100A3 | 33511 |
| 410.2 | 36.13 | G43.619 | 02100A8 | 33512 |
| 410.20 | 36.14 | G45.0 | 02100A9 | 33513 |
| 410.21 | 36.15 | G45.1 | 02100AC | 33514 |
| 410.22 | 36.16 | G45.2 | 02100AF | 33516 |
| 410.3 | 36.17 | G45.3 | 02100AW | 33517 |
| 410.30 | 36.19 | G45.8 | 02100J3 | 33518 |
| 410.31 | 36.2 | G45.9 | 02100J8 | 33519 |
| 410.32 | 38.03 | G46.0 | 02100J9 | 33520 |
| 410.4 | 38.04 | G46.1 | 02100JC | 33521 |
| 410.40 | 38.06 | G46.2 | 02100JF | 33522 |
| 410.41 | 38.08 | G46.3 | 02100JW | 33523 |
| 410.42 | 38.12 | G46.4 | 02100K3 | 33533 |
| 410.5 | 38.13 | G46.5 | 02100K8 | 33534 |
| 410.50 | 38.14 | G46.6 | 02100K9 | 33535 |
| 410.51 | 38.16 | G46.7 | 02100KC | 33536 |
| 410.52 | 38.18 | G46.8 | 02100KF | 33545 |
| 410.6 | 38.44 | H34.00 | 02100KW | 33572 |
| 410.60 | 38.48 | H34.01 | 02100Z3 | 33877 |
| 410.61 | 39.22 | H34.02 | 02100Z8 | 34001 |
| 410.62 | 39.24 | H34.03 | 02100Z9 | 34051 |
| 410.7 | 39.25 | I20.0 | 02100ZC | 34101 |
| 410.70 | 39.26 | I20.1 | 02100ZF | 34111 |
| 410.71 | 39.29 | I20.8 | 0210344 | 34151 |
| 410.72 | 39.50 | I20.9 | 02103D4 | 34201 |
| 410.8 | 39.71 | I21.01 | 0210444 | 34203 |
| 410.80 | 39.90 | I21.02 | 0210483 | 34800 |
| 410.81 | | I21.09 | 0210488 | 34802 |
| 410.82 | | I21.11 | 0210489 | 34803 |
| 410.9 | | I21.19 | 021048C | 34804 |

| | | | |
|---|---|---|---|
| 410.90 | I21.21 | 021048F | 34805 |
| 410.91 | I21.29 | 021048W | 34812 |
| 410.92 | I21.3 | 0210493 | 34820 |
| 411.0 | I21.4 | 0210498 | 34825 |
| 411.1 | I21.9 | 0210499 | 34830 |
| 411.8 | I21.A1 | 021049C | 34831 |
| 411.81 | I21.A9 | 021049F | 34832 |
| 411.89 | I22.0 | 021049W | 34833 |
| 412. | I22.1 | 02104A3 | 34834 |
| 413.0 | I22.2 | 02104A8 | 35001 |
| 413.1 | I22.8 | 02104A9 | 35005 |
| 413.9 | I22.9 | 02104AC | 35011 |
| 414.0 | I23.0 | 02104AF | 35021 |
| 414.00 | I23.1 | 02104AW | 35045 |
| 414.01 | I23.2 | 02104D4 | 35081 |
| 414.02 | I23.3 | 02104J3 | 35082 |
| 414.03 | I23.4 | 02104J8 | 35091 |
| 414.04 | I23.5 | 02104J9 | 35092 |
| 414.05 | I23.6 | 02104JC | 35102 |
| 414.06 | I23.7 | 02104JF | 35103 |
| 414.07 | I23.8 | 02104JW | 35301 |
| 414.10 | I24.0 | 02104K3 | 35302 |
| 414.11 | I24.1 | 02104K8 | 35303 |
| 414.12 | I24.8 | 02104K9 | 35304 |
| 414.19 | I24.9 | 02104KC | 35305 |
| 414.2 | I25.10 | 02104KF | 35306 |
| 414.3 | I25.110 | 02104KW | 35311 |
| 414.4 | I25.111 | 02104Z3 | 35321 |
| 414.8 | I25.118 | 02104Z8 | 35331 |
| 414.9 | I25.119 | 02104Z9 | 35341 |
| 429.71 | I25.2 | 02104ZC | 35351 |
| 429.79 | I25.3 | 02104ZF | 35355 |
| 430. | I25.41 | 0211083 | 35361 |
| 431. | I25.42 | 0211088 | 35363 |
| 432.0 | I25.5 | 0211089 | 35371 |
| 432.1 | I25.6 | 021108C | 35372 |
| 432.9 | I25.700 | 021108F | 35381 |
| 433.0 | I25.701 | 021108W | 35390 |
| 433.00 | I25.708 | 0211093 | 35450 |
| 433.01 | I25.709 | 0211098 | 35452 |
| 433.1 | I25.710 | 0211099 | 35454 |
| 433.10 | I25.711 | 021109C | 35456 |
| 433.11 | I25.718 | 021109F | 35458 |

| | | | |
|---|---|---|---|
| 433.2 | I25.719 | 021109W | 35459 |
| 433.20 | I25.720 | 02110A3 | 35470 |
| 433.21 | I25.721 | 02110A8 | 35471 |
| 433.3 | I25.728 | 02110A9 | 35472 |
| 433.30 | I25.729 | 02110AC | 35473 |
| 433.31 | I25.730 | 02110AF | 35474 |
| 433.8 | I25.731 | 02110AW | 35475 |
| 433.80 | I25.738 | 02110J3 | 35480 |
| 433.81 | I25.739 | 02110J8 | 35481 |
| 433.9 | I25.750 | 02110J9 | 35482 |
| 433.90 | I25.751 | 02110JC | 35483 |
| 433.91 | I25.758 | 02110JF | 35484 |
| 434.0 | I25.759 | 02110JW | 35485 |
| 434.00 | I25.760 | 02110K3 | 35490 |
| 434.01 | I25.761 | 02110K8 | 35491 |
| 434.1 | I25.768 | 02110K9 | 35492 |
| 434.10 | I25.769 | 02110KC | 35493 |
| 434.11 | I25.790 | 02110KF | 35494 |
| 434.9 | I25.791 | 02110KW | 35495 |
| 434.90 | I25.798 | 02110Z3 | 35501 |
| 434.91 | I25.799 | 02110Z8 | 35507 |
| 435.0 | I25.810 | 02110Z9 | 35508 |
| 435.1 | I25.811 | 02110ZC | 35509 |
| 435.2 | I25.812 | 02110ZF | 35510 |
| 435.3 | I25.82 | 0211344 | 35511 |
| 435.8 | I25.83 | 02113D4 | 35512 |
| 435.9 | I25.84 | 0211444 | 35515 |
| 436. | I25.89 | 0211483 | 35516 |
| 437.0 | I25.9 | 0211488 | 35518 |
| 437.1 | I51.0 | 0211489 | 35521 |
| 437.2 | I51.7 | 021148C | 35522 |
| 437.3 | I60.00 | 021148F | 35523 |
| 437.4 | I60.01 | 021148W | 35525 |
| 437.5 | I60.02 | 0211493 | 35526 |
| 437.6 | I60.10 | 0211498 | 35531 |
| 437.7 | I60.11 | 0211499 | 35533 |
| 437.8 | I60.12 | 021149C | 35535 |
| 437.9 | I60.20 | 021149F | 35536 |
| 438 | I60.21 | 021149W | 35537 |
| 438.0 | I60.22 | 02114A3 | 35538 |
| 438.10 | I60.30 | 02114A8 | 35539 |
| 438.11 | I60.31 | 02114A9 | 35540 |
| 438.12 | I60.32 | 02114AC | 35541 |

| | | | |
|---|---|---|---|
| 438.13 | I60.4 | 02114AF | 35546 |
| 438.14 | I60.50 | 02114AW | 35548 |
| 438.19 | I60.51 | 02114D4 | 35549 |
| 438.20 | I60.52 | 02114J3 | 35551 |
| 438.21 | I60.6 | 02114J8 | 35556 |
| 438.22 | I60.7 | 02114J9 | 35558 |
| 438.30 | I60.8 | 02114JC | 35560 |
| 438.31 | I60.9 | 02114JF | 35563 |
| 438.32 | I61.0 | 02114JW | 35565 |
| 438.40 | I61.1 | 02114K3 | 35566 |
| 438.41 | I61.2 | 02114K8 | 35570 |
| 438.42 | I61.3 | 02114K9 | 35571 |
| 438.50 | I61.4 | 02114KC | 35582 |
| 438.51 | I61.5 | 02114KF | 35583 |
| 438.52 | I61.6 | 02114KW | 35585 |
| 438.53 | I61.8 | 02114Z3 | 35587 |
| 438.6 | I61.9 | 02114Z8 | 35601 |
| 438.7 | I62.00 | 02114Z9 | 35612 |
| 438.81 | I62.01 | 02114ZC | 35616 |
| 438.82 | I62.02 | 02114ZF | 35621 |
| 438.83 | I62.03 | 0212083 | 35623 |
| 438.84 | I62.1 | 0212088 | 35626 |
| 438.85 | I62.9 | 0212089 | 35631 |
| 438.89 | I63.00 | 021208C | 35632 |
| 438.9 | I63.011 | 021208F | 35633 |
| 440.0 | I63.012 | 021208W | 35634 |
| 440.1 | I63.013 | 0212093 | 35636 |
| 440.2 | I63.019 | 0212098 | 35637 |
| 440.20 | I63.02 | 0212099 | 35638 |
| 440.21 | I63.031 | 021209C | 35641 |
| 440.22 | I63.032 | 021209F | 35642 |
| 440.23 | I63.033 | 021209W | 35645 |
| 440.24 | I63.039 | 02120A3 | 35646 |
| 440.29 | I63.09 | 02120A8 | 35647 |
| 440.30 | I63.10 | 02120A9 | 35650 |
| 440.31 | I63.111 | 02120AC | 35651 |
| 440.32 | I63.112 | 02120AF | 35654 |
| 440.4 | I63.113 | 02120AW | 35656 |
| 440.8 | I63.119 | 02120J3 | 35661 |
| 440.9 | I63.12 | 02120J8 | 35663 |
| 441.3 | I63.131 | 02120J9 | 35665 |
| 441.4 | I63.132 | 02120JC | 35666 |
| 441.5 | I63.133 | 02120JF | 35671 |

| | | | |
|---|---|---|---|
| 441.6 | I63.139 | 02120JW | 35700 |
| 441.7 | I63.19 | 02120K3 | 35879 |
| 441.9 | I63.20 | 02120K8 | 35881 |
| 443.81 | I63.211 | 02120K9 | 35883 |
| 443.9 | I63.212 | 02120KC | 35884 |
| 444.0 | I63.213 | 02120KF | 35901 |
| 444.01 | I63.219 | 02120KW | 35903 |
| 444.09 | I63.22 | 02120Z3 | 35907 |
| 444.21 | I63.231 | 02120Z8 | 36140 |
| 444.22 | I63.232 | 02120Z9 | 36245 |
| 444.81 | I63.233 | 02120ZC | 36246 |
| 444.89 | I63.239 | 02120ZF | 36247 |
| 445.01 | I63.29 | 0212344 | 36248 |
| 445.02 | I63.30 | 02123D4 | 37184 |
| 674.00 | I63.311 | 0212444 | 37185 |
| 674.01 | I63.312 | 0212483 | 37186 |
| 674.02 | I63.313 | 0212488 | 37195 |
| 674.03 | I63.319 | 0212489 | 37201 |
| 674.04 | I63.321 | 021248C | 37205 |
| 996.03 | I63.322 | 021248F | 37206 |
| 997.02 | I63.323 | 021248W | 37207 |
| V12.54 | I63.329 | 0212493 | 37208 |
| V45.81 | I63.331 | 0212498 | 37211 |
| V45.82 | I63.332 | 0212499 | 37215 |
| | I63.333 | 021249C | 37216 |
| | I63.339 | 021249F | 37217 |
| | I63.341 | 021249W | 37218 |
| | I63.342 | 02124A3 | 37220 |
| | I63.343 | 02124A8 | 37221 |
| | I63.349 | 02124A9 | 37222 |
| | I63.39 | 02124AC | 37223 |
| | I63.40 | 02124AF | 37224 |
| | I63.411 | 02124AW | 37225 |
| | I63.412 | 02124D4 | 37226 |
| | I63.413 | 02124J3 | 37227 |
| | I63.419 | 02124J8 | 37228 |
| | I63.421 | 02124J9 | 37229 |
| | I63.422 | 02124JC | 37230 |
| | I63.423 | 02124JF | 37231 |
| | I63.429 | 02124JW | 37232 |
| | I63.431 | 02124K3 | 37233 |
| | I63.432 | 02124K8 | 37234 |
| | I63.433 | 02124K9 | 37235 |

| | | |
|---|---|---|
| I63.439 | 02124KC | 37236 |
| I63.441 | 02124KF | 37237 |
| I63.442 | 02124KW | 37246 |
| I63.443 | 02124Z3 | 37247 |
| I63.449 | 02124Z8 | 4110F |
| I63.49 | 02124Z9 | 75966 |
| I63.50 | 02124ZC | 75968 |
| I63.511 | 02124ZF | 75992 |
| I63.512 | 0213083 | 75993 |
| I63.513 | 0213088 | 75994 |
| I63.519 | 0213089 | 75995 |
| I63.521 | 021308C | 75996 |
| I63.522 | 021308F | 92920 |
| I63.523 | 021308W | 92921 |
| I63.529 | 0213093 | 92924 |
| I63.531 | 0213098 | 92925 |
| I63.532 | 0213099 | 92928 |
| I63.533 | 021309C | 92929 |
| I63.539 | 021309F | 92933 |
| I63.541 | 021309W | 92934 |
| I63.542 | 02130A3 | 92937 |
| I63.543 | 02130A8 | 92938 |
| I63.549 | 02130A9 | 92941 |
| I63.59 | 02130AC | 92943 |
| I63.6 | 02130AF | 92944 |
| I63.81 | 02130AW | 92973 |
| I63.89 | 02130J3 | 92974 |
| I63.9 | 02130J8 | 92975 |
| I65.01 | 02130J9 | 92977 |
| I65.02 | 02130JC | 92980 |
| I65.03 | 02130JF | 92981 |
| I65.09 | 02130JW | 92982 |
| I65.1 | 02130K3 | 92984 |
| I65.21 | 02130K8 | 92995 |
| I65.22 | 02130K9 | 92996 |
| I65.23 | 02130KC | 93540 |
| I65.29 | 02130KF | 93564 |
| I65.8 | 02130KW | 93570 |
| I65.9 | 02130Z3 | C9600 |
| I66.01 | 02130Z8 | C9601 |
| I66.02 | 02130Z9 | C9602 |
| I66.03 | 02130ZC | C9603 |
| I66.09 | 02130ZF | C9604 |

| | | |
|---|---|---|
| I66.11 | 0213344 | C9605 |
| I66.12 | 02133D4 | C9606 |
| I66.13 | 0213444 | C9607 |
| I66.19 | 0213483 | C9608 |
| I66.21 | 0213488 | G0290 |
| I66.22 | 0213489 | G0291 |
| I66.23 | 021348C | S2205 |
| I66.29 | 021348F | S2206 |
| I66.3 | 021348W | S2207 |
| I66.8 | 0213493 | S2208 |
| I66.9 | 0213498 | S2209 |
| I67.0 | 0213499 | S2211 |
| I67.1 | 021349C | |
| I67.2 | 021349F | |
| I67.3 | 021349W | |
| I67.4 | 02134A3 | |
| I67.5 | 02134A8 | |
| I67.6 | 02134A9 | |
| I67.7 | 02134AC | |
| I67.81 | 02134AF | |
| I67.82 | 02134AW | |
| I67.83 | 02134D4 | |
| I67.841 | 02134J3 | |
| I67.848 | 02134J8 | |
| I67.850 | 02134J9 | |
| I67.858 | 02134JC | |
| I67.89 | 02134JF | |
| I67.9 | 02134JW | |
| I68.0 | 02134K3 | |
| I68.2 | 02134K8 | |
| I68.8 | 02134K9 | |
| I69.00 | 02134KC | |
| I69.01 | 02134KF | |
| I69.010 | 02134KW | |
| I69.011 | 02134Z3 | |
| I69.012 | 02134Z8 | |
| I69.013 | 02134Z9 | |
| I69.014 | 02134ZC | |
| I69.015 | 02134ZF | |
| I69.018 | 0270346 | |
| I69.019 | 027034Z | |
| I69.020 | 0270356 | |
| I69.021 | 027035Z | |

| | |
|---|---|
| I69.022 | 0270366 |
| I69.023 | 027036Z |
| I69.028 | 0270376 |
| I69.031 | 027037Z |
| I69.032 | 02703D6 |
| I69.033 | 02703DZ |
| I69.034 | 02703E6 |
| I69.039 | 02703EZ |
| I69.041 | 02703F6 |
| I69.042 | 02703FZ |
| I69.043 | 02703G6 |
| I69.044 | 02703GZ |
| I69.049 | 02703T6 |
| I69.051 | 02703TZ |
| I69.052 | 02703Z6 |
| I69.053 | 02703ZZ |
| I69.054 | 0271346 |
| I69.059 | 027134Z |
| I69.061 | 0271356 |
| I69.062 | 027135Z |
| I69.063 | 0271366 |
| I69.064 | 027136Z |
| I69.065 | 0271376 |
| I69.069 | 027137Z |
| I69.090 | 02713D6 |
| I69.091 | 02713DZ |
| I69.092 | 02713E6 |
| I69.093 | 02713EZ |
| I69.098 | 02713F6 |
| I69.10 | 02713FZ |
| I69.11 | 02713G6 |
| I69.110 | 02713GZ |
| I69.111 | 02713T6 |
| I69.112 | 02713TZ |
| I69.113 | 02713Z6 |
| I69.114 | 02713ZZ |
| I69.115 | 0272346 |
| I69.118 | 027234Z |
| I69.119 | 0272356 |
| I69.120 | 027235Z |
| I69.121 | 0272366 |
| I69.122 | 027236Z |
| I69.123 | 0272376 |

| | |
|---|---|
| I69.128 | 027237Z |
| I69.131 | 02723D6 |
| I69.132 | 02723DZ |
| I69.133 | 02723E6 |
| I69.134 | 02723EZ |
| I69.139 | 02723F6 |
| I69.141 | 02723FZ |
| I69.142 | 02723G6 |
| I69.143 | 02723GZ |
| I69.144 | 02723T6 |
| I69.149 | 02723TZ |
| I69.151 | 02723Z6 |
| I69.152 | 02723ZZ |
| I69.153 | 0273346 |
| I69.154 | 027334Z |
| I69.159 | 0273356 |
| I69.161 | 027335Z |
| I69.162 | 0273366 |
| I69.163 | 027336Z |
| I69.164 | 0273376 |
| I69.165 | 027337Z |
| I69.169 | 02733D6 |
| I69.190 | 02733DZ |
| I69.191 | 02733E6 |
| I69.192 | 02733EZ |
| I69.193 | 02733F6 |
| I69.198 | 02733FZ |
| I69.20 | 02733G6 |
| I69.21 | 02733GZ |
| I69.210 | 02733T6 |
| I69.211 | 02733TZ |
| I69.212 | 02733Z6 |
| I69.213 | 02733ZZ |
| I69.214 | 02C03Z6 |
| I69.215 | 02C03ZZ |
| I69.218 | 02C13Z6 |
| I69.219 | 02C13ZZ |
| I69.220 | 02C23Z6 |
| I69.221 | 02C23ZZ |
| I69.222 | 02C33Z6 |
| I69.223 | 02C33ZZ |
| I69.228 | 0312090 |
| I69.231 | 0312091 |

| | |
|---|---|
| I69.232 | 0312092 |
| I69.233 | 0312093 |
| I69.234 | 0312094 |
| I69.239 | 0312095 |
| I69.241 | 0312096 |
| I69.242 | 0312097 |
| I69.243 | 0312098 |
| I69.244 | 0312099 |
| I69.249 | 031209B |
| I69.251 | 031209C |
| I69.252 | 031209J |
| I69.253 | 031209K |
| I69.254 | 03120A0 |
| I69.259 | 03120A1 |
| I69.261 | 03120A2 |
| I69.262 | 03120A3 |
| I69.263 | 03120A4 |
| I69.264 | 03120A5 |
| I69.265 | 03120A6 |
| I69.269 | 03120A7 |
| I69.290 | 03120A8 |
| I69.291 | 03120A9 |
| I69.292 | 03120AB |
| I69.293 | 03120AC |
| I69.298 | 03120AJ |
| I69.30 | 03120AK |
| I69.31 | 03120J0 |
| I69.310 | 03120J1 |
| I69.311 | 03120J2 |
| I69.312 | 03120J3 |
| I69.313 | 03120J4 |
| I69.314 | 03120J5 |
| I69.315 | 03120J6 |
| I69.318 | 03120J7 |
| I69.319 | 03120J8 |
| I69.320 | 03120J9 |
| I69.321 | 03120JB |
| I69.322 | 03120JC |
| I69.323 | 03120JJ |
| I69.328 | 03120JK |
| I69.331 | 03120K0 |
| I69.332 | 03120K1 |
| I69.333 | 03120K2 |

| | |
|---|---|
| I69.334 | 03120K3 |
| I69.339 | 03120K4 |
| I69.341 | 03120K5 |
| I69.342 | 03120K6 |
| I69.343 | 03120K7 |
| I69.344 | 03120K8 |
| I69.349 | 03120K9 |
| I69.351 | 03120KB |
| I69.352 | 03120KC |
| I69.353 | 03120KJ |
| I69.354 | 03120KK |
| I69.359 | 03120Z0 |
| I69.361 | 03120Z1 |
| I69.362 | 03120Z2 |
| I69.363 | 03120Z3 |
| I69.364 | 03120Z4 |
| I69.365 | 03120Z5 |
| I69.369 | 03120Z6 |
| I69.390 | 03120Z7 |
| I69.391 | 03120Z8 |
| I69.392 | 03120Z9 |
| I69.393 | 03120ZB |
| I69.398 | 03120ZC |
| I69.80 | 03120ZJ |
| I69.81 | 03120ZK |
| I69.810 | 0313090 |
| I69.811 | 0313091 |
| I69.812 | 0313092 |
| I69.813 | 0313093 |
| I69.814 | 0313094 |
| I69.815 | 0313095 |
| I69.818 | 0313096 |
| I69.819 | 0313097 |
| I69.820 | 0313098 |
| I69.821 | 0313099 |
| I69.822 | 031309B |
| I69.823 | 031309C |
| I69.828 | 031309J |
| I69.831 | 031309K |
| I69.832 | 03130A0 |
| I69.833 | 03130A1 |
| I69.834 | 03130A2 |
| I69.839 | 03130A3 |

| | |
|---|---|
| I69.841 | 03130A4 |
| I69.842 | 03130A5 |
| I69.843 | 03130A6 |
| I69.844 | 03130A7 |
| I69.849 | 03130A8 |
| I69.851 | 03130A9 |
| I69.852 | 03130AB |
| I69.853 | 03130AC |
| I69.854 | 03130AJ |
| I69.859 | 03130AK |
| I69.861 | 03130J0 |
| I69.862 | 03130J1 |
| I69.863 | 03130J2 |
| I69.864 | 03130J3 |
| I69.865 | 03130J4 |
| I69.869 | 03130J5 |
| I69.890 | 03130J6 |
| I69.891 | 03130J7 |
| I69.892 | 03130J8 |
| I69.893 | 03130J9 |
| I69.898 | 03130JB |
| I69.90 | 03130JC |
| I69.91 | 03130JJ |
| I69.910 | 03130JK |
| I69.911 | 03130K0 |
| I69.912 | 03130K1 |
| I69.913 | 03130K2 |
| I69.914 | 03130K3 |
| I69.915 | 03130K4 |
| I69.918 | 03130K5 |
| I69.919 | 03130K6 |
| I69.920 | 03130K7 |
| I69.921 | 03130K8 |
| I69.922 | 03130K9 |
| I69.923 | 03130KB |
| I69.928 | 03130KC |
| I69.931 | 03130KJ |
| I69.932 | 03130KK |
| I69.933 | 03130Z0 |
| I69.934 | 03130Z1 |
| I69.939 | 03130Z2 |
| I69.941 | 03130Z3 |
| I69.942 | 03130Z4 |

| | |
|---|---|
| I69.943 | 03130Z5 |
| I69.944 | 03130Z6 |
| I69.949 | 03130Z7 |
| I69.951 | 03130Z8 |
| I69.952 | 03130Z9 |
| I69.953 | 03130ZB |
| I69.954 | 03130ZC |
| I69.959 | 03130ZJ |
| I69.961 | 03130ZK |
| I69.962 | 0314090 |
| I69.963 | 0314091 |
| I69.964 | 0314092 |
| I69.965 | 0314093 |
| I69.969 | 0314094 |
| I69.990 | 0314095 |
| I69.991 | 0314096 |
| I69.992 | 0314097 |
| I69.993 | 0314098 |
| I69.998 | 0314099 |
| I70.0 | 031409B |
| I70.1 | 031409C |
| I70.201 | 031409J |
| I70.202 | 031409K |
| I70.203 | 03140A0 |
| I70.208 | 03140A1 |
| I70.209 | 03140A2 |
| I70.211 | 03140A3 |
| I70.212 | 03140A4 |
| I70.213 | 03140A5 |
| I70.218 | 03140A6 |
| I70.219 | 03140A7 |
| I70.221 | 03140A8 |
| I70.222 | 03140A9 |
| I70.223 | 03140AB |
| I70.228 | 03140AC |
| I70.229 | 03140AJ |
| I70.231 | 03140AK |
| I70.232 | 03140J0 |
| I70.233 | 03140J1 |
| I70.234 | 03140J2 |
| I70.235 | 03140J3 |
| I70.238 | 03140J4 |
| I70.239 | 03140J5 |

| | |
|---|---|
| I70.241 | 03140J6 |
| I70.242 | 03140J7 |
| I70.243 | 03140J8 |
| I70.244 | 03140J9 |
| I70.245 | 03140JB |
| I70.248 | 03140JC |
| I70.249 | 03140JJ |
| I70.25 | 03140JK |
| I70.261 | 03140K0 |
| I70.262 | 03140K1 |
| I70.263 | 03140K2 |
| I70.268 | 03140K3 |
| I70.269 | 03140K4 |
| I70.291 | 03140K5 |
| I70.292 | 03140K6 |
| I70.293 | 03140K7 |
| I70.298 | 03140K8 |
| I70.299 | 03140K9 |
| I70.301 | 03140KB |
| I70.302 | 03140KC |
| I70.303 | 03140KJ |
| I70.308 | 03140KK |
| I70.309 | 03140Z0 |
| I70.311 | 03140Z1 |
| I70.312 | 03140Z2 |
| I70.313 | 03140Z3 |
| I70.318 | 03140Z4 |
| I70.319 | 03140Z5 |
| I70.321 | 03140Z6 |
| I70.322 | 03140Z7 |
| I70.323 | 03140Z8 |
| I70.328 | 03140Z9 |
| I70.329 | 03140ZB |
| I70.331 | 03140ZC |
| I70.332 | 03140ZJ |
| I70.333 | 03140ZK |
| I70.334 | 0315090 |
| I70.335 | 0315091 |
| I70.338 | 0315092 |
| I70.339 | 0315093 |
| I70.341 | 0315094 |
| I70.342 | 0315095 |
| I70.343 | 0315096 |

| | |
|---|---|
| I70.344 | 0315097 |
| I70.345 | 0315098 |
| I70.348 | 0315099 |
| I70.349 | 031509B |
| I70.35 | 031509C |
| I70.361 | 031509J |
| I70.362 | 031509K |
| I70.363 | 03150A0 |
| I70.368 | 03150A1 |
| I70.369 | 03150A2 |
| I70.391 | 03150A3 |
| I70.392 | 03150A4 |
| I70.393 | 03150A5 |
| I70.398 | 03150A6 |
| I70.399 | 03150A7 |
| I70.401 | 03150A8 |
| I70.402 | 03150A9 |
| I70.403 | 03150AB |
| I70.408 | 03150AC |
| I70.409 | 03150AJ |
| I70.411 | 03150AK |
| I70.412 | 03150J0 |
| I70.413 | 03150J1 |
| I70.418 | 03150J2 |
| I70.419 | 03150J3 |
| I70.421 | 03150J4 |
| I70.422 | 03150J5 |
| I70.423 | 03150J6 |
| I70.428 | 03150J7 |
| I70.429 | 03150J8 |
| I70.431 | 03150J9 |
| I70.432 | 03150JB |
| I70.433 | 03150JC |
| I70.434 | 03150JJ |
| I70.435 | 03150JK |
| I70.438 | 03150K0 |
| I70.439 | 03150K1 |
| I70.441 | 03150K2 |
| I70.442 | 03150K3 |
| I70.443 | 03150K4 |
| I70.444 | 03150K5 |
| I70.445 | 03150K6 |
| I70.448 | 03150K7 |

| | |
|---|---|
| I70.449 | 03150K8 |
| I70.45 | 03150K9 |
| I70.461 | 03150KB |
| I70.462 | 03150KC |
| I70.463 | 03150KJ |
| I70.468 | 03150KK |
| I70.469 | 03150Z0 |
| I70.491 | 03150Z1 |
| I70.492 | 03150Z2 |
| I70.493 | 03150Z3 |
| I70.498 | 03150Z4 |
| I70.499 | 03150Z5 |
| I70.501 | 03150Z6 |
| I70.502 | 03150Z7 |
| I70.503 | 03150Z8 |
| I70.508 | 03150Z9 |
| I70.509 | 03150ZB |
| I70.511 | 03150ZC |
| I70.512 | 03150ZJ |
| I70.513 | 03150ZK |
| I70.518 | 0316090 |
| I70.519 | 0316091 |
| I70.521 | 0316092 |
| I70.522 | 0316093 |
| I70.523 | 0316094 |
| I70.528 | 0316095 |
| I70.529 | 0316096 |
| I70.531 | 0316097 |
| I70.532 | 0316098 |
| I70.533 | 0316099 |
| I70.534 | 031609B |
| I70.535 | 031609C |
| I70.538 | 031609J |
| I70.539 | 031609K |
| I70.541 | 03160A0 |
| I70.542 | 03160A1 |
| I70.543 | 03160A2 |
| I70.544 | 03160A3 |
| I70.545 | 03160A4 |
| I70.548 | 03160A5 |
| I70.549 | 03160A6 |
| I70.55 | 03160A7 |
| I70.561 | 03160A8 |

| | |
|---|---|
| I70.562 | 03160A9 |
| I70.563 | 03160AB |
| I70.568 | 03160AC |
| I70.569 | 03160AJ |
| I70.591 | 03160AK |
| I70.592 | 03160J0 |
| I70.593 | 03160J1 |
| I70.598 | 03160J2 |
| I70.599 | 03160J3 |
| I70.601 | 03160J4 |
| I70.602 | 03160J5 |
| I70.603 | 03160J6 |
| I70.608 | 03160J7 |
| I70.609 | 03160J8 |
| I70.611 | 03160J9 |
| I70.612 | 03160JB |
| I70.613 | 03160JC |
| I70.618 | 03160JJ |
| I70.619 | 03160JK |
| I70.621 | 03160K0 |
| I70.622 | 03160K1 |
| I70.623 | 03160K2 |
| I70.628 | 03160K3 |
| I70.629 | 03160K4 |
| I70.631 | 03160K5 |
| I70.632 | 03160K6 |
| I70.633 | 03160K7 |
| I70.634 | 03160K8 |
| I70.635 | 03160K9 |
| I70.638 | 03160KB |
| I70.639 | 03160KC |
| I70.641 | 03160KJ |
| I70.642 | 03160KK |
| I70.643 | 03160Z0 |
| I70.644 | 03160Z1 |
| I70.645 | 03160Z2 |
| I70.648 | 03160Z3 |
| I70.649 | 03160Z4 |
| I70.65 | 03160Z5 |
| I70.661 | 03160Z6 |
| I70.662 | 03160Z7 |
| I70.663 | 03160Z8 |
| I70.668 | 03160Z9 |

| | |
|---|---|
| I70.669 | 03160ZB |
| I70.691 | 03160ZC |
| I70.692 | 03160ZJ |
| I70.693 | 03160ZK |
| I70.698 | 0317090 |
| I70.699 | 0317093 |
| I70.701 | 03170A0 |
| I70.702 | 03170A3 |
| I70.703 | 03170J0 |
| I70.708 | 03170J3 |
| I70.709 | 03170K0 |
| I70.711 | 03170K3 |
| I70.712 | 03170Z0 |
| I70.713 | 03170Z3 |
| I70.718 | 0318091 |
| I70.719 | 0318094 |
| I70.721 | 03180A1 |
| I70.722 | 03180A4 |
| I70.723 | 03180J1 |
| I70.728 | 03180J4 |
| I70.729 | 03180K1 |
| I70.731 | 03180K4 |
| I70.732 | 03180Z1 |
| I70.733 | 03180Z4 |
| I70.734 | 0319093 |
| I70.735 | 03190A3 |
| I70.738 | 03190J3 |
| I70.739 | 03190K3 |
| I70.741 | 03190Z3 |
| I70.742 | 031A094 |
| I70.743 | 031A0A4 |
| I70.744 | 031A0J4 |
| I70.745 | 031A0K4 |
| I70.748 | 031A0Z4 |
| I70.749 | 031B093 |
| I70.75 | 031B0A3 |
| I70.761 | 031B0J3 |
| I70.762 | 031B0K3 |
| I70.763 | 031B0Z3 |
| I70.768 | 031C094 |
| I70.769 | 031C0A4 |
| I70.791 | 031C0J4 |
| I70.792 | 031C0K4 |

| | |
|---|---|
| I70.793 | 031C0Z4 |
| I70.798 | 031G09G |
| I70.799 | 031G0AG |
| I70.8 | 031G0JG |
| I70.90 | 031G0KG |
| I70.91 | 031G0ZG |
| I70.92 | 031H09G |
| I71.3 | 031H09J |
| I71.4 | 031H0AG |
| I71.5 | 031H0AJ |
| I71.6 | 031H0JG |
| I71.8 | 031H0JJ |
| I71.9 | 031H0KG |
| I73.9 | 031H0KJ |
| I74.01 | 031H0ZG |
| I74.09 | 031H0ZJ |
| I74.10 | 031J09G |
| I74.19 | 031J09K |
| I74.2 | 031J0AG |
| I74.3 | 031J0AK |
| I74.4 | 031J0JG |
| I74.5 | 031J0JK |
| I74.8 | 031J0KG |
| I75.011 | 031J0KK |
| I75.012 | 031J0ZG |
| I75.013 | 031J0ZK |
| I75.019 | 031K09J |
| I75.021 | 031K0AJ |
| I75.022 | 031K0JJ |
| I75.023 | 031K0KJ |
| I75.029 | 031K0ZJ |
| I79.0 | 031L09K |
| I79.1 | 031L0AK |
| I79.8 | 031L0JK |
| I97.810 | 031L0KK |
| I97.811 | 031L0ZK |
| I97.820 | 031M09J |
| I97.821 | 031M0AJ |
| O99.411 | 031M0JJ |
| O99.412 | 031M0KJ |
| O99.413 | 031M0ZJ |
| O99.419 | 031N09K |
| O99.42 | 031N0AK |

| | |
|---|---|
| O99.43 | 031N0JK |
| T82.211A | 031N0KK |
| T82.211D | 031N0ZK |
| T82.211S | 031S09G |
| T82.212A | 031S0AG |
| T82.212D | 031S0JG |
| T82.212S | 031S0KG |
| T82.213A | 031S0ZG |
| T82.213D | 031T09G |
| T82.213S | 031T0AG |
| T82.218A | 031T0JG |
| T82.218D | 031T0KG |
| T82.218S | 031T0ZG |
| T82.310A | 0372046 |
| T82.310D | 037204Z |
| T82.310S | 0372056 |
| T82.311A | 037205Z |
| T82.311D | 0372066 |
| T82.311S | 037206Z |
| T82.312A | 0372076 |
| T82.312D | 037207Z |
| T82.312S | 03720D6 |
| T82.320A | 03720DZ |
| T82.320D | 03720E6 |
| T82.320S | 03720EZ |
| T82.321A | 03720F6 |
| T82.321D | 03720FZ |
| T82.321S | 03720G6 |
| T82.322A | 03720GZ |
| T82.322D | 03720Z6 |
| T82.322S | 03720ZZ |
| T82.330A | 0372346 |
| T82.330D | 037234Z |
| T82.330S | 0372356 |
| T82.331A | 037235Z |
| T82.331D | 0372366 |
| T82.331S | 037236Z |
| T82.332A | 0372376 |
| T82.332D | 037237Z |
| T82.332S | 03723D6 |
| T82.390A | 03723DZ |
| T82.390D | 03723E6 |
| T82.390S | 03723EZ |

| | |
|---|---|
| T82.391A | 03723F6 |
| T82.391D | 03723FZ |
| T82.391S | 03723G6 |
| T82.392A | 03723GZ |
| T82.392D | 03723Z6 |
| T82.392S | 03723ZZ |
| T82.855A | 0372446 |
| T82.855D | 037244Z |
| T82.855S | 0372456 |
| Z86.73 | 037245Z |
| Z95.1 | 0372466 |
| Z95.5 | 037246Z |
| Z98.61 | 0372476 |
| | 037247Z |
| | 03724D6 |
| | 03724DZ |
| | 03724E6 |
| | 03724EZ |
| | 03724F6 |
| | 03724FZ |
| | 03724G6 |
| | 03724GZ |
| | 03724Z6 |
| | 03724ZZ |
| | 0373046 |
| | 037304Z |
| | 0373056 |
| | 037305Z |
| | 0373066 |
| | 037306Z |
| | 0373076 |
| | 037307Z |
| | 03730D6 |
| | 03730DZ |
| | 03730E6 |
| | 03730EZ |
| | 03730F6 |
| | 03730FZ |
| | 03730G6 |
| | 03730GZ |
| | 03730Z6 |
| | 03730ZZ |
| | 0373346 |

037334Z
0373356
037335Z
0373366
037336Z
0373376
037337Z
03733D6
03733DZ
03733E6
03733EZ
03733F6
03733FZ
03733G6
03733GZ
03733Z6
03733ZZ
0373446
037344Z
0373456
037345Z
0373466
037346Z
0373476
037347Z
03734D6
03734DZ
03734E6
03734EZ
03734F6
03734FZ
03734G6
03734GZ
03734Z6
03734ZZ
0374046
037404Z
0374056
037405Z
0374066
037406Z
0374076
037407Z

03740D6
03740DZ
03740E6
03740EZ
03740F6
03740FZ
03740G6
03740GZ
03740Z6
03740ZZ
0374346
037434Z
0374356
037435Z
0374366
037436Z
0374376
037437Z
03743D6
03743DZ
03743E6
03743EZ
03743F6
03743FZ
03743G6
03743GZ
03743Z6
03743ZZ
0374446
037444Z
0374456
037445Z
0374466
037446Z
0374476
037447Z
03744D6
03744DZ
03744E6
03744EZ
03744F6
03744FZ
03744G6

03744GZ
03744Z6
03744ZZ
0375046
037504Z
0375056
037505Z
0375066
037506Z
0375076
037507Z
03750D6
03750DZ
03750E6
03750EZ
03750F6
03750FZ
03750G6
03750GZ
03750Z6
03750ZZ
0375346
037534Z
0375356
037535Z
0375366
037536Z
0375376
037537Z
03753D6
03753DZ
03753E6
03753EZ
03753F6
03753FZ
03753G6
03753GZ
03753Z6
03753ZZ
0375446
037544Z
0375456
037545Z

0375466
037546Z
0375476
037547Z
03754D6
03754DZ
03754E6
03754EZ
03754F6
03754FZ
03754G6
03754GZ
03754Z6
03754ZZ
0376046
037604Z
0376056
037605Z
0376066
037606Z
0376076
037607Z
03760D6
03760DZ
03760E6
03760EZ
03760F6
03760FZ
03760G6
03760GZ
03760Z6
03760ZZ
0376346
037634Z
0376356
037635Z
0376366
037636Z
0376376
037637Z
03763D6
03763DZ
03763E6

03763EZ
03763F6
03763FZ
03763G6
03763GZ
03763Z6
03763ZZ
0376446
037644Z
0376456
037645Z
0376466
037646Z
0376476
037647Z
03764D6
03764DZ
03764E6
03764EZ
03764F6
03764FZ
03764G6
03764GZ
03764Z6
03764ZZ
0377046
037704Z
0377056
037705Z
0377066
037706Z
0377076
037707Z
03770D6
03770DZ
03770E6
03770EZ
03770F6
03770FZ
03770G6
03770GZ
03770Z6
03770ZZ

0377346
037734Z
0377356
037735Z
0377366
037736Z
0377376
037737Z
03773D6
03773DZ
03773E6
03773EZ
03773F6
03773FZ
03773G6
03773GZ
03773Z6
03773ZZ
0377446
037744Z
0377456
037745Z
0377466
037746Z
0377476
037747Z
03774D6
03774DZ
03774E6
03774EZ
03774F6
03774FZ
03774G6
03774GZ
03774Z6
03774ZZ
0378046
037804Z
0378056
037805Z
0378066
037806Z
0378076

037807Z
03780D6
03780DZ
03780E6
03780EZ
03780F6
03780FZ
03780G6
03780GZ
03780Z6
03780ZZ
0378346
037834Z
0378356
037835Z
0378366
037836Z
0378376
037837Z
03783D6
03783DZ
03783E6
03783EZ
03783F6
03783FZ
03783G6
03783GZ
03783Z6
03783ZZ
0378446
037844Z
0378456
037845Z
0378466
037846Z
0378476
037847Z
03784D6
03784DZ
03784E6
03784EZ
03784F6
03784FZ

03784G6
03784GZ
03784Z6
03784ZZ
0379046
037904Z
0379056
037905Z
0379066
037906Z
0379076
037907Z
03790D6
03790DZ
03790E6
03790EZ
03790F6
03790FZ
03790G6
03790GZ
03790Z6
03790ZZ
0379346
037934Z
0379356
037935Z
0379366
037936Z
0379376
037937Z
03793D6
03793DZ
03793E6
03793EZ
03793F6
03793FZ
03793G6
03793GZ
03793Z6
03793ZZ
0379446
037944Z
0379456

037945Z
0379466
037946Z
0379476
037947Z
03794D6
03794DZ
03794E6
03794EZ
03794F6
03794FZ
03794G6
03794GZ
03794Z6
03794ZZ
037A046
037A04Z
037A056
037A05Z
037A066
037A06Z
037A076
037A07Z
037A0D6
037A0DZ
037A0E6
037A0EZ
037A0F6
037A0FZ
037A0G6
037A0GZ
037A0Z6
037A0ZZ
037A346
037A34Z
037A356
037A35Z
037A366
037A36Z
037A376
037A37Z
037A3D6
037A3DZ

037A3E6
037A3EZ
037A3F6
037A3FZ
037A3G6
037A3GZ
037A3Z6
037A3ZZ
037A446
037A44Z
037A456
037A45Z
037A466
037A46Z
037A476
037A47Z
037A4D6
037A4DZ
037A4E6
037A4EZ
037A4F6
037A4FZ
037A4G6
037A4GZ
037A4Z6
037A4ZZ
037B046
037B04Z
037B056
037B05Z
037B066
037B06Z
037B076
037B07Z
037B0D6
037B0DZ
037B0E6
037B0EZ
037B0F6
037B0FZ
037B0G6
037B0GZ
037B0Z6

037B0ZZ
037B346
037B34Z
037B356
037B35Z
037B366
037B36Z
037B376
037B37Z
037B3D6
037B3DZ
037B3E6
037B3EZ
037B3F6
037B3FZ
037B3G6
037B3GZ
037B3Z6
037B3ZZ
037B446
037B44Z
037B456
037B45Z
037B466
037B46Z
037B476
037B47Z
037B4D6
037B4DZ
037B4E6
037B4EZ
037B4F6
037B4FZ
037B4G6
037B4GZ
037B4Z6
037B4ZZ
037C046
037C04Z
037C056
037C05Z
037C066
037C06Z

037C076
037C07Z
037C0D6
037C0DZ
037C0E6
037C0EZ
037C0F6
037C0FZ
037C0G6
037C0GZ
037C0Z6
037C0ZZ
037C346
037C34Z
037C356
037C35Z
037C366
037C36Z
037C376
037C37Z
037C3D6
037C3DZ
037C3E6
037C3EZ
037C3F6
037C3FZ
037C3G6
037C3GZ
037C3Z6
037C3ZZ
037C446
037C44Z
037C456
037C45Z
037C466
037C46Z
037C476
037C47Z
037C4D6
037C4DZ
037C4E6
037C4EZ
037C4F6

037C4FZ
037C4G6
037C4GZ
037C4Z6
037C4ZZ
037D046
037D04Z
037D056
037D05Z
037D066
037D06Z
037D076
037D07Z
037D0D6
037D0DZ
037D0E6
037D0EZ
037D0F6
037D0FZ
037D0G6
037D0GZ
037D0Z6
037D0ZZ
037D346
037D34Z
037D356
037D35Z
037D366
037D36Z
037D376
037D37Z
037D3D6
037D3DZ
037D3E6
037D3EZ
037D3F6
037D3FZ
037D3G6
037D3GZ
037D3Z6
037D3ZZ
037D446
037D44Z

037D456
037D45Z
037D466
037D46Z
037D476
037D47Z
037D4D6
037D4DZ
037D4E6
037D4EZ
037D4F6
037D4FZ
037D4G6
037D4GZ
037D4Z6
037D4ZZ
037F046
037F04Z
037F056
037F05Z
037F066
037F06Z
037F076
037F07Z
037F0D6
037F0DZ
037F0E6
037F0EZ
037F0F6
037F0FZ
037F0G6
037F0GZ
037F0Z6
037F0ZZ
037F346
037F34Z
037F356
037F35Z
037F366
037F36Z
037F376
037F37Z
037F3D6

037F3DZ
037F3E6
037F3EZ
037F3F6
037F3FZ
037F3G6
037F3GZ
037F3Z6
037F3ZZ
037F446
037F44Z
037F456
037F45Z
037F466
037F46Z
037F476
037F47Z
037F4D6
037F4DZ
037F4E6
037F4EZ
037F4F6
037F4FZ
037F4G6
037F4GZ
037F4Z6
037F4ZZ
037H046
037H04Z
037H056
037H05Z
037H066
037H06Z
037H076
037H07Z
037H0D6
037H0DZ
037H0E6
037H0EZ
037H0F6
037H0FZ
037H0G6
037H0GZ

037H0Z6
037H0ZZ
037H346
037H34Z
037H356
037H35Z
037H366
037H36Z
037H376
037H37Z
037H3D6
037H3DZ
037H3E6
037H3EZ
037H3F6
037H3FZ
037H3G6
037H3GZ
037H3Z6
037H3ZZ
037H446
037H44Z
037H456
037H45Z
037H466
037H46Z
037H476
037H47Z
037H4D6
037H4DZ
037H4E6
037H4EZ
037H4F6
037H4FZ
037H4G6
037H4GZ
037H4Z6
037H4ZZ
037J046
037J04Z
037J056
037J05Z
037J066

037J06Z
037J076
037J07Z
037J0D6
037J0DZ
037J0E6
037J0EZ
037J0F6
037J0FZ
037J0G6
037J0GZ
037J0Z6
037J0ZZ
037J346
037J34Z
037J356
037J35Z
037J366
037J36Z
037J376
037J37Z
037J3D6
037J3DZ
037J3E6
037J3EZ
037J3F6
037J3FZ
037J3G6
037J3GZ
037J3Z6
037J3ZZ
037J446
037J44Z
037J456
037J45Z
037J466
037J46Z
037J476
037J47Z
037J4D6
037J4DZ
037J4E6
037J4EZ

037J4F6
037J4FZ
037J4G6
037J4GZ
037J4Z6
037J4ZZ
037K046
037K04Z
037K056
037K05Z
037K066
037K06Z
037K076
037K07Z
037K0D6
037K0DZ
037K0E6
037K0EZ
037K0F6
037K0FZ
037K0G6
037K0GZ
037K0Z6
037K0ZZ
037K346
037K34Z
037K356
037K35Z
037K366
037K36Z
037K376
037K37Z
037K3D6
037K3DZ
037K3E6
037K3EZ
037K3F6
037K3FZ
037K3G6
037K3GZ
037K3Z6
037K3ZZ
037K446

037K44Z
037K456
037K45Z
037K466
037K46Z
037K476
037K47Z
037K4D6
037K4DZ
037K4E6
037K4EZ
037K4F6
037K4FZ
037K4G6
037K4GZ
037K4Z6
037K4ZZ
037L046
037L04Z
037L056
037L05Z
037L066
037L06Z
037L076
037L07Z
037L0D6
037L0DZ
037L0E6
037L0EZ
037L0F6
037L0FZ
037L0G6
037L0GZ
037L0Z6
037L0ZZ
037L346
037L34Z
037L356
037L35Z
037L366
037L36Z
037L376
037L37Z

037L3D6
037L3DZ
037L3E6
037L3EZ
037L3F6
037L3FZ
037L3G6
037L3GZ
037L3Z6
037L3ZZ
037L446
037L44Z
037L456
037L45Z
037L466
037L46Z
037L476
037L47Z
037L4D6
037L4DZ
037L4E6
037L4EZ
037L4F6
037L4FZ
037L4G6
037L4GZ
037L4Z6
037L4ZZ
037M046
037M04Z
037M056
037M05Z
037M066
037M06Z
037M076
037M07Z
037M0D6
037M0DZ
037M0E6
037M0EZ
037M0F6
037M0FZ
037M0G6

037M0GZ
037M0Z6
037M0ZZ
037M346
037M34Z
037M356
037M35Z
037M366
037M36Z
037M376
037M37Z
037M3D6
037M3DZ
037M3E6
037M3EZ
037M3F6
037M3FZ
037M3G6
037M3GZ
037M3Z6
037M3ZZ
037M446
037M44Z
037M456
037M45Z
037M466
037M46Z
037M476
037M47Z
037M4D6
037M4DZ
037M4E6
037M4EZ
037M4F6
037M4FZ
037M4G6
037M4GZ
037M4Z6
037M4ZZ
037N046
037N04Z
037N056
037N05Z

037N066
037N06Z
037N076
037N07Z
037N0D6
037N0DZ
037N0E6
037N0EZ
037N0F6
037N0FZ
037N0G6
037N0GZ
037N0Z6
037N0ZZ
037N346
037N34Z
037N356
037N35Z
037N366
037N36Z
037N376
037N37Z
037N3D6
037N3DZ
037N3E6
037N3EZ
037N3F6
037N3FZ
037N3G6
037N3GZ
037N3Z6
037N3ZZ
037N446
037N44Z
037N456
037N45Z
037N466
037N46Z
037N476
037N47Z
037N4D6
037N4DZ
037N4E6

037N4EZ
037N4F6
037N4FZ
037N4G6
037N4GZ
037N4Z6
037N4ZZ
037P046
037P04Z
037P056
037P05Z
037P066
037P06Z
037P076
037P07Z
037P0D6
037P0DZ
037P0E6
037P0EZ
037P0F6
037P0FZ
037P0G6
037P0GZ
037P0Z6
037P0ZZ
037P346
037P34Z
037P356
037P35Z
037P366
037P36Z
037P376
037P37Z
037P3D6
037P3DZ
037P3E6
037P3EZ
037P3F6
037P3FZ
037P3G6
037P3GZ
037P3Z6
037P3ZZ

037P446
037P44Z
037P456
037P45Z
037P466
037P46Z
037P476
037P47Z
037P4D6
037P4DZ
037P4E6
037P4EZ
037P4F6
037P4FZ
037P4G6
037P4GZ
037P4Z6
037P4ZZ
037Q046
037Q04Z
037Q056
037Q05Z
037Q066
037Q06Z
037Q076
037Q07Z
037Q0D6
037Q0DZ
037Q0E6
037Q0EZ
037Q0F6
037Q0FZ
037Q0G6
037Q0GZ
037Q0Z6
037Q0ZZ
037Q346
037Q34Z
037Q356
037Q35Z
037Q366
037Q36Z
037Q376

037Q37Z
037Q3D6
037Q3DZ
037Q3E6
037Q3EZ
037Q3F6
037Q3FZ
037Q3G6
037Q3GZ
037Q3Z6
037Q3ZZ
037Q446
037Q44Z
037Q456
037Q45Z
037Q466
037Q46Z
037Q476
037Q47Z
037Q4D6
037Q4DZ
037Q4E6
037Q4EZ
037Q4F6
037Q4FZ
037Q4G6
037Q4GZ
037Q4Z6
037Q4ZZ
037R046
037R04Z
037R056
037R05Z
037R066
037R06Z
037R076
037R07Z
037R0D6
037R0DZ
037R0E6
037R0EZ
037R0F6
037R0FZ

037R0G6
037R0GZ
037R0Z6
037R0ZZ
037R346
037R34Z
037R356
037R35Z
037R366
037R36Z
037R376
037R37Z
037R3D6
037R3DZ
037R3E6
037R3EZ
037R3F6
037R3FZ
037R3G6
037R3GZ
037R3Z6
037R3ZZ
037R446
037R44Z
037R456
037R45Z
037R466
037R46Z
037R476
037R47Z
037R4D6
037R4DZ
037R4E6
037R4EZ
037R4F6
037R4FZ
037R4G6
037R4GZ
037R4Z6
037R4ZZ
037S046
037S04Z
037S056

037S05Z
037S066
037S06Z
037S076
037S07Z
037S0D6
037S0DZ
037S0E6
037S0EZ
037S0F6
037S0FZ
037S0G6
037S0GZ
037S0Z6
037S0ZZ
037S346
037S34Z
037S356
037S35Z
037S366
037S36Z
037S376
037S37Z
037S3D6
037S3DZ
037S3E6
037S3EZ
037S3F6
037S3FZ
037S3G6
037S3GZ
037S3Z6
037S3ZZ
037S446
037S44Z
037S456
037S45Z
037S466
037S46Z
037S476
037S47Z
037S4D6
037S4DZ

037S4E6
037S4EZ
037S4F6
037S4FZ
037S4G6
037S4GZ
037S4Z6
037S4ZZ
037T046
037T04Z
037T056
037T05Z
037T066
037T06Z
037T076
037T07Z
037T0D6
037T0DZ
037T0E6
037T0EZ
037T0F6
037T0FZ
037T0G6
037T0GZ
037T0Z6
037T0ZZ
037T346
037T34Z
037T356
037T35Z
037T366
037T36Z
037T376
037T37Z
037T3D6
037T3DZ
037T3E6
037T3EZ
037T3F6
037T3FZ
037T3G6
037T3GZ
037T3Z6

037T3ZZ
037T446
037T44Z
037T456
037T45Z
037T466
037T46Z
037T476
037T47Z
037T4D6
037T4DZ
037T4E6
037T4EZ
037T4F6
037T4FZ
037T4G6
037T4GZ
037T4Z6
037T4ZZ
03C20Z6
03C20ZZ
03C23Z6
03C23ZZ
03C24Z6
03C24ZZ
03C30Z6
03C30ZZ
03C33Z6
03C33ZZ
03C34Z6
03C34ZZ
03C40Z6
03C40ZZ
03C43Z6
03C43ZZ
03C44Z6
03C44ZZ
03C50Z6
03C50ZZ
03C53Z6
03C53ZZ
03C54Z6
03C54ZZ

03C60Z6
03C60ZZ
03C63Z6
03C63ZZ
03C64Z6
03C64ZZ
03C70Z6
03C70ZZ
03C73Z6
03C73ZZ
03C74Z6
03C74ZZ
03C80Z6
03C80ZZ
03C83Z6
03C83ZZ
03C84Z6
03C84ZZ
03C90Z6
03C90ZZ
03C93Z6
03C93ZZ
03C94Z6
03C94ZZ
03CA0Z6
03CA0ZZ
03CA3Z6
03CA3ZZ
03CA4Z6
03CA4ZZ
03CB0Z6
03CB0ZZ
03CB3Z6
03CB3ZZ
03CB4Z6
03CB4ZZ
03CC0Z6
03CC0ZZ
03CC3Z6
03CC3ZZ
03CC4Z6
03CC4ZZ
03CD0Z6

03CD0ZZ
03CD3Z6
03CD3ZZ
03CD4Z6
03CD4ZZ
03CF0Z6
03CF0ZZ
03CF3Z6
03CF3ZZ
03CF4Z6
03CF4ZZ
03CG0Z6
03CG0ZZ
03CG3Z6
03CG3ZZ
03CG4Z6
03CG4ZZ
03CH0Z6
03CH0ZZ
03CH3Z6
03CH3ZZ
03CH4Z6
03CH4ZZ
03CJ0Z6
03CJ0ZZ
03CJ3Z6
03CJ3ZZ
03CJ4Z6
03CJ4ZZ
03CK0Z6
03CK0ZZ
03CK3Z6
03CK3ZZ
03CK4Z6
03CK4ZZ
03CL0Z6
03CL0ZZ
03CL3Z6
03CL3ZZ
03CL4Z6
03CL4ZZ
03CM0Z6
03CM0ZZ

03CM3Z6
03CM3ZZ
03CM4Z6
03CM4ZZ
03CN0Z6
03CN0ZZ
03CN3Z6
03CN3ZZ
03CN4Z6
03CN4ZZ
03CP0Z6
03CP0ZZ
03CP3Z6
03CP3ZZ
03CP4Z6
03CP4ZZ
03CQ0Z6
03CQ0ZZ
03CQ3Z6
03CQ3ZZ
03CQ4Z6
03CQ4ZZ
03CR0Z6
03CR0ZZ
03CR3Z6
03CR3ZZ
03CR4Z6
03CR4ZZ
03CS0Z6
03CS0ZZ
03CS3Z6
03CS3ZZ
03CS4Z6
03CS4ZZ
03CT0Z6
03CT0ZZ
03CT3Z6
03CT3ZZ
03CT4Z6
03CT4ZZ
03H20DZ
03H23DZ
03H24DZ

03H30DZ
03H33DZ
03H34DZ
03H40DZ
03H43DZ
03H44DZ
03H50DZ
03H53DZ
03H54DZ
03H60DZ
03H63DZ
03H64DZ
03H70DZ
03H73DZ
03H74DZ
03H80DZ
03H83DZ
03H84DZ
03H90DZ
03H93DZ
03H94DZ
03HA0DZ
03HA3DZ
03HA4DZ
03HB0DZ
03HB3DZ
03HB4DZ
03HC0DZ
03HC3DZ
03HC4DZ
03HD0DZ
03HD3DZ
03HD4DZ
03HF0DZ
03HF3DZ
03HF4DZ
03HG0DZ
03HG3DZ
03HG4DZ
03HH0DZ
03HH3DZ
03HH4DZ
03HJ0DZ

03HJ3DZ
03HJ4DZ
03HK0DZ
03HK3DZ
03HK4DZ
03HL0DZ
03HL3DZ
03HL4DZ
03HM0DZ
03HM3DZ
03HM4DZ
03HN0DZ
03HN3DZ
03HN4DZ
03HP0DZ
03HP3DZ
03HP4DZ
03HQ0DZ
03HQ3DZ
03HQ4DZ
03HR0DZ
03HR3DZ
03HR4DZ
03HS0DZ
03HS3DZ
03HS4DZ
03HT0DZ
03HT3DZ
03HT4DZ
03R207Z
03R20JZ
03R20KZ
03R247Z
03R24JZ
03R24KZ
03R307Z
03R30JZ
03R30KZ
03R347Z
03R34JZ
03R34KZ
03R407Z
03R40JZ

03R40KZ
03R447Z
03R44JZ
03R44KZ
03R507Z
03R50JZ
03R50KZ
03R547Z
03R54JZ
03R54KZ
03R607Z
03R60JZ
03R60KZ
03R647Z
03R64JZ
03R64KZ
03R707Z
03R70JZ
03R70KZ
03R747Z
03R74JZ
03R74KZ
03R807Z
03R80JZ
03R80KZ
03R847Z
03R84JZ
03R84KZ
03R907Z
03R90JZ
03R90KZ
03R947Z
03R94JZ
03R94KZ
03RA07Z
03RA0JZ
03RA0KZ
03RA47Z
03RA4JZ
03RA4KZ
03RB07Z
03RB0JZ
03RB0KZ

03RB47Z
03RB4JZ
03RB4KZ
03RC07Z
03RC0JZ
03RC0KZ
03RC47Z
03RC4JZ
03RC4KZ
03RD07Z
03RD0JZ
03RD0KZ
03RD47Z
03RD4JZ
03RD4KZ
03RF07Z
03RF0JZ
03RF0KZ
03RF47Z
03RF4JZ
03RF4KZ
03RG07Z
03RG0JZ
03RG0KZ
03RG47Z
03RG4JZ
03RG4KZ
03RH07Z
03RH0JZ
03RH0KZ
03RH47Z
03RH4JZ
03RH4KZ
03RJ07Z
03RJ0JZ
03RJ0KZ
03RJ47Z
03RJ4JZ
03RJ4KZ
03RK07Z
03RK0JZ
03RK0KZ
03RK47Z

03RK4JZ
03RK4KZ
03RL07Z
03RL0JZ
03RL0KZ
03RL47Z
03RL4JZ
03RL4KZ
03RM07Z
03RM0JZ
03RM0KZ
03RM47Z
03RM4JZ
03RM4KZ
03RN07Z
03RN0JZ
03RN0KZ
03RN47Z
03RN4JZ
03RN4KZ
03RP07Z
03RP0JZ
03RP0KZ
03RP47Z
03RP4JZ
03RP4KZ
03RQ07Z
03RQ0JZ
03RQ0KZ
03RQ47Z
03RQ4JZ
03RQ4KZ
03RR07Z
03RR0JZ
03RR0KZ
03RR47Z
03RR4JZ
03RR4KZ
03RS07Z
03RS0JZ
03RS0KZ
03RS47Z
03RS4JZ

03RS4KZ
03RT07Z
03RT0JZ
03RT0KZ
03RT47Z
03RT4JZ
03RT4KZ
03U207Z
03U20JZ
03U20KZ
03U237Z
03U23JZ
03U23KZ
03U247Z
03U24JZ
03U24KZ
03U307Z
03U30JZ
03U30KZ
03U337Z
03U33JZ
03U33KZ
03U347Z
03U34JZ
03U34KZ
03U407Z
03U40JZ
03U40KZ
03U437Z
03U43JZ
03U43KZ
03U447Z
03U44JZ
03U44KZ
03U507Z
03U50JZ
03U50KZ
03U537Z
03U53JZ
03U53KZ
03U547Z
03U54JZ
03U54KZ

03U607Z
03U60JZ
03U60KZ
03U637Z
03U63JZ
03U63KZ
03U647Z
03U64JZ
03U64KZ
03U707Z
03U70JZ
03U70KZ
03U737Z
03U73JZ
03U73KZ
03U747Z
03U74JZ
03U74KZ
03U807Z
03U80JZ
03U80KZ
03U837Z
03U83JZ
03U83KZ
03U847Z
03U84JZ
03U84KZ
03U907Z
03U90JZ
03U90KZ
03U937Z
03U93JZ
03U93KZ
03U947Z
03U94JZ
03U94KZ
03UA07Z
03UA0JZ
03UA0KZ
03UA37Z
03UA3JZ
03UA3KZ
03UA47Z

03UA4JZ
03UA4KZ
03UB07Z
03UB0JZ
03UB0KZ
03UB37Z
03UB3JZ
03UB3KZ
03UB47Z
03UB4JZ
03UB4KZ
03UC07Z
03UC0JZ
03UC0KZ
03UC37Z
03UC3JZ
03UC3KZ
03UC47Z
03UC4JZ
03UC4KZ
03UD07Z
03UD0JZ
03UD0KZ
03UD37Z
03UD3JZ
03UD3KZ
03UD47Z
03UD4JZ
03UD4KZ
03UF07Z
03UF0JZ
03UF0KZ
03UF37Z
03UF3JZ
03UF3KZ
03UF47Z
03UF4JZ
03UF4KZ
03UG07Z
03UG0JZ
03UG0KZ
03UG37Z
03UG3JZ

03UG3KZ
03UG47Z
03UG4JZ
03UG4KZ
03UH07Z
03UH0JZ
03UH0KZ
03UH37Z
03UH3JZ
03UH3KZ
03UH47Z
03UH4JZ
03UH4KZ
03UJ07Z
03UJ0JZ
03UJ0KZ
03UJ37Z
03UJ3JZ
03UJ3KZ
03UJ47Z
03UJ4JZ
03UJ4KZ
03UK07Z
03UK0JZ
03UK0KZ
03UK37Z
03UK3JZ
03UK3KZ
03UK47Z
03UK4JZ
03UK4KZ
03UL07Z
03UL0JZ
03UL0KZ
03UL37Z
03UL3JZ
03UL3KZ
03UL47Z
03UL4JZ
03UL4KZ
03UM07Z
03UM0JZ
03UM0KZ

03UM37Z
03UM3JZ
03UM3KZ
03UM47Z
03UM4JZ
03UM4KZ
03UN07Z
03UN0JZ
03UN0KZ
03UN37Z
03UN3JZ
03UN3KZ
03UN47Z
03UN4JZ
03UN4KZ
03UP07Z
03UP0JZ
03UP0KZ
03UP37Z
03UP3JZ
03UP3KZ
03UP47Z
03UP4JZ
03UP4KZ
03UQ07Z
03UQ0JZ
03UQ0KZ
03UQ37Z
03UQ3JZ
03UQ3KZ
03UQ47Z
03UQ4JZ
03UQ4KZ
03UR07Z
03UR0JZ
03UR0KZ
03UR37Z
03UR3JZ
03UR3KZ
03UR47Z
03UR4JZ
03UR4KZ
03US07Z

03US0JZ
03US0KZ
03US37Z
03US3JZ
03US3KZ
03US47Z
03US4JZ
03US4KZ
03UT07Z
03UT0JZ
03UT0KZ
03UT37Z
03UT3JZ
03UT3KZ
03UT47Z
03UT4JZ
03UT4KZ
0410090
0410091
0410092
0410093
0410094
0410095
0410096
0410097
0410098
0410099
041009B
041009C
041009D
041009F
041009G
041009H
041009J
041009K
041009Q
041009R
04100A0
04100A1
04100A2
04100A3
04100A4
04100A5

04100A6
04100A7
04100A8
04100A9
04100AB
04100AC
04100AD
04100AF
04100AG
04100AH
04100AJ
04100AK
04100AQ
04100AR
04100J0
04100J1
04100J2
04100J3
04100J4
04100J5
04100J6
04100J7
04100J8
04100J9
04100JB
04100JC
04100JD
04100JF
04100JG
04100JH
04100JJ
04100JK
04100JQ
04100JR
04100K0
04100K1
04100K2
04100K3
04100K4
04100K5
04100K6
04100K7
04100K8

04100K9
04100KB
04100KC
04100KD
04100KF
04100KG
04100KH
04100KJ
04100KK
04100KQ
04100KR
04100Z0
04100Z1
04100Z2
04100Z3
04100Z4
04100Z5
04100Z6
04100Z7
04100Z8
04100Z9
04100ZB
04100ZC
04100ZD
04100ZF
04100ZG
04100ZH
04100ZJ
04100ZK
04100ZQ
04100ZR
0410490
0410491
0410492
0410493
0410494
0410495
0410496
0410497
0410498
0410499
041049B
041049C

041049D
041049F
041049G
041049H
041049J
041049K
041049Q
041049R
04104A0
04104A1
04104A2
04104A3
04104A4
04104A5
04104A6
04104A7
04104A8
04104A9
04104AB
04104AC
04104AD
04104AF
04104AG
04104AH
04104AJ
04104AK
04104AQ
04104AR
04104J0
04104J1
04104J2
04104J3
04104J4
04104J5
04104J6
04104J7
04104J8
04104J9
04104JB
04104JC
04104JD
04104JF
04104JG

04104JH
04104JJ
04104JK
04104JQ
04104JR
04104K0
04104K1
04104K2
04104K3
04104K4
04104K5
04104K6
04104K7
04104K8
04104K9
04104KB
04104KC
04104KD
04104KF
04104KG
04104KH
04104KJ
04104KK
04104KQ
04104KR
04104Z0
04104Z1
04104Z2
04104Z3
04104Z4
04104Z5
04104Z6
04104Z7
04104Z8
04104Z9
04104ZB
04104ZC
04104ZD
04104ZF
04104ZG
04104ZH
04104ZJ
04104ZK

04104ZQ
04104ZR
0414093
0414094
0414095
04140A3
04140A4
04140A5
04140J3
04140J4
04140J5
04140K3
04140K4
04140K5
04140Z3
04140Z4
04140Z5
0414493
0414494
0414495
04144A3
04144A4
04144A5
04144J3
04144J4
04144J5
04144K3
04144K4
04144K5
04144Z3
04144Z4
04144Z5
041C090
041C091
041C092
041C093
041C094
041C095
041C096
041C097
041C098
041C099
041C09B

041C09C
041C09D
041C09F
041C09G
041C09H
041C09J
041C09K
041C09Q
041C09R
041C0A0
041C0A1
041C0A2
041C0A3
041C0A4
041C0A5
041C0A6
041C0A7
041C0A8
041C0A9
041C0AB
041C0AC
041C0AD
041C0AF
041C0AG
041C0AH
041C0AJ
041C0AK
041C0AQ
041C0AR
041C0J0
041C0J1
041C0J2
041C0J3
041C0J4
041C0J5
041C0J6
041C0J7
041C0J8
041C0J9
041C0JB
041C0JC
041C0JD
041C0JF

041C0JG
041C0JH
041C0JJ
041C0JK
041C0JQ
041C0JR
041C0K0
041C0K1
041C0K2
041C0K3
041C0K4
041C0K5
041C0K6
041C0K7
041C0K8
041C0K9
041C0KB
041C0KC
041C0KD
041C0KF
041C0KG
041C0KH
041C0KJ
041C0KK
041C0KQ
041C0KR
041C0Z0
041C0Z1
041C0Z2
041C0Z3
041C0Z4
041C0Z5
041C0Z6
041C0Z7
041C0Z8
041C0Z9
041C0ZB
041C0ZC
041C0ZD
041C0ZF
041C0ZG
041C0ZH
041C0ZJ

041C0ZK
041C0ZQ
041C0ZR
041C490
041C491
041C492
041C493
041C494
041C495
041C496
041C497
041C498
041C499
041C49B
041C49C
041C49D
041C49F
041C49G
041C49H
041C49J
041C49K
041C49Q
041C49R
041C4A0
041C4A1
041C4A2
041C4A3
041C4A4
041C4A5
041C4A6
041C4A7
041C4A8
041C4A9
041C4AB
041C4AC
041C4AD
041C4AF
041C4AG
041C4AH
041C4AJ
041C4AK
041C4AQ
041C4AR

041C4J0
041C4J1
041C4J2
041C4J3
041C4J4
041C4J5
041C4J6
041C4J7
041C4J8
041C4J9
041C4JB
041C4JC
041C4JD
041C4JF
041C4JG
041C4JH
041C4JJ
041C4JK
041C4JQ
041C4JR
041C4K0
041C4K1
041C4K2
041C4K3
041C4K4
041C4K5
041C4K6
041C4K7
041C4K8
041C4K9
041C4KB
041C4KC
041C4KD
041C4KF
041C4KG
041C4KH
041C4KJ
041C4KK
041C4KQ
041C4KR
041C4Z0
041C4Z1
041C4Z2

041C4Z3
041C4Z4
041C4Z5
041C4Z6
041C4Z7
041C4Z8
041C4Z9
041C4ZB
041C4ZC
041C4ZD
041C4ZF
041C4ZG
041C4ZH
041C4ZJ
041C4ZK
041C4ZQ
041C4ZR
041D090
041D091
041D092
041D093
041D094
041D095
041D096
041D097
041D098
041D099
041D09B
041D09C
041D09D
041D09F
041D09G
041D09H
041D09J
041D09K
041D09Q
041D09R
041D0A0
041D0A1
041D0A2
041D0A3
041D0A4
041D0A5

041D0A6
041D0A7
041D0A8
041D0A9
041D0AB
041D0AC
041D0AD
041D0AF
041D0AG
041D0AH
041D0AJ
041D0AK
041D0AQ
041D0AR
041D0J0
041D0J1
041D0J2
041D0J3
041D0J4
041D0J5
041D0J6
041D0J7
041D0J8
041D0J9
041D0JB
041D0JC
041D0JD
041D0JF
041D0JG
041D0JH
041D0JJ
041D0JK
041D0JQ
041D0JR
041D0K0
041D0K1
041D0K2
041D0K3
041D0K4
041D0K5
041D0K6
041D0K7
041D0K8

041D0K9
041D0KB
041D0KC
041D0KD
041D0KF
041D0KG
041D0KH
041D0KJ
041D0KK
041D0KQ
041D0KR
041D0Z0
041D0Z1
041D0Z2
041D0Z3
041D0Z4
041D0Z5
041D0Z6
041D0Z7
041D0Z8
041D0Z9
041D0ZB
041D0ZC
041D0ZD
041D0ZF
041D0ZG
041D0ZH
041D0ZJ
041D0ZK
041D0ZQ
041D0ZR
041D490
041D491
041D492
041D493
041D494
041D495
041D496
041D497
041D498
041D499
041D49B
041D49C

041D49D
041D49F
041D49G
041D49H
041D49J
041D49K
041D49Q
041D49R
041D4A0
041D4A1
041D4A2
041D4A3
041D4A4
041D4A5
041D4A6
041D4A7
041D4A8
041D4A9
041D4AB
041D4AC
041D4AD
041D4AF
041D4AG
041D4AH
041D4AJ
041D4AK
041D4AQ
041D4AR
041D4J0
041D4J1
041D4J2
041D4J3
041D4J4
041D4J5
041D4J6
041D4J7
041D4J8
041D4J9
041D4JB
041D4JC
041D4JD
041D4JF
041D4JG

041D4JH
041D4JJ
041D4JK
041D4JQ
041D4JR
041D4K0
041D4K1
041D4K2
041D4K3
041D4K4
041D4K5
041D4K6
041D4K7
041D4K8
041D4K9
041D4KB
041D4KC
041D4KD
041D4KF
041D4KG
041D4KH
041D4KJ
041D4KK
041D4KQ
041D4KR
041D4Z0
041D4Z1
041D4Z2
041D4Z3
041D4Z4
041D4Z5
041D4Z6
041D4Z7
041D4Z8
041D4Z9
041D4ZB
041D4ZC
041D4ZD
041D4ZF
041D4ZG
041D4ZH
041D4ZJ
041D4ZK

041D4ZQ
041D4ZR
041E099
041E09B
041E09C
041E09D
041E09F
041E09G
041E09H
041E09J
041E09K
041E09P
041E09Q
041E0A9
041E0AB
041E0AC
041E0AD
041E0AF
041E0AG
041E0AH
041E0AJ
041E0AK
041E0AP
041E0AQ
041E0J9
041E0JB
041E0JC
041E0JD
041E0JF
041E0JG
041E0JH
041E0JJ
041E0JK
041E0JP
041E0JQ
041E0K9
041E0KB
041E0KC
041E0KD
041E0KF
041E0KG
041E0KH
041E0KJ

041E0KK
041E0KP
041E0KQ
041E0Z9
041E0ZB
041E0ZC
041E0ZD
041E0ZF
041E0ZG
041E0ZH
041E0ZJ
041E0ZK
041E0ZP
041E0ZQ
041E499
041E49B
041E49C
041E49D
041E49F
041E49G
041E49H
041E49J
041E49K
041E49P
041E49Q
041E4A9
041E4AB
041E4AC
041E4AD
041E4AF
041E4AG
041E4AH
041E4AJ
041E4AK
041E4AP
041E4AQ
041E4J9
041E4JB
041E4JC
041E4JD
041E4JF
041E4JG
041E4JH

041E4JJ
041E4JK
041E4JP
041E4JQ
041E4K9
041E4KB
041E4KC
041E4KD
041E4KF
041E4KG
041E4KH
041E4KJ
041E4KK
041E4KP
041E4KQ
041E4Z9
041E4ZB
041E4ZC
041E4ZD
041E4ZF
041E4ZG
041E4ZH
041E4ZJ
041E4ZK
041E4ZP
041E4ZQ
041F099
041F09B
041F09C
041F09D
041F09F
041F09G
041F09H
041F09J
041F09K
041F09P
041F09Q
041F0A9
041F0AB
041F0AC
041F0AD
041F0AF
041F0AG

041F0AH
041F0AJ
041F0AK
041F0AP
041F0AQ
041F0J9
041F0JB
041F0JC
041F0JD
041F0JF
041F0JG
041F0JH
041F0JJ
041F0JK
041F0JP
041F0JQ
041F0K9
041F0KB
041F0KC
041F0KD
041F0KF
041F0KG
041F0KH
041F0KJ
041F0KK
041F0KP
041F0KQ
041F0Z9
041F0ZB
041F0ZC
041F0ZD
041F0ZF
041F0ZG
041F0ZH
041F0ZJ
041F0ZK
041F0ZP
041F0ZQ
041F499
041F49B
041F49C
041F49D
041F49F

041F49G
041F49H
041F49J
041F49K
041F49P
041F49Q
041F4A9
041F4AB
041F4AC
041F4AD
041F4AF
041F4AG
041F4AH
041F4AJ
041F4AK
041F4AP
041F4AQ
041F4J9
041F4JB
041F4JC
041F4JD
041F4JF
041F4JG
041F4JH
041F4JJ
041F4JK
041F4JP
041F4JQ
041F4K9
041F4KB
041F4KC
041F4KD
041F4KF
041F4KG
041F4KH
041F4KJ
041F4KK
041F4KP
041F4KQ
041F4Z9
041F4ZB
041F4ZC
041F4ZD

041F4ZF
041F4ZG
041F4ZH
041F4ZJ
041F4ZK
041F4ZP
041F4ZQ
041H099
041H09B
041H09C
041H09D
041H09F
041H09G
041H09H
041H09J
041H09K
041H09P
041H09Q
041H0A9
041H0AB
041H0AC
041H0AD
041H0AF
041H0AG
041H0AH
041H0AJ
041H0AK
041H0AP
041H0AQ
041H0J9
041H0JB
041H0JC
041H0JD
041H0JF
041H0JG
041H0JH
041H0JJ
041H0JK
041H0JP
041H0JQ
041H0K9
041H0KB
041H0KC

041H0KD
041H0KF
041H0KG
041H0KH
041H0KJ
041H0KK
041H0KP
041H0KQ
041H0Z9
041H0ZB
041H0ZC
041H0ZD
041H0ZF
041H0ZG
041H0ZH
041H0ZJ
041H0ZK
041H0ZP
041H0ZQ
041H499
041H49B
041H49C
041H49D
041H49F
041H49G
041H49H
041H49J
041H49K
041H49P
041H49Q
041H4A9
041H4AB
041H4AC
041H4AD
041H4AF
041H4AG
041H4AH
041H4AJ
041H4AK
041H4AP
041H4AQ
041H4J9
041H4JB

041H4JC
041H4JD
041H4JF
041H4JG
041H4JH
041H4JJ
041H4JK
041H4JP
041H4JQ
041H4K9
041H4KB
041H4KC
041H4KD
041H4KF
041H4KG
041H4KH
041H4KJ
041H4KK
041H4KP
041H4KQ
041H4Z9
041H4ZB
041H4ZC
041H4ZD
041H4ZF
041H4ZG
041H4ZH
041H4ZJ
041H4ZK
041H4ZP
041H4ZQ
041J099
041J09B
041J09C
041J09D
041J09F
041J09G
041J09H
041J09J
041J09K
041J09P
041J09Q
041J0A9

041J0AB
041J0AC
041J0AD
041J0AF
041J0AG
041J0AH
041J0AJ
041J0AK
041J0AP
041J0AQ
041J0J9
041J0JB
041J0JC
041J0JD
041J0JF
041J0JG
041J0JH
041J0JJ
041J0JK
041J0JP
041J0JQ
041J0K9
041J0KB
041J0KC
041J0KD
041J0KF
041J0KG
041J0KH
041J0KJ
041J0KK
041J0KP
041J0KQ
041J0Z9
041J0ZB
041J0ZC
041J0ZD
041J0ZF
041J0ZG
041J0ZH
041J0ZJ
041J0ZK
041J0ZP
041J0ZQ

041J499
041J49B
041J49C
041J49D
041J49F
041J49G
041J49H
041J49J
041J49K
041J49P
041J49Q
041J4A9
041J4AB
041J4AC
041J4AD
041J4AF
041J4AG
041J4AH
041J4AJ
041J4AK
041J4AP
041J4AQ
041J4J9
041J4JB
041J4JC
041J4JD
041J4JF
041J4JG
041J4JH
041J4JJ
041J4JK
041J4JP
041J4JQ
041J4K9
041J4KB
041J4KC
041J4KD
041J4KF
041J4KG
041J4KH
041J4KJ
041J4KK
041J4KP

041J4KQ
041J4Z9
041J4ZB
041J4ZC
041J4ZD
041J4ZF
041J4ZG
041J4ZH
041J4ZJ
041J4ZK
041J4ZP
041J4ZQ
041K09H
041K09J
041K09K
041K09L
041K09M
041K09N
041K09P
041K09Q
041K0AH
041K0AJ
041K0AK
041K0AL
041K0AM
041K0AN
041K0AP
041K0AQ
041K0JH
041K0JJ
041K0JK
041K0JL
041K0JM
041K0JN
041K0JP
041K0JQ
041K0KH
041K0KJ
041K0KK
041K0KL
041K0KM
041K0KN
041K0KP

041K0KQ
041K0ZH
041K0ZJ
041K0ZK
041K0ZL
041K0ZM
041K0ZN
041K0ZP
041K0ZQ
041K49H
041K49J
041K49K
041K49L
041K49M
041K49N
041K49P
041K49Q
041K4AH
041K4AJ
041K4AK
041K4AL
041K4AM
041K4AN
041K4AP
041K4AQ
041K4JH
041K4JJ
041K4JK
041K4JL
041K4JM
041K4JN
041K4JP
041K4JQ
041K4KH
041K4KJ
041K4KK
041K4KL
041K4KM
041K4KN
041K4KP
041K4KQ
041K4ZH
041K4ZJ

041K4ZK
041K4ZL
041K4ZM
041K4ZN
041K4ZP
041K4ZQ
041L09H
041L09J
041L09K
041L09L
041L09M
041L09N
041L09P
041L09Q
041L0AH
041L0AJ
041L0AK
041L0AL
041L0AM
041L0AN
041L0AP
041L0AQ
041L0JH
041L0JJ
041L0JK
041L0JL
041L0JM
041L0JN
041L0JP
041L0JQ
041L0KH
041L0KJ
041L0KK
041L0KL
041L0KM
041L0KN
041L0KP
041L0KQ
041L0ZH
041L0ZJ
041L0ZK
041L0ZL
041L0ZM

041L0ZN
041L0ZP
041L0ZQ
041L49H
041L49J
041L49K
041L49L
041L49M
041L49N
041L49P
041L49Q
041L4AH
041L4AJ
041L4AK
041L4AL
041L4AM
041L4AN
041L4AP
041L4AQ
041L4JH
041L4JJ
041L4JK
041L4JL
041L4JM
041L4JN
041L4JP
041L4JQ
041L4KH
041L4KJ
041L4KK
041L4KL
041L4KM
041L4KN
041L4KP
041L4KQ
041L4ZH
041L4ZJ
041L4ZK
041L4ZL
041L4ZM
041L4ZN
041L4ZP
041L4ZQ

041M09L
041M09M
041M09P
041M09Q
041M0AL
041M0AM
041M0AP
041M0AQ
041M0JL
041M0JM
041M0JP
041M0JQ
041M0KL
041M0KM
041M0KP
041M0KQ
041M0ZL
041M0ZM
041M0ZP
041M0ZQ
041M49L
041M49M
041M49P
041M49Q
041M4AL
041M4AM
041M4AP
041M4AQ
041M4JL
041M4JM
041M4JP
041M4JQ
041M4KL
041M4KM
041M4KP
041M4KQ
041M4ZL
041M4ZM
041M4ZP
041M4ZQ
041N09L
041N09M
041N09P

041N09Q
041N0AL
041N0AM
041N0AP
041N0AQ
041N0JL
041N0JM
041N0JP
041N0JQ
041N0KL
041N0KM
041N0KP
041N0KQ
041N0ZL
041N0ZM
041N0ZP
041N0ZQ
041N49L
041N49M
041N49P
041N49Q
041N4AL
041N4AM
041N4AP
041N4AQ
041N4JL
041N4JM
041N4JP
041N4JQ
041N4KL
041N4KM
041N4KP
041N4KQ
041N4ZL
041N4ZM
041N4ZP
041N4ZQ
0470046
047004Z
0470056
047005Z
0470066
047006Z

0470076
047007Z
04700D6
04700DZ
04700E6
04700EZ
04700F6
04700FZ
04700G6
04700GZ
04700Z6
04700ZZ
0470346
047034Z
0470356
047035Z
0470366
047036Z
0470376
047037Z
04703D6
04703DZ
04703E6
04703EZ
04703F6
04703FZ
04703G6
04703GZ
04703Z6
04703ZZ
0470446
047044Z
0470456
047045Z
0470466
047046Z
0470476
047047Z
04704D6
04704DZ
04704E6
04704EZ
04704F6

04704FZ
04704G6
04704GZ
04704Z6
04704ZZ
0471046
047104Z
0471056
047105Z
0471066
047106Z
0471076
047107Z
04710D6
04710DZ
04710E6
04710EZ
04710F6
04710FZ
04710G6
04710GZ
04710Z6
04710ZZ
0471346
047134Z
0471356
047135Z
0471366
047136Z
0471376
047137Z
04713D6
04713DZ
04713E6
04713EZ
04713F6
04713FZ
04713G6
04713GZ
04713Z6
04713ZZ
0471446
047144Z

0471456
047145Z
0471466
047146Z
0471476
047147Z
04714D6
04714DZ
04714E6
04714EZ
04714F6
04714FZ
04714G6
04714GZ
04714Z6
04714ZZ
0472046
047204Z
0472056
047205Z
0472066
047206Z
0472076
047207Z
04720D6
04720DZ
04720E6
04720EZ
04720F6
04720FZ
04720G6
04720GZ
04720Z6
04720ZZ
0472346
047234Z
0472356
047235Z
0472366
047236Z
0472376
047237Z
04723D6

04723DZ
04723E6
04723EZ
04723F6
04723FZ
04723G6
04723GZ
04723Z6
04723ZZ
0472446
047244Z
0472456
047245Z
0472466
047246Z
0472476
047247Z
04724D6
04724DZ
04724E6
04724EZ
04724F6
04724FZ
04724G6
04724GZ
04724Z6
04724ZZ
0473046
047304Z
0473056
047305Z
0473066
047306Z
0473076
047307Z
04730D6
04730DZ
04730E6
04730EZ
04730F6
04730FZ
04730G6
04730GZ

04730Z6
04730ZZ
0473346
047334Z
0473356
047335Z
0473366
047336Z
0473376
047337Z
04733D6
04733DZ
04733E6
04733EZ
04733F6
04733FZ
04733G6
04733GZ
04733Z6
04733ZZ
0473446
047344Z
0473456
047345Z
0473466
047346Z
0473476
047347Z
04734D6
04734DZ
04734E6
04734EZ
04734F6
04734FZ
04734G6
04734GZ
04734Z6
04734ZZ
0474046
047404Z
0474056
047405Z
0474066

047406Z
0474076
047407Z
04740D6
04740DZ
04740E6
04740EZ
04740F6
04740FZ
04740G6
04740GZ
04740Z6
04740ZZ
0474346
047434Z
0474356
047435Z
0474366
047436Z
0474376
047437Z
04743D6
04743DZ
04743E6
04743EZ
04743F6
04743FZ
04743G6
04743GZ
04743Z6
04743ZZ
0474446
047444Z
0474456
047445Z
0474466
047446Z
0474476
047447Z
04744D6
04744DZ
04744E6
04744EZ

04744F6
04744FZ
04744G6
04744GZ
04744Z6
04744ZZ
0475046
047504Z
0475056
047505Z
0475066
047506Z
0475076
047507Z
04750D6
04750DZ
04750E6
04750EZ
04750F6
04750FZ
04750G6
04750GZ
04750Z6
04750ZZ
0475346
047534Z
0475356
047535Z
0475366
047536Z
0475376
047537Z
04753D6
04753DZ
04753E6
04753EZ
04753F6
04753FZ
04753G6
04753GZ
04753Z6
04753ZZ
0475446

047544Z
0475456
047545Z
0475466
047546Z
0475476
047547Z
04754D6
04754DZ
04754E6
04754EZ
04754F6
04754FZ
04754G6
04754GZ
04754Z6
04754ZZ
0476046
047604Z
0476056
047605Z
0476066
047606Z
0476076
047607Z
04760D6
04760DZ
04760E6
04760EZ
04760F6
04760FZ
04760G6
04760GZ
04760Z6
04760ZZ
0476346
047634Z
0476356
047635Z
0476366
047636Z
0476376
047637Z

04763D6
04763DZ
04763E6
04763EZ
04763F6
04763FZ
04763G6
04763GZ
04763Z6
04763ZZ
0476446
047644Z
0476456
047645Z
0476466
047646Z
0476476
047647Z
04764D6
04764DZ
04764E6
04764EZ
04764F6
04764FZ
04764G6
04764GZ
04764Z6
04764ZZ
0477046
047704Z
0477056
047705Z
0477066
047706Z
0477076
047707Z
04770D6
04770DZ
04770E6
04770EZ
04770F6
04770FZ
04770G6

04770GZ
04770Z6
04770ZZ
0477346
047734Z
0477356
047735Z
0477366
047736Z
0477376
047737Z
04773D6
04773DZ
04773E6
04773EZ
04773F6
04773FZ
04773G6
04773GZ
04773Z6
04773ZZ
0477446
047744Z
0477456
047745Z
0477466
047746Z
0477476
047747Z
04774D6
04774DZ
04774E6
04774EZ
04774F6
04774FZ
04774G6
04774GZ
04774Z6
04774ZZ
0478046
047804Z
0478056
047805Z

0478066
047806Z
0478076
047807Z
04780D6
04780DZ
04780E6
04780EZ
04780F6
04780FZ
04780G6
04780GZ
04780Z6
04780ZZ
0478346
047834Z
0478356
047835Z
0478366
047836Z
0478376
047837Z
04783D6
04783DZ
04783E6
04783EZ
04783F6
04783FZ
04783G6
04783GZ
04783Z6
04783ZZ
0478446
047844Z
0478456
047845Z
0478466
047846Z
0478476
047847Z
04784D6
04784DZ
04784E6

04784EZ
04784F6
04784FZ
04784G6
04784GZ
04784Z6
04784ZZ
0479046
047904Z
0479056
047905Z
0479066
047906Z
0479076
047907Z
04790D6
04790DZ
04790E6
04790EZ
04790F6
04790FZ
04790G6
04790GZ
04790Z6
04790ZZ
0479346
047934Z
0479356
047935Z
0479366
047936Z
0479376
047937Z
04793D6
04793DZ
04793E6
04793EZ
04793F6
04793FZ
04793G6
04793GZ
04793Z6
04793ZZ

0479446
047944Z
0479456
047945Z
0479466
047946Z
0479476
047947Z
04794D6
04794DZ
04794E6
04794EZ
04794F6
04794FZ
04794G6
04794GZ
04794Z6
04794ZZ
047A046
047A04Z
047A056
047A05Z
047A066
047A06Z
047A076
047A07Z
047A0D6
047A0DZ
047A0E6
047A0EZ
047A0F6
047A0FZ
047A0G6
047A0GZ
047A0Z6
047A0ZZ
047A346
047A34Z
047A356
047A35Z
047A366
047A36Z
047A376

047A37Z
047A3D6
047A3DZ
047A3E6
047A3EZ
047A3F6
047A3FZ
047A3G6
047A3GZ
047A3Z6
047A3ZZ
047A446
047A44Z
047A456
047A45Z
047A466
047A46Z
047A476
047A47Z
047A4D6
047A4DZ
047A4E6
047A4EZ
047A4F6
047A4FZ
047A4G6
047A4GZ
047A4Z6
047A4ZZ
047B046
047B04Z
047B056
047B05Z
047B066
047B06Z
047B076
047B07Z
047B0D6
047B0DZ
047B0E6
047B0EZ
047B0F6
047B0FZ

047B0G6
047B0GZ
047B0Z6
047B0ZZ
047B346
047B34Z
047B356
047B35Z
047B366
047B36Z
047B376
047B37Z
047B3D6
047B3DZ
047B3E6
047B3EZ
047B3F6
047B3FZ
047B3G6
047B3GZ
047B3Z6
047B3ZZ
047B446
047B44Z
047B456
047B45Z
047B466
047B46Z
047B476
047B47Z
047B4D6
047B4DZ
047B4E6
047B4EZ
047B4F6
047B4FZ
047B4G6
047B4GZ
047B4Z6
047B4ZZ
047C046
047C04Z
047C056

047C05Z
047C066
047C06Z
047C076
047C07Z
047C0D6
047C0DZ
047C0E6
047C0EZ
047C0F6
047C0FZ
047C0G6
047C0GZ
047C0Z6
047C0ZZ
047C346
047C34Z
047C356
047C35Z
047C366
047C36Z
047C376
047C37Z
047C3D6
047C3DZ
047C3E6
047C3EZ
047C3F6
047C3FZ
047C3G6
047C3GZ
047C3Z6
047C3ZZ
047C446
047C44Z
047C456
047C45Z
047C466
047C46Z
047C476
047C47Z
047C4D6
047C4DZ

047C4E6
047C4EZ
047C4F6
047C4FZ
047C4G6
047C4GZ
047C4Z6
047C4ZZ
047D046
047D04Z
047D056
047D05Z
047D066
047D06Z
047D076
047D07Z
047D0D6
047D0DZ
047D0E6
047D0EZ
047D0F6
047D0FZ
047D0G6
047D0GZ
047D0Z6
047D0ZZ
047D346
047D34Z
047D356
047D35Z
047D366
047D36Z
047D376
047D37Z
047D3D6
047D3DZ
047D3E6
047D3EZ
047D3F6
047D3FZ
047D3G6
047D3GZ
047D3Z6

047D3ZZ
047D446
047D44Z
047D456
047D45Z
047D466
047D46Z
047D476
047D47Z
047D4D6
047D4DZ
047D4E6
047D4EZ
047D4F6
047D4FZ
047D4G6
047D4GZ
047D4Z6
047D4ZZ
047E046
047E04Z
047E056
047E05Z
047E066
047E06Z
047E076
047E07Z
047E0D6
047E0DZ
047E0E6
047E0EZ
047E0F6
047E0FZ
047E0G6
047E0GZ
047E0Z6
047E0ZZ
047E346
047E34Z
047E356
047E35Z
047E366
047E36Z

047E376
047E37Z
047E3D6
047E3DZ
047E3E6
047E3EZ
047E3F6
047E3FZ
047E3G6
047E3GZ
047E3Z6
047E3ZZ
047E446
047E44Z
047E456
047E45Z
047E466
047E46Z
047E476
047E47Z
047E4D6
047E4DZ
047E4E6
047E4EZ
047E4F6
047E4FZ
047E4G6
047E4GZ
047E4Z6
047E4ZZ
047F046
047F04Z
047F056
047F05Z
047F066
047F06Z
047F076
047F07Z
047F0D6
047F0DZ
047F0E6
047F0EZ
047F0F6

047F0FZ
047F0G6
047F0GZ
047F0Z6
047F0ZZ
047F346
047F34Z
047F356
047F35Z
047F366
047F36Z
047F376
047F37Z
047F3D6
047F3DZ
047F3E6
047F3EZ
047F3F6
047F3FZ
047F3G6
047F3GZ
047F3Z6
047F3ZZ
047F446
047F44Z
047F456
047F45Z
047F466
047F46Z
047F476
047F47Z
047F4D6
047F4DZ
047F4E6
047F4EZ
047F4F6
047F4FZ
047F4G6
047F4GZ
047F4Z6
047F4ZZ
047H046
047H04Z

047H056
047H05Z
047H066
047H06Z
047H076
047H07Z
047H0D6
047H0DZ
047H0E6
047H0EZ
047H0F6
047H0FZ
047H0G6
047H0GZ
047H0Z6
047H0ZZ
047H346
047H34Z
047H356
047H35Z
047H366
047H36Z
047H376
047H37Z
047H3D6
047H3DZ
047H3E6
047H3EZ
047H3F6
047H3FZ
047H3G6
047H3GZ
047H3Z6
047H3ZZ
047H446
047H44Z
047H456
047H45Z
047H466
047H46Z
047H476
047H47Z
047H4D6

047H4DZ
047H4E6
047H4EZ
047H4F6
047H4FZ
047H4G6
047H4GZ
047H4Z6
047H4ZZ
047J046
047J04Z
047J056
047J05Z
047J066
047J06Z
047J076
047J07Z
047J0D6
047J0DZ
047J0E6
047J0EZ
047J0F6
047J0FZ
047J0G6
047J0GZ
047J0Z6
047J0ZZ
047J346
047J34Z
047J356
047J35Z
047J366
047J36Z
047J376
047J37Z
047J3D6
047J3DZ
047J3E6
047J3EZ
047J3F6
047J3FZ
047J3G6
047J3GZ

047J3Z6
047J3ZZ
047J446
047J44Z
047J456
047J45Z
047J466
047J46Z
047J476
047J47Z
047J4D6
047J4DZ
047J4E6
047J4EZ
047J4F6
047J4FZ
047J4G6
047J4GZ
047J4Z6
047J4ZZ
047K041
047K046
047K04Z
047K056
047K05Z
047K066
047K06Z
047K076
047K07Z
047K0D1
047K0D6
047K0DZ
047K0E6
047K0EZ
047K0F6
047K0FZ
047K0G6
047K0GZ
047K0Z1
047K0Z6
047K0ZZ
047K341
047K346

047K34Z
047K356
047K35Z
047K366
047K36Z
047K376
047K37Z
047K3D1
047K3D6
047K3DZ
047K3E6
047K3EZ
047K3F6
047K3FZ
047K3G6
047K3GZ
047K3Z1
047K3Z6
047K3ZZ
047K441
047K446
047K44Z
047K456
047K45Z
047K466
047K46Z
047K476
047K47Z
047K4D1
047K4D6
047K4DZ
047K4E6
047K4EZ
047K4F6
047K4FZ
047K4G6
047K4GZ
047K4Z1
047K4Z6
047K4ZZ
047L041
047L046
047L04Z

047L056
047L05Z
047L066
047L06Z
047L076
047L07Z
047L0D1
047L0D6
047L0DZ
047L0E6
047L0EZ
047L0F6
047L0FZ
047L0G6
047L0GZ
047L0Z1
047L0Z6
047L0ZZ
047L341
047L346
047L34Z
047L356
047L35Z
047L366
047L36Z
047L376
047L37Z
047L3D1
047L3D6
047L3DZ
047L3E6
047L3EZ
047L3F6
047L3FZ
047L3G6
047L3GZ
047L3Z1
047L3Z6
047L3ZZ
047L441
047L446
047L44Z
047L456

047L45Z
047L466
047L46Z
047L476
047L47Z
047L4D1
047L4D6
047L4DZ
047L4E6
047L4EZ
047L4F6
047L4FZ
047L4G6
047L4GZ
047L4Z1
047L4Z6
047L4ZZ
047M041
047M046
047M04Z
047M056
047M05Z
047M066
047M06Z
047M076
047M07Z
047M0D1
047M0D6
047M0DZ
047M0E6
047M0EZ
047M0F6
047M0FZ
047M0G6
047M0GZ
047M0Z1
047M0Z6
047M0ZZ
047M341
047M346
047M34Z
047M356
047M35Z

047M366
047M36Z
047M376
047M37Z
047M3D1
047M3D6
047M3DZ
047M3E6
047M3EZ
047M3F6
047M3FZ
047M3G6
047M3GZ
047M3Z1
047M3Z6
047M3ZZ
047M441
047M446
047M44Z
047M456
047M45Z
047M466
047M46Z
047M476
047M47Z
047M4D1
047M4D6
047M4DZ
047M4E6
047M4EZ
047M4F6
047M4FZ
047M4G6
047M4GZ
047M4Z1
047M4Z6
047M4ZZ
047N041
047N046
047N04Z
047N056
047N05Z
047N066

047N06Z
047N076
047N07Z
047N0D1
047N0D6
047N0DZ
047N0E6
047N0EZ
047N0F6
047N0FZ
047N0G6
047N0GZ
047N0Z1
047N0Z6
047N0ZZ
047N341
047N346
047N34Z
047N356
047N35Z
047N366
047N36Z
047N376
047N37Z
047N3D1
047N3D6
047N3DZ
047N3E6
047N3EZ
047N3F6
047N3FZ
047N3G6
047N3GZ
047N3Z1
047N3Z6
047N3ZZ
047N441
047N446
047N44Z
047N456
047N45Z
047N466
047N46Z

047N476
047N47Z
047N4D1
047N4D6
047N4DZ
047N4E6
047N4EZ
047N4F6
047N4FZ
047N4G6
047N4GZ
047N4Z1
047N4Z6
047N4ZZ
047P046
047P04Z
047P056
047P05Z
047P066
047P06Z
047P076
047P07Z
047P0D6
047P0DZ
047P0E6
047P0EZ
047P0F6
047P0FZ
047P0G6
047P0GZ
047P0Z6
047P0ZZ
047P346
047P34Z
047P356
047P35Z
047P366
047P36Z
047P376
047P37Z
047P3D6
047P3DZ
047P3E6

047P3EZ
047P3F6
047P3FZ
047P3G6
047P3GZ
047P3Z6
047P3ZZ
047P446
047P44Z
047P456
047P45Z
047P466
047P46Z
047P476
047P47Z
047P4D6
047P4DZ
047P4E6
047P4EZ
047P4F6
047P4FZ
047P4G6
047P4GZ
047P4Z6
047P4ZZ
047Q046
047Q04Z
047Q056
047Q05Z
047Q066
047Q06Z
047Q076
047Q07Z
047Q0D6
047Q0DZ
047Q0E6
047Q0EZ
047Q0F6
047Q0FZ
047Q0G6
047Q0GZ
047Q0Z6
047Q0ZZ

047Q346
047Q34Z
047Q356
047Q35Z
047Q366
047Q36Z
047Q376
047Q37Z
047Q3D6
047Q3DZ
047Q3E6
047Q3EZ
047Q3F6
047Q3FZ
047Q3G6
047Q3GZ
047Q3Z6
047Q3ZZ
047Q446
047Q44Z
047Q456
047Q45Z
047Q466
047Q46Z
047Q476
047Q47Z
047Q4D6
047Q4DZ
047Q4E6
047Q4EZ
047Q4F6
047Q4FZ
047Q4G6
047Q4GZ
047Q4Z6
047Q4ZZ
047R046
047R04Z
047R056
047R05Z
047R066
047R06Z
047R076

047R07Z
047R0D6
047R0DZ
047R0E6
047R0EZ
047R0F6
047R0FZ
047R0G6
047R0GZ
047R0Z6
047R0ZZ
047R346
047R34Z
047R356
047R35Z
047R366
047R36Z
047R376
047R37Z
047R3D6
047R3DZ
047R3E6
047R3EZ
047R3F6
047R3FZ
047R3G6
047R3GZ
047R3Z6
047R3ZZ
047R446
047R44Z
047R456
047R45Z
047R466
047R46Z
047R476
047R47Z
047R4D6
047R4DZ
047R4E6
047R4EZ
047R4F6
047R4FZ

047R4G6
047R4GZ
047R4Z6
047R4ZZ
047S046
047S04Z
047S056
047S05Z
047S066
047S06Z
047S076
047S07Z
047S0D6
047S0DZ
047S0E6
047S0EZ
047S0F6
047S0FZ
047S0G6
047S0GZ
047S0Z6
047S0ZZ
047S346
047S34Z
047S356
047S35Z
047S366
047S36Z
047S376
047S37Z
047S3D6
047S3DZ
047S3E6
047S3EZ
047S3F6
047S3FZ
047S3G6
047S3GZ
047S3Z6
047S3ZZ
047S446
047S44Z
047S456

047S45Z
047S466
047S46Z
047S476
047S47Z
047S4D6
047S4DZ
047S4E6
047S4EZ
047S4F6
047S4FZ
047S4G6
047S4GZ
047S4Z6
047S4ZZ
047T046
047T04Z
047T056
047T05Z
047T066
047T06Z
047T076
047T07Z
047T0D6
047T0DZ
047T0E6
047T0EZ
047T0F6
047T0FZ
047T0G6
047T0GZ
047T0Z6
047T0ZZ
047T346
047T34Z
047T356
047T35Z
047T366
047T36Z
047T376
047T37Z
047T3D6
047T3DZ

047T3E6
047T3EZ
047T3F6
047T3FZ
047T3G6
047T3GZ
047T3Z6
047T3ZZ
047T446
047T44Z
047T456
047T45Z
047T466
047T46Z
047T476
047T47Z
047T4D6
047T4DZ
047T4E6
047T4EZ
047T4F6
047T4FZ
047T4G6
047T4GZ
047T4Z6
047T4ZZ
047U046
047U04Z
047U056
047U05Z
047U066
047U06Z
047U076
047U07Z
047U0D6
047U0DZ
047U0E6
047U0EZ
047U0F6
047U0FZ
047U0G6
047U0GZ
047U0Z6

047U0ZZ
047U346
047U34Z
047U356
047U35Z
047U366
047U36Z
047U376
047U37Z
047U3D6
047U3DZ
047U3E6
047U3EZ
047U3F6
047U3FZ
047U3G6
047U3GZ
047U3Z6
047U3ZZ
047U446
047U44Z
047U456
047U45Z
047U466
047U46Z
047U476
047U47Z
047U4D6
047U4DZ
047U4E6
047U4EZ
047U4F6
047U4FZ
047U4G6
047U4GZ
047U4Z6
047U4ZZ
047V046
047V04Z
047V056
047V05Z
047V066
047V06Z

047V076
047V07Z
047V0D6
047V0DZ
047V0E6
047V0EZ
047V0F6
047V0FZ
047V0G6
047V0GZ
047V0Z6
047V0ZZ
047V346
047V34Z
047V356
047V35Z
047V366
047V36Z
047V376
047V37Z
047V3D6
047V3DZ
047V3E6
047V3EZ
047V3F6
047V3FZ
047V3G6
047V3GZ
047V3Z6
047V3ZZ
047V446
047V44Z
047V456
047V45Z
047V466
047V46Z
047V476
047V47Z
047V4D6
047V4DZ
047V4E6
047V4EZ
047V4F6

047V4FZ
047V4G6
047V4GZ
047V4Z6
047V4ZZ
047W046
047W04Z
047W056
047W05Z
047W066
047W06Z
047W076
047W07Z
047W0D6
047W0DZ
047W0E6
047W0EZ
047W0F6
047W0FZ
047W0G6
047W0GZ
047W0Z6
047W0ZZ
047W346
047W34Z
047W356
047W35Z
047W366
047W36Z
047W376
047W37Z
047W3D6
047W3DZ
047W3E6
047W3EZ
047W3F6
047W3FZ
047W3G6
047W3GZ
047W3Z6
047W3ZZ
047W446
047W44Z

047W456
047W45Z
047W466
047W46Z
047W476
047W47Z
047W4D6
047W4DZ
047W4E6
047W4EZ
047W4F6
047W4FZ
047W4G6
047W4GZ
047W4Z6
047W4ZZ
047Y046
047Y04Z
047Y056
047Y05Z
047Y066
047Y06Z
047Y076
047Y07Z
047Y0D6
047Y0DZ
047Y0E6
047Y0EZ
047Y0F6
047Y0FZ
047Y0G6
047Y0GZ
047Y0Z6
047Y0ZZ
047Y346
047Y34Z
047Y356
047Y35Z
047Y366
047Y36Z
047Y376
047Y37Z
047Y3D6

047Y3DZ
047Y3E6
047Y3EZ
047Y3F6
047Y3FZ
047Y3G6
047Y3GZ
047Y3Z6
047Y3ZZ
047Y446
047Y44Z
047Y456
047Y45Z
047Y466
047Y46Z
047Y476
047Y47Z
047Y4D6
047Y4DZ
047Y4E6
047Y4EZ
047Y4F6
047Y4FZ
047Y4G6
047Y4GZ
047Y4Z6
047Y4ZZ
04C00Z6
04C00ZZ
04C03Z6
04C03ZZ
04C04Z6
04C04ZZ
04C10Z6
04C10ZZ
04C13Z6
04C13ZZ
04C14Z6
04C14ZZ
04C20Z6
04C20ZZ
04C23Z6
04C23ZZ

04C24Z6
04C24ZZ
04C30Z6
04C30ZZ
04C33Z6
04C33ZZ
04C34Z6
04C34ZZ
04C40Z6
04C40ZZ
04C43Z6
04C43ZZ
04C44Z6
04C44ZZ
04C50Z6
04C50ZZ
04C53Z6
04C53ZZ
04C54Z6
04C54ZZ
04C60Z6
04C60ZZ
04C63Z6
04C63ZZ
04C64Z6
04C64ZZ
04C70Z6
04C70ZZ
04C73Z6
04C73ZZ
04C74Z6
04C74ZZ
04C80Z6
04C80ZZ
04C83Z6
04C83ZZ
04C84Z6
04C84ZZ
04C90Z6
04C90ZZ
04C93Z6
04C93ZZ
04C94Z6

04C94ZZ
04CA0Z6
04CA0ZZ
04CA3Z6
04CA3ZZ
04CA4Z6
04CA4ZZ
04CB0Z6
04CB0ZZ
04CB3Z6
04CB3ZZ
04CB4Z6
04CB4ZZ
04CC0Z6
04CC0ZZ
04CC3Z6
04CC3ZZ
04CC4Z6
04CC4ZZ
04CD0Z6
04CD0ZZ
04CD3Z6
04CD3ZZ
04CD4Z6
04CD4ZZ
04CE0Z6
04CE0ZZ
04CE3Z6
04CE3ZZ
04CE4Z6
04CE4ZZ
04CF0Z6
04CF0ZZ
04CF3Z6
04CF3ZZ
04CF4Z6
04CF4ZZ
04CH0Z6
04CH0ZZ
04CH3Z6
04CH3ZZ
04CH4Z6
04CH4ZZ

04CJ0Z6
04CJ0ZZ
04CJ3Z6
04CJ3ZZ
04CJ4Z6
04CJ4ZZ
04CK0Z6
04CK0ZZ
04CK3Z6
04CK3ZZ
04CK4Z6
04CK4ZZ
04CL0Z6
04CL0ZZ
04CL3Z6
04CL3ZZ
04CL4Z6
04CL4ZZ
04CM0Z6
04CM0ZZ
04CM3Z6
04CM3ZZ
04CM4Z6
04CM4ZZ
04CN0Z6
04CN0ZZ
04CN3Z6
04CN3ZZ
04CN4Z6
04CN4ZZ
04CP0Z6
04CP0ZZ
04CP3Z6
04CP3ZZ
04CP4Z6
04CP4ZZ
04CQ0Z6
04CQ0ZZ
04CQ3Z6
04CQ3ZZ
04CQ4Z6
04CQ4ZZ
04CR0Z6

04CR0ZZ
04CR3Z6
04CR3ZZ
04CR4Z6
04CR4ZZ
04CS0Z6
04CS0ZZ
04CS3Z6
04CS3ZZ
04CS4Z6
04CS4ZZ
04CT0Z6
04CT0ZZ
04CT3Z6
04CT3ZZ
04CT4Z6
04CT4ZZ
04CU0Z6
04CU0ZZ
04CU3Z6
04CU3ZZ
04CU4Z6
04CU4ZZ
04CV0Z6
04CV0ZZ
04CV3Z6
04CV3ZZ
04CV4Z6
04CV4ZZ
04CW0Z6
04CW0ZZ
04CW3Z6
04CW3ZZ
04CW4Z6
04CW4ZZ
04CY0Z6
04CY0ZZ
04CY3Z6
04CY3ZZ
04CY4Z6
04CY4ZZ
04H00DZ
04H03DZ

04H04DZ
04H10DZ
04H13DZ
04H14DZ
04H20DZ
04H23DZ
04H24DZ
04H30DZ
04H33DZ
04H34DZ
04H40DZ
04H43DZ
04H44DZ
04H50DZ
04H53DZ
04H54DZ
04H60DZ
04H63DZ
04H64DZ
04H70DZ
04H73DZ
04H74DZ
04H80DZ
04H83DZ
04H84DZ
04H90DZ
04H93DZ
04H94DZ
04HA0DZ
04HA3DZ
04HA4DZ
04HB0DZ
04HB3DZ
04HB4DZ
04HC0DZ
04HC3DZ
04HC4DZ
04HD0DZ
04HD3DZ
04HD4DZ
04HE0DZ
04HE3DZ
04HE4DZ

04HF0DZ
04HF3DZ
04HF4DZ
04HH0DZ
04HH3DZ
04HH4DZ
04HJ0DZ
04HJ3DZ
04HJ4DZ
04HK0DZ
04HK3DZ
04HK4DZ
04HL0DZ
04HL3DZ
04HL4DZ
04HM0DZ
04HM3DZ
04HM4DZ
04HN0DZ
04HN3DZ
04HN4DZ
04HP0DZ
04HP3DZ
04HP4DZ
04HQ0DZ
04HQ3DZ
04HQ4DZ
04HR0DZ
04HR3DZ
04HR4DZ
04HS0DZ
04HS3DZ
04HS4DZ
04HT0DZ
04HT3DZ
04HT4DZ
04HU0DZ
04HU3DZ
04HU4DZ
04HV0DZ
04HV3DZ
04HV4DZ
04HW0DZ

04HW3DZ
04HW4DZ
04HY0DZ
04HY3DZ
04HY4DZ
04R007Z
04R00JZ
04R00KZ
04R047Z
04R04JZ
04R04KZ
04R107Z
04R10JZ
04R10KZ
04R147Z
04R14JZ
04R14KZ
04R207Z
04R20JZ
04R20KZ
04R247Z
04R24JZ
04R24KZ
04R307Z
04R30JZ
04R30KZ
04R347Z
04R34JZ
04R34KZ
04R407Z
04R40JZ
04R40KZ
04R447Z
04R44JZ
04R44KZ
04R507Z
04R50JZ
04R50KZ
04R547Z
04R54JZ
04R54KZ
04R607Z
04R60JZ

04R60KZ
04R647Z
04R64JZ
04R64KZ
04R707Z
04R70JZ
04R70KZ
04R747Z
04R74JZ
04R74KZ
04R807Z
04R80JZ
04R80KZ
04R847Z
04R84JZ
04R84KZ
04R907Z
04R90JZ
04R90KZ
04R947Z
04R94JZ
04R94KZ
04RA07Z
04RA0JZ
04RA0KZ
04RA47Z
04RA4JZ
04RA4KZ
04RB07Z
04RB0JZ
04RB0KZ
04RB47Z
04RB4JZ
04RB4KZ
04RC07Z
04RC0JZ
04RC0KZ
04RC47Z
04RC4JZ
04RC4KZ
04RD07Z
04RD0JZ
04RD0KZ

04RD47Z
04RD4JZ
04RD4KZ
04RE07Z
04RE0JZ
04RE0KZ
04RE47Z
04RE4JZ
04RE4KZ
04RF07Z
04RF0JZ
04RF0KZ
04RF47Z
04RF4JZ
04RF4KZ
04RH07Z
04RH0JZ
04RH0KZ
04RH47Z
04RH4JZ
04RH4KZ
04RJ07Z
04RJ0JZ
04RJ0KZ
04RJ47Z
04RJ4JZ
04RJ4KZ
04RK07Z
04RK0JZ
04RK0KZ
04RK47Z
04RK4JZ
04RK4KZ
04RL07Z
04RL0JZ
04RL0KZ
04RL47Z
04RL4JZ
04RL4KZ
04RM07Z
04RM0JZ
04RM0KZ
04RM47Z

04RM4JZ
04RM4KZ
04RN07Z
04RN0JZ
04RN0KZ
04RN47Z
04RN4JZ
04RN4KZ
04RP07Z
04RP0JZ
04RP0KZ
04RP47Z
04RP4JZ
04RP4KZ
04RQ07Z
04RQ0JZ
04RQ0KZ
04RQ47Z
04RQ4JZ
04RQ4KZ
04RR07Z
04RR0JZ
04RR0KZ
04RR47Z
04RR4JZ
04RR4KZ
04RS07Z
04RS0JZ
04RS0KZ
04RS47Z
04RS4JZ
04RS4KZ
04RT07Z
04RT0JZ
04RT0KZ
04RT47Z
04RT4JZ
04RT4KZ
04RU07Z
04RU0JZ
04RU0KZ
04RU47Z
04RU4JZ

04RU4KZ
04RV07Z
04RV0JZ
04RV0KZ
04RV47Z
04RV4JZ
04RV4KZ
04RW07Z
04RW0JZ
04RW0KZ
04RW47Z
04RW4JZ
04RW4KZ
04RY07Z
04RY0JZ
04RY0KZ
04RY47Z
04RY4JZ
04RY4KZ
04U007Z
04U00JZ
04U00KZ
04U037Z
04U03JZ
04U03KZ
04U047Z
04U04JZ
04U04KZ
04U107Z
04U10JZ
04U10KZ
04U137Z
04U13JZ
04U13KZ
04U147Z
04U14JZ
04U14KZ
04U207Z
04U20JZ
04U20KZ
04U237Z
04U23JZ
04U23KZ

04U247Z
04U24JZ
04U24KZ
04U307Z
04U30JZ
04U30KZ
04U337Z
04U33JZ
04U33KZ
04U347Z
04U34JZ
04U34KZ
04U407Z
04U40JZ
04U40KZ
04U437Z
04U43JZ
04U43KZ
04U447Z
04U44JZ
04U44KZ
04U507Z
04U50JZ
04U50KZ
04U537Z
04U53JZ
04U53KZ
04U547Z
04U54JZ
04U54KZ
04U607Z
04U60JZ
04U60KZ
04U637Z
04U63JZ
04U63KZ
04U647Z
04U64JZ
04U64KZ
04U707Z
04U70JZ
04U70KZ
04U737Z

04U73JZ
04U73KZ
04U747Z
04U74JZ
04U74KZ
04U807Z
04U80JZ
04U80KZ
04U837Z
04U83JZ
04U83KZ
04U847Z
04U84JZ
04U84KZ
04U907Z
04U90JZ
04U90KZ
04U937Z
04U93JZ
04U93KZ
04U947Z
04U94JZ
04U94KZ
04UA07Z
04UA0JZ
04UA0KZ
04UA37Z
04UA3JZ
04UA3KZ
04UA47Z
04UA4JZ
04UA4KZ
04UB07Z
04UB0JZ
04UB0KZ
04UB37Z
04UB3JZ
04UB3KZ
04UB47Z
04UB4JZ
04UB4KZ
04UC07Z
04UC0JZ

04UC0KZ
04UC37Z
04UC3JZ
04UC3KZ
04UC47Z
04UC4JZ
04UC4KZ
04UD07Z
04UD0JZ
04UD0KZ
04UD37Z
04UD3JZ
04UD3KZ
04UD47Z
04UD4JZ
04UD4KZ
04UE07Z
04UE0JZ
04UE0KZ
04UE37Z
04UE3JZ
04UE3KZ
04UE47Z
04UE4JZ
04UE4KZ
04UF07Z
04UF0JZ
04UF0KZ
04UF37Z
04UF3JZ
04UF3KZ
04UF47Z
04UF4JZ
04UF4KZ
04UH07Z
04UH0JZ
04UH0KZ
04UH37Z
04UH3JZ
04UH3KZ
04UH47Z
04UH4JZ
04UH4KZ

04UJ07Z
04UJ0JZ
04UJ0KZ
04UJ37Z
04UJ3JZ
04UJ3KZ
04UJ47Z
04UJ4JZ
04UJ4KZ
04UK07Z
04UK0JZ
04UK0KZ
04UK37Z
04UK3JZ
04UK3KZ
04UK47Z
04UK4JZ
04UK4KZ
04UL07Z
04UL0JZ
04UL0KZ
04UL37Z
04UL3JZ
04UL3KZ
04UL47Z
04UL4JZ
04UL4KZ
04UM07Z
04UM0JZ
04UM0KZ
04UM37Z
04UM3JZ
04UM3KZ
04UM47Z
04UM4JZ
04UM4KZ
04UN07Z
04UN0JZ
04UN0KZ
04UN37Z
04UN3JZ
04UN3KZ
04UN47Z

04UN4JZ
04UN4KZ
04UP07Z
04UP0JZ
04UP0KZ
04UP37Z
04UP3JZ
04UP3KZ
04UP47Z
04UP4JZ
04UP4KZ
04UQ07Z
04UQ0JZ
04UQ0KZ
04UQ37Z
04UQ3JZ
04UQ3KZ
04UQ47Z
04UQ4JZ
04UQ4KZ
04UR07Z
04UR0JZ
04UR0KZ
04UR37Z
04UR3JZ
04UR3KZ
04UR47Z
04UR4JZ
04UR4KZ
04US07Z
04US0JZ
04US0KZ
04US37Z
04US3JZ
04US3KZ
04US47Z
04US4JZ
04US4KZ
04UT07Z
04UT0JZ
04UT0KZ
04UT37Z
04UT3JZ

04UT3KZ
04UT47Z
04UT4JZ
04UT4KZ
04UU07Z
04UU0JZ
04UU0KZ
04UU37Z
04UU3JZ
04UU3KZ
04UU47Z
04UU4JZ
04UU4KZ
04UV07Z
04UV0JZ
04UV0KZ
04UV37Z
04UV3JZ
04UV3KZ
04UV47Z
04UV4JZ
04UV4KZ
04UW07Z
04UW0JZ
04UW0KZ
04UW37Z
04UW3JZ
04UW3KZ
04UW47Z
04UW4JZ
04UW4KZ
04UY07Z
04UY0JZ
04UY0KZ
04UY37Z
04UY3JZ
04UY3KZ
04UY47Z
04UY4JZ
04UY4KZ
04V00D6
04V00DJ
04V00DZ

04V00E6
04V00EZ
04V00F6
04V00FZ
04V03D6
04V03DJ
04V03DZ
04V03E6
04V03EZ
04V03F6
04V03FZ
04V04D6
04V04DJ
04V04DZ
04V04E6
04V04EZ
04V04F6
04V04FZ
