## Supplemental Results for "Gaps in lipid management after diabetes diagnosis and associated cardiovascular outcomes in a cohort of US adults"

**eTable 1:** Proportional LDL reduction from baseline at 1-year and end of follow up, stratified by statin use and ASCVD risk class.
**eTable 2:** Crude percentage of individuals started on statin therapy and achieving target LDL reduction within 5 years of initial diabetes diagnosis, stratified by year of diagnosis.
**eTable 3:** Baseline characteristics of those started on statin therapy within one year of diabetes diagnosis, with and without follow-up LDL value between 6 and 18 months post-diagnosis.
**eTable 4:** Distribution of individuals started on statin therapy within one year of diabetes diagnosis by ASCVD risk class and one-year LDL.
**eTable 5:** Crude number of events and adjusted hazard ratios for ASCVD diagnoses, stratified by one-year LDL values and ASCVD risk.
**eTable 6:** Sensitivity analyses with adjusted hazard ratios for ASCVD diagnoses, stratified by one-year LDL values and ASCVD risk, limited to those with baseline LDL values >100 mg/dL and >120 mg/dL to account for intraindividual variability.
**eTable 7:** Baseline characteristics of study population stratified by year of initial diabetes diagnosis.
**eFigure 1:** Crude cumulative incidence of initiation of statin therapy following initial diabetes diagnosis, excluding those initiated on therapy within initial 4 weeks, stratified by ASCVD risk category.
**eFigure 2:** Forest plot of HR for statin initiation and target LDL reduction within 5 years of initial diabetes, stratified by year of diagnosis, after covariate adjustment.
**eFigure 3a:** Crude cumulative incidence of initiation of statin therapy following initial diabetes diagnosis, stratified by sex.
**eFigure 3b:** Forest plot of HR for statin initiation within 5 years for women compared to men after covariate adjustment.
**eFigure 3c:** Crude cumulative incidence of time to target LDL reduction following initial diabetes diagnosis, stratified by sex.
**eFigure 3d:** Forest plot of HR for target LDL reduction within 5 years for women compared to men after covariate adjustment.
**eFigure 4a:** Crude cumulative incidence of initiation of statin therapy following initial diabetes diagnosis, stratified by race/ethnicity.
**eFigure 4b:** Forest plot of HR for statin initiation within 5 years for Black, Hispanic, and "other" race/ethnicity compared to white individuals after covariate adjustment.
**eFigure 4c:** Crude cumulative incidence of time to target LDL reduction following initial diabetes diagnosis, stratified by race/ethnicity.
**eFigure 4d:** Forest plot of HR for target LDL reduction within 5 years for Black, Hispanic, and "other" race/ethnicity compared to white individuals after covariate adjustment.

**eTable 1:** Proportional LDL reduction from baseline at 1-year and end of follow up, stratified by statin use and ASCVD risk class.

| | **Average Proportional Reduction At 1 Year (SD)** | **Average Proportional Reduction At End of Follow-Up (SD)** |
|---|---|---|
| **Low Risk** | 0.2% (0.33) | 2.9% (0.37) |
| *On Statin at 5 Years* | 2.6 (0.34) | 7.9% (0.38) |
| *No Statin at 5 Years* | -4.0% (0.31) | -5.0% (0.35) |
| **Intermediate Risk** | 2.5% (0.33) | 8.5% (0.37) |
| *On Statin at 5 Years* | 5.1% (0.33) | 14.2% (0.36) |
| *No Statin at 5 Years* | -2.0% (0.31) | -1.0% (0.37) |
| **High Risk** | 5.9% (0.33) | 13.5% (0.36) |
| *On Statin at 5 Years* | 9.4% (0.35) | 20.1% (0.36) |
| *No Statin at 5 Years* | -1.0% (0.29) | 1.6% (0.33) |

**eTable 2.** Crude percentage of individuals started on statin therapy and achieving target LDL reduction within 5 years of initial diabetes diagnosis, stratified by year of diagnosis.

| Year of Diagnosis | Statin Initiation within 5 Years | Achievement of LDL <100 mg/dL within 5 Years |
|---:|---:|---:|
| 2005 | 61.20% | 58.40% |
| 2006 | 60.80% | 59.40% |
| 2007 | 60.50% | 61.70% |
| 2008 | 58.10% | 60.10% |
| 2009 | 58.00% | 61.00% |
| 2010 | 56.70% | 57.70% |
| 2011 | 56.10% | 57.40% |
| 2012 | 57.10% | 54.40% |
| 2013 | 59.60% | 54.30% |
| 2014 | 61.90% | 54.40% |
| 2015 | 65.20% | 52.80% |
| 2016 | 55.40% | 44.50% |

**eTable 3.** Baseline characteristics of those started on statin therapy within one year of diabetes diagnosis, with and without follow-up LDL value between 6 and 18 months post-diagnosis.

| **Baseline characteristics** | **One-Year LDL** | **No One-Year LDL** |
|---|---|---|
| Total population | 30,462 | 6185 |
| Age, median (IQR) | 57.7 (49.9-63.6) | 58.3 (50.3-64.8) |
| Male sex, n (%) | 28,556 (93.7) | 5919 (95.7) |
| Race/ethnicity, n (%) | | |
| *White* | 17,478 (57.4) | 3456 (55.9) |
| *Black* | 8133 (26.7) | 1601 (25.9) |
| *Hispanic* | 2429 (8.0) | 506 (8.2) |
| *Other* | 2422 (8.0) | 622 (10.1) |
| Body mass index, median (IQR) | 33.0 (29.3-37.5) | 32.2 (28.5-36.7) |
| Total cholesterol, median (IQR) | 200 (177-226) | 200 (174-229) |
| *High-density lipoprotein* | 38 (32-45) | 38 (32-45) |
| *Low-density lipoprotein* | 123 (103-143) | 122 (100-143) |
| **Comorbidities, n (%)** | | |
| Hypertension | 21,574 (70.8) | 4313 (69.7) |
| Hyperlipidemia | 15,256 (50.1) | 2843 (46.0) |
| Atrial fibrillation | 555 (1.8) | 127 (2.1) |
| Obstructive sleep apnea | 5422 (17.9) | 959 (15.5) |
| Liver disease | 2544 (8.4) | 462 (7.5) |
| Chronic kidney disease | 3062 (10.1) | 780 (12.6) |
| Smoking status | | |
| *Current* | 10,108 (33.2) | 2223 (35.9) |
| *Former* | 12,471 (40.9) | 2434 (39.4) |
| **Medications, n (%)** | | |
| Aspirin | 1848 (6.1) | 351 (5.7) |
| Antihypertensive agent | 17,976 (59.0) | 3478 (56.2) |
| Other lipid-lowering medication | 2102 (6.9) | 306 (4.9) |
| **Psychiatric diagnoses, n (%)** | | |
| Anxiety/depression | 12,201 (40.1) | 2301 (37.2) |
| Schizophrenia | 1161 (3.8) | 208 (3.4) |
| Substance use | 3396 (11.1) | 736 (11.9) |
| **Aging-specific variables, n (%)** | | |
| Dementia | 305 (1.0) | 120 (1.9) |
| Alzheimer's disease | 39 (0.1) | 16 (0.3) |
| Arthritis | 11,727 (38.5) | 2129 (34.4) |
| Fatigue | 755 (2.5) | 149 (2.4) |
| Walking difficulty | 796 (2.6) | 137 (2.2) |
| **Predicted 10-year ASCVD risk, n (%)** | | |
| Low-risk (<7.5%) | 4216 (13.8) | 776 (12.5) |
| Intermediate-risk (7.5-20%) | 9691 (31.8) | 1739 (28.1) |
| High-risk (>20%) | 16,555 (54.3) | 3671 (59.4) |

**eTable 4.** Distribution of individuals started on statin therapy within one year of diabetes diagnosis by ASCVD risk class and one-year LDL.

| | | One-Year LDL Value (2 Cat) | | One-Year LDL Value (3 Cat) | | |
|---|---|---|---|---|---|---|
| | | **<100 mg/dL** | **>100 mg/dL** | **<70 mg/dL** | **70-100 mg/dL** | **>100 mg/dL** |
| **PCE Class** | **Low** | 1997 (47.4%) | 2219 (52.6%) | 615 (14.6%) | 1382 (32.8%) | 2219 (52.6%) |
| | **Intermediate** | 7954 (53.3%) | 4522 (46.7%) | 1737 (17.9%) | 3432 (35.4%) | 4522 (46.7%) |
| | **High** | 9389 (56.7%) | 7166 (43.3%) | 3461 (20.9%) | 5928 (35.8%) | 7166 (43.3%) |

**eTable 5.** Crude number of events and adjusted hazard ratios for ASCVD diagnoses, stratified by one-year LDL values and ASCVD risk.

| Risk Category | One-Year LDL | Crude # of Events (%) | Adjusted HR (95% CI) | p-value |
|---|---|---|---|---|
| Low Risk (<7.5%) (n=4216) | <100 mg/dL (n=1997) | 71 (3.6) | 0.70 (0.48-1.03) | 0.069 |
| | ≥100 mg/dL (n=2219) | 83 (3.7) | Reference | --- |
| Intermediate Risk (7.5-20%) (n=9691) | <100 mg/dL (n=5169) | 405 (7.8) | 0.81 (0.68-0.96) | 0.014 |
| | ≥100 mg/dL (n=4522) | 372 (8.2) | Reference | --- |
| High Risk (>20%) (n=16,555) | <100 mg/dL (n=9389) | 1093 (15.3) | 0.86 (0.77-0.96) | 0.006 |
| | ≥100 mg/dL (n=7166) | 900 (12.6) | Reference | --- |
| Low Risk (<7.5%) (n=4216) | <70 mg/dL (n=615) | 18 (2.9) | 0.59 (0.32-1.09) | 0.092 |
| | 70-100 mg/dL (n=1382) | 53 (3.8) | 0.75 (0.49-1.13) | 0.163 |
| | >100 mg/dL (n=2219) | 83 (3.7) | Reference | --- |
| Intermediate Risk (7.5-20%) (n=9691) | <70 mg/dL (n=1737) | 138 (7.9) | 0.75 (0.59-0.95) | 0.019 |
| | 70-100 mg/dL (n=3432) | 267 (7.8) | 0.83 (0.69-1.00) | 0.055 |
| | >100 mg/dL (n=4522) | 372 (8.2) | Reference | --- |
| High Risk (>20%) (n=16,555) | <70 mg/dL (n=3461) | 356 (10.3) | 0.71 (0.61-0.84) | <0.001 |
| | 70-100 mg/dL (n=5928) | 737 (12.4) | 0.93 (0.83-1.05) | 0.237 |
| | >100 mg/dL (n=7166) | 900 (12.6) | Reference | --- |

**eTable 6.** Sensitivity analyses with adjusted hazard ratios for ASCVD diagnoses, stratified by one-year LDL values and ASCVD risk, limited to those with baseline LDL values >100 mg/dL and >120 mg/dL to account for intraindividual variability.

| **Sensitivity Analysis 1: Limiting population to those with baseline LDL ≥100 mg/dL** | | | |
|---|---|---|---|
| | **LDL <70 mg/dL** | **LDL 70-100 mg/dL** | **LDL >100 mg/dL** |
| **All (N=23,903)** | 0.70 (0.60-0.82), p<0.001 | 0.88 (0.79-0.98), p=0.020 | Reference |
| **Low-Risk (N=3201)** | 0.51 (0.22-1.20), p=0.123 | 0.60 (0.36-1.00), p=0.049 | Reference |
| **Intermediate-Risk (N=7384)** | 0.71 (0.52-0.97), p=0.031 | 0.78 (0.63-0.98), p=0.030 | Reference |
| **High-Risk (N=13,318)** | 0.71 (0.59-0.85), p<0.001 | 0.94 (0.82-1.07), p=0.330 | Reference |
| **Sensitivity Analysis 2: Limiting population to those with baseline LDL ≥120 mg/dL** | | | |
| | **LDL <70 mg/dL** | **LDL 70-100 mg/dL** | **LDL >100 mg/dL** |
| **All (N=16,561)** | 0.71 (0.58-0.87), p<0.001 | 0.87 (0.77-0.99), p=0.039 | Reference |
| **Low-Risk (N=2175)** | 0.18 (0.02-1.32), p=0.092 | 0.72 (0.39-1.34), p=0.298 | Reference |
| **Intermediate-Risk (N=5019)** | 0.72 (0.46-1.11), p=0.137 | 0.82 (0.63-1.08), p=0.160 | Reference |
| **High-Risk (N=9367)** | 0.74 (0.59-0.93), p=0.009 | 0.89 (0.77-1.04), p=0.149 | Reference |

**eTable 7.** Baseline characteristics of study population stratified by year of initial diabetes diagnosis.

| Year of Diagnosis | 2005 | 2006 | 2007 | 2008 | 2009 | 2010 | 2011 | 2012 | 2013 | 2014 | 2015 | 2016 | Total |
|---|---|---|---|---|---|---|---|---|---|---|---|---|---|
| Total population | 9998 | 10,188 | 9652 | 9032 | 8981 | 8893 | 9096 | 8715 | 8845 | 8897 | 6451 | 1658 | 100,406 |
| Age, median (IQR) | 58.1 (51.9-67.0) | 58.5 (51.9-66.0) | 58.7 (51.9-64.6) | 58.8 (51.1-64.3) | 58.6 (50.7-63.7) | 58.8 (50.3-63.8) | 58.7 (50.1-64.4) | 58.4 (50.0-64.7) | 58.6 (49.6-65.2) | 58.0 (48.8-65.5) | 57.6 (48.0-66.0) | 63.1 (54.3-69.3) | 58.5 (50.6-65.0) |
| Male sex, n (%) | 9572 (95.7) | 9719 (95.4) | 9140 (94.7) | 8609 (95.3) | 8528 (95.0) | 8394 (94.4) | 8531 (93.8) | 8189 (94.0) | 8250 (93.3) | 8247 (92.7) | 5927 (91.9) | 1528 (92.2) | 94,634 (94.3) |
| Race/ethnicity, n (%) | | | | | | | | | | | | | |
| White | 5947 (59.5) | 6170 (60.6) | 5818 (60.3) | 5383 (59.6) | 5286 (58.9) | 5186 (58.3) | 5254 (57.8) | 4924 (56.5) | 5092 (57.6) | 5049 (56.7) | 3654 (56.6) | 980 (59.1) | 58,743 (58.5) |
| Black | 2153 (21.5) | 2202 (21.6) | 2212 (22.9) | 2169 (24.0) | 2158 (24.0) | 2272 (25.5) | 2410 (26.5) | 2410 (27.7) | 2420 (27.4) | 2409 (27.1) | 1770 (27.4) | 404 (24.4) | 24,989 (24.9) |
| Hispanic | 707 (7.1) | 740 (7.3) | 705 (7.3) | 685 (7.6) | 740 (8.2) | 698 (7.8) | 712 (7.8) | 729 (8.4) | 678 (7.7) | 767 (8.6) | 586 (9.1) | 137 (8.3) | 7,884 (7.9) |
| Other | 1191 (11.9) | 1076 (10.6) | 917 (9.5) | 795 (8.8) | 797 (8.9) | 737 (8.3) | 720 (7.9) | 652 (7.5) | 655 (7.4) | 672 (7.6) | 441 (6.8) | 137 (8.3) | 8,790 (8.8) |
| Body mass index, median (IQR) | 31.7 (28.1-36.1) | 32.0 (28.2-36.4) | 32.2 (28.5-36.6) | 32.3 (28.6-36.9) | 32.4 (28.7-36.9) | 32.8 (28.9-37.3) | 32.7 (29.0-37.3) | 32.9 (29.0-37.5) | 33.1 (29.2-37.7) | 33.3 (29.4-37.8) | 33.3 (29.4-37.9) | 32.7 (28.9-37.5) | 32.6 (28.8-37.1) |
| Total cholesterol, median (IQR) | 186 (161-213) | 184 (160-211) | 183 (159-209) | 182 (157-209) | 181 (157-208) | 181 (156-208) | 180 (156-206) | 180 (156-207) | 181 (157-208) | 179 (155-205) | 180 (156-207) | 175 (150-200) | 182 (157-208) |
| High-density lipoprotein | 38 (32-45) | 38 (32-45) | 37 (31-44) | 36 (31-44) | 36 (30-43) | 37 (31-44) | 37 (31-44) | 38 (32-45) | 38 (32-45) | 39 (33-46) | 39 (33-46) | 39 (33-47) | 32 (37-45) |
| Low-density lipoprotein | 108 (86-131) | 108 (86-130) | 107 (85-128) | 105 (84-127) | 105 (83-128) | 106 (85-128) | 104 (83-126) | 104 (83-127) | 104 (82-126) | 103 (81-126) | 104 (82-127) | 99 (80-120) | 105 (84-128) |
| Comorbidities, n (%) | | | | | | | | | | | | | |
| Hypertension | 7313 (73.1) | 7426 (72.9) | 7126 (73.8) | 6641 (73.5) | 6573 (73.2) | 6382 (71.8) | 6470 (71.1) | 6179 (70.9) | 6133 (69.3) | 5948 (66.9) | 4123 (63.9) | 1156 (69.7) | 71,470 (71.2) |
| Hyperlipidemia | 3285 (32.9) | 3664 (36.0) | 3608 (37.4) | 3504 (38.8) | 3606 (40.2) | 3604 (40.5) | 3692 (40.6) | 3540 (40.6) | 3603 (40.7) | 3374 (37.9) | 2203 (34.1) | 661 (39.9) | 38,344 (38.2) |
| Atrial fibrillation | 267 (2.7) | 233 (2.3) | 249 (2.6) | 206 (2.3) | 219 (2.4) | 183 (2.1) | 207 (2.3) | 201 (2.3) | 217 (2.5) | 226 (2.5) | 140 (2.2) | 61 (3.7) | 2,409 (2.4) |
| Obstructive sleep apnea | 989 (9.9) | 1074 (10.5) | 1194 (12.4) | 1332 (14.7) | 1373 (15.3) | 1568 (17.6) | 1712 (18.8) | 1854 (21.3) | 2119 (24.0) | 2238 (25.2) | 1859 (28.8) | 419 (25.3) | 17,731 (17.7) |
| Liver disease | 1204 (12.0) | 1269 (12.5) | 1262 (13.1) | 1227 (13.6) | 1292 (14.4) | 1233 (13.9) | 1199 (13.2) | 1101 (12.6) | 1069 (12.1) | 1092 (12.3) | 781 (12.1) | 207 (12.5) | 12,936 (12.9) |
| Chronic kidney disease | 1590 (15.9) | 1491 (14.6) | 1458 (15.1) | 1107 (12.3) | 1035 (11.5) | 916 (10.3) | 889 (9.8) | 740 (8.5) | 830 (9.4) | 799 (9.0) | 535 (8.3) | 192 (11.6) | 11,582 (11.5) |
| Smoking status | | | | | | | | | | | | | |
| Current | 2758 (27.6) | 2947 (28.9) | 3045 (31.5) | 3070 (34.0) | 3106 (34.6) | 3108 (34.9) | 3196 (35.1) | 2985 (34.3) | 3063 (34.6) | 3005 (33.8) | 2118 (32.8) | 435 (26.2) | 32,836 (32.7) |
| Medications, n (%) | | | | | | | | | | | | | |
| Aspirin | 693 (6.9) | 638 (6.3) | 609 (6.3) | 551 (6.1) | 500 (5.6) | 541 (6.1) | 543 (6.0) | 505 (5.8) | 511 (5.8) | 498 (5.6) | 348 (5.4) | 121 (7.3) | 6,058 (6.0) |
| Antihypertensive agent | 6315 (63.2) | 6366 (62.5) | 6083 (63.0) | 5509 (61.0) | 5386 (60.0) | 5336 (60.0) | 5253 (57.8) | 5021 (57.6) | 4995 (56.5) | 4913 (55.2) | 3457 (53.6) | 897 (54.1) | 59,531 (59.3) |
| Other lipid-lowering medication | 852 (8.5) | 816 (8.0) | 768 (8.0) | 682 (7.6) | 716 (8.0) | 586 (6.6) | 606 (6.7) | 490 (5.6) | 468 (5.3) | 419 (4.7) | 265 (4.1) | 66 (4.0) | 6,734 (6.7) |
| Psychiatric diagnoses, n (%) | | | | | | | | | | | | | |
| Anxiety/depression | 3424 (34.2) | 3486 (34.2) | 3479 (36.0) | 3539 (39.2) | 3681 (41.0) | 3679 (41.4) | 3865 (42.5) | 3768 (43.2) | 3802 (43.0) | 3996 (44.9) | 2945 (45.7) | 702 (42.3) | 40,366 (40.2) |
| Schizophrenia | 447 (4.5) | 498 (4.9) | 440 (4.6) | 365 (4.0) | 376 (4.2) | 336 (3.8) | 333 (3.7) | 298 (3.4) | 284 (3.2) | 284 (3.2) | 215 (3.3) | 41 (2.5) | 3,917 (3.9) |
| Substance use | 943 (9.4) | 1015 (10.0) | 1042 (10.8) | 1118 (12.4) | 1200 (13.4) | 1187 (13.3) | 1218 (13.4) | 1116 (12.8) | 1152 (13.0) | 1140 (12.8) | 714 (11.1) | 166 (10.0) | 12,011 (12.0) |
| Aging-specific variables, n (%) | | | | | | | | | | | | | |
| Dementia | 168 (1.7) | 203 (2.0) | 152 (1.6) | 153 (1.7) | 146 (1.6) | 107 (1.2) | 135 (1.5) | 126 (1.4) | 103 (1.2) | 110 (1.2) | 80 (1.2) | 26 (1.6) | 1,509 (1.5) |
| Alzheimer’s disease | 25 (0.3) | 24 (0.2) | 21 (0.2) | 18 (0.2) | 15 (0.2) | 17 (0.2) | 15 (0.2) | 20 (0.2) | 10 (0.1) | 11 (0.1) | 5 (0.0) | 4 (0.2) | 185 (0.2) |
| Arthritis | 4100 (41.0) | 4249 (41.7) | 4013 (41.6) | 3807 (42.2) | 3621 (40.3) | 3502 (39.4) | 3534 (38.9) | 3304 (37.9) | 3419 (38.7) | 3401 (38.2) | 2395 (37.1) | 738 (44.5) | 40,083 (39.9) |
| Fatigue | 232 (2.3) | 228 (2.2) | 237 (2.5) | 228 (2.5) | 261 (2.9) | 265 (3.0) | 287 (3.2) | 258 (3.0) | 312 (3.5) | 308 (3.5) | 238 (3.7) | 67 (4.0) | 2,921 (2.9) |
| Walking difficulty | 252 (2.5) | 269 (2.6) | 259 (2.7) | 267 (3.0) | 286 (3.2) | 286 (3.2) | 298 (3.3) | 313 (3.6) | 293 (3.3) | 289 (3.2) | 208 (3.2) | 47 (2.8) | 3,067 (3.1) |
| Predicted 10-year ASCVD risk, n (%) | | | | | | | | | | | | | |
| Low-risk (<7.5%) | 1265 (12.7) | 1309 (12.8) | 1242 (12.9) | 1217 (13.5) | 1239 (13.8) | 1332 (15.0) | 1486 (16.3) | 1471 (16.9) | 1527 (17.3) | 1771 (19.9) | 1359 (21.1) | 202 (12.2) | 15,420 (15.4) |
| Intermediate-risk (7.5-20%) | 3202 (32.0) | 3357 (33.0) | 3197 (33.1) | 2899 (32.1) | 2982 (33.2) | 2827 (31.8) | 2832 (31.1) | 2807 (32.2) | 2705 (30.6) | 2706 (30.4) | 1943 (30.1) | 439 (26.5) | 31,896 (31.8) |
| High-risk (>20%) | 5531 (55.3) | 5522 (54.2) | 5213 (54.0) | 4916 (54.4) | 4760 (53.0) | 4734 (53.2) | 4778 (52.5) | 4437 (50.9) | 4613 (52.2) | 4420 (49.7) | 3149 (48.8) | 1017 (61.3) | 53,090 (52.9) |

**eFigure 1.** Crude cumulative incidence of initiation of statin therapy following initial diabetes diagnosis, excluding those initiated on therapy within initial 4 weeks, stratified by ASCVD risk category.

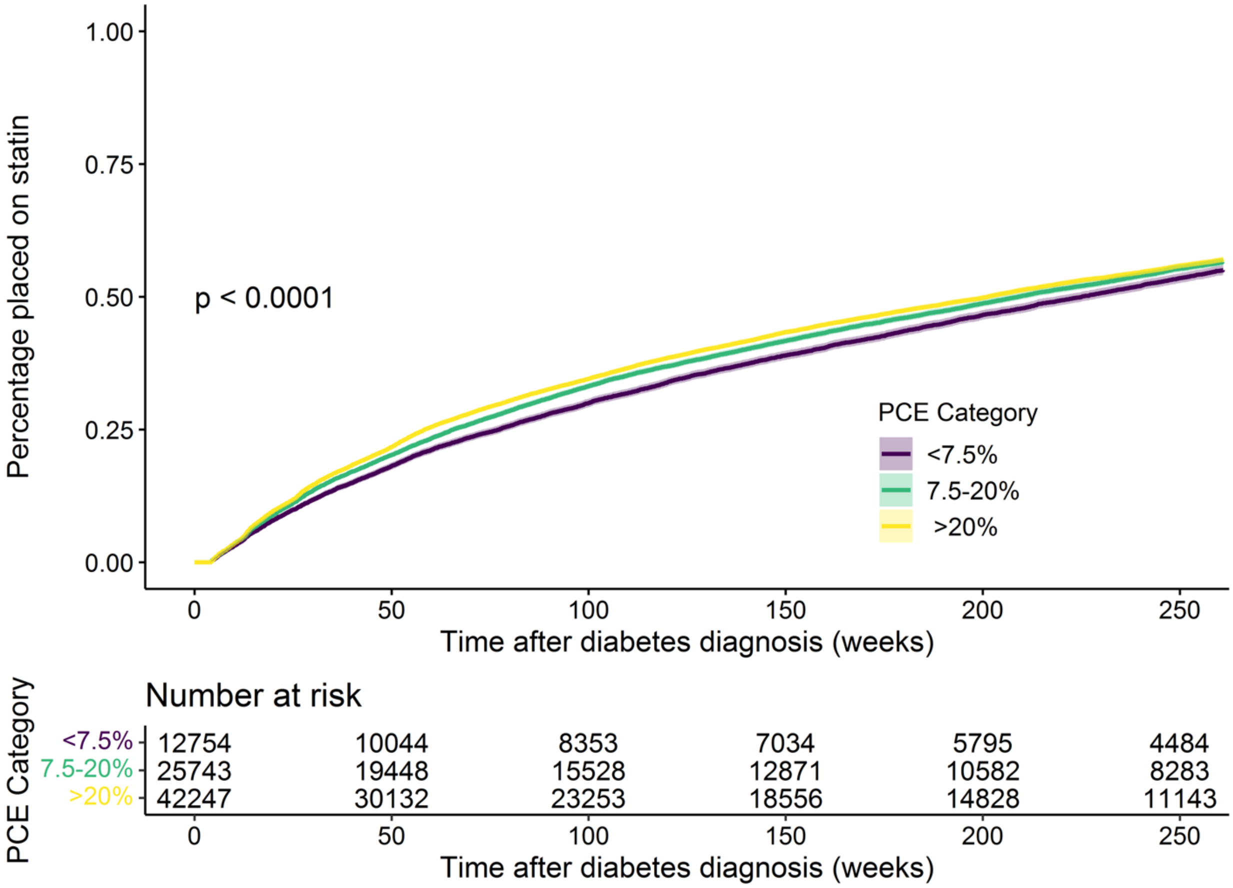

**eFigure 2.** Crude cumulative incidence of time to LDL reduction <70 mg/dL in those with baseline values above this level, following initial diabetes diagnosis, stratified by 10-year ASCVD risk as predicted by the ACC/AHA Pooled Cohorts Equation (PCE, low risk <7.5%, intermediate risk 7.5-20%, and high risk >20%).

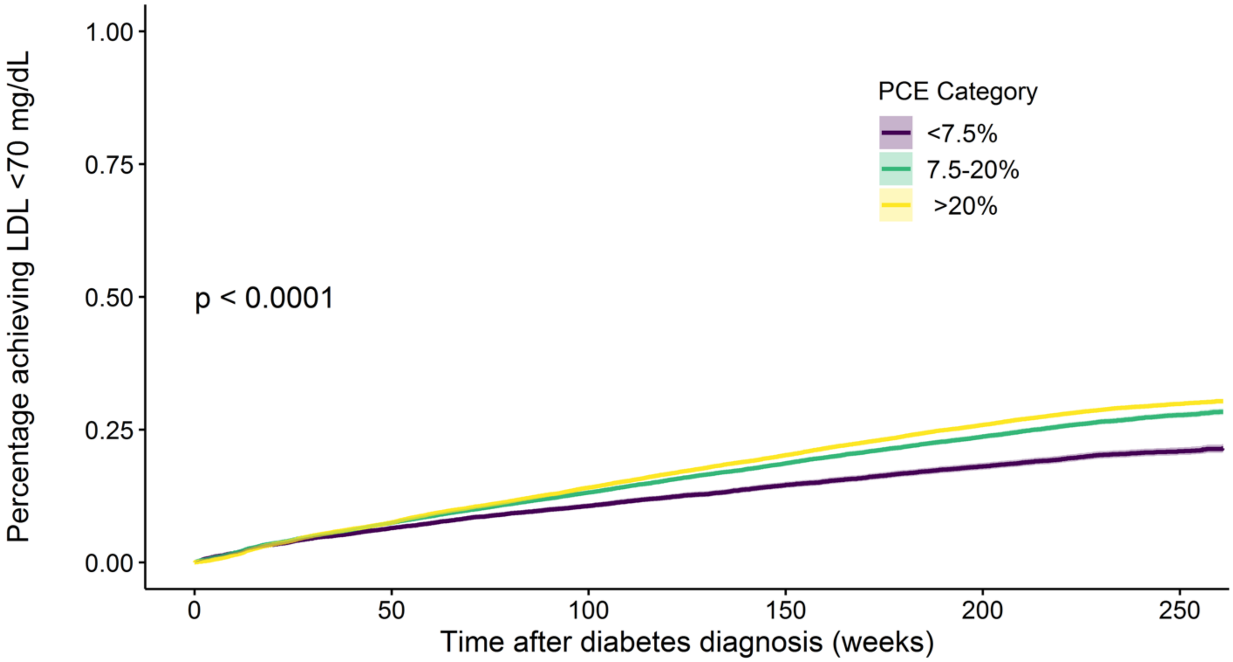

Number at risk

| PCE Category | 0 | 50 | 100 | 150 | 200 | 250 |
|---|---|---|---|---|---|---|
| <7.5% | 14746 | 13138 | 12148 | 11049 | 9134 | 2642 |
| 7.5-20% | 30761 | 27185 | 24669 | 21975 | 18056 | 5258 |
| >20% | 52260 | 45011 | 39599 | 34350 | 27623 | 7823 |

Time after diabetes diagnosis (weeks)

**eFigure 3.** Forest plot of HR for statin initiation and target LDL reduction within 5 years of initial diabetes, stratified by year of diagnosis, after covariate adjustment.

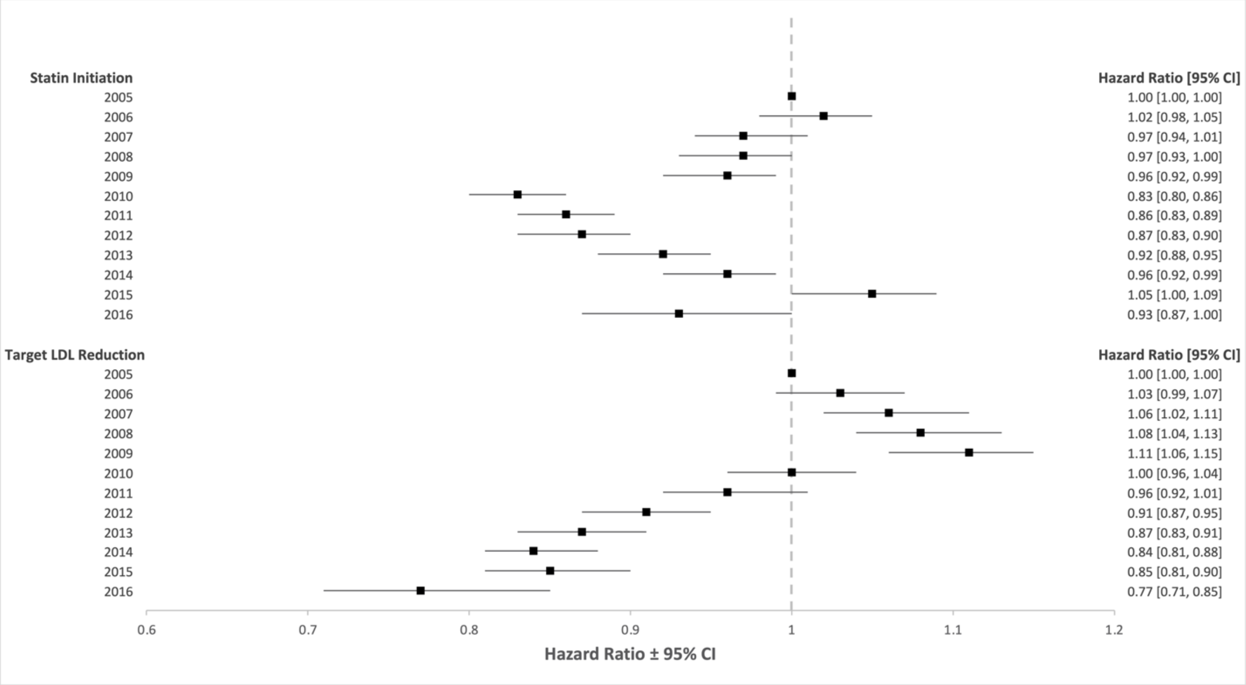

**eFigure 4a.** Crude cumulative incidence of initiation of statin therapy following initial diabetes diagnosis, stratified by sex.

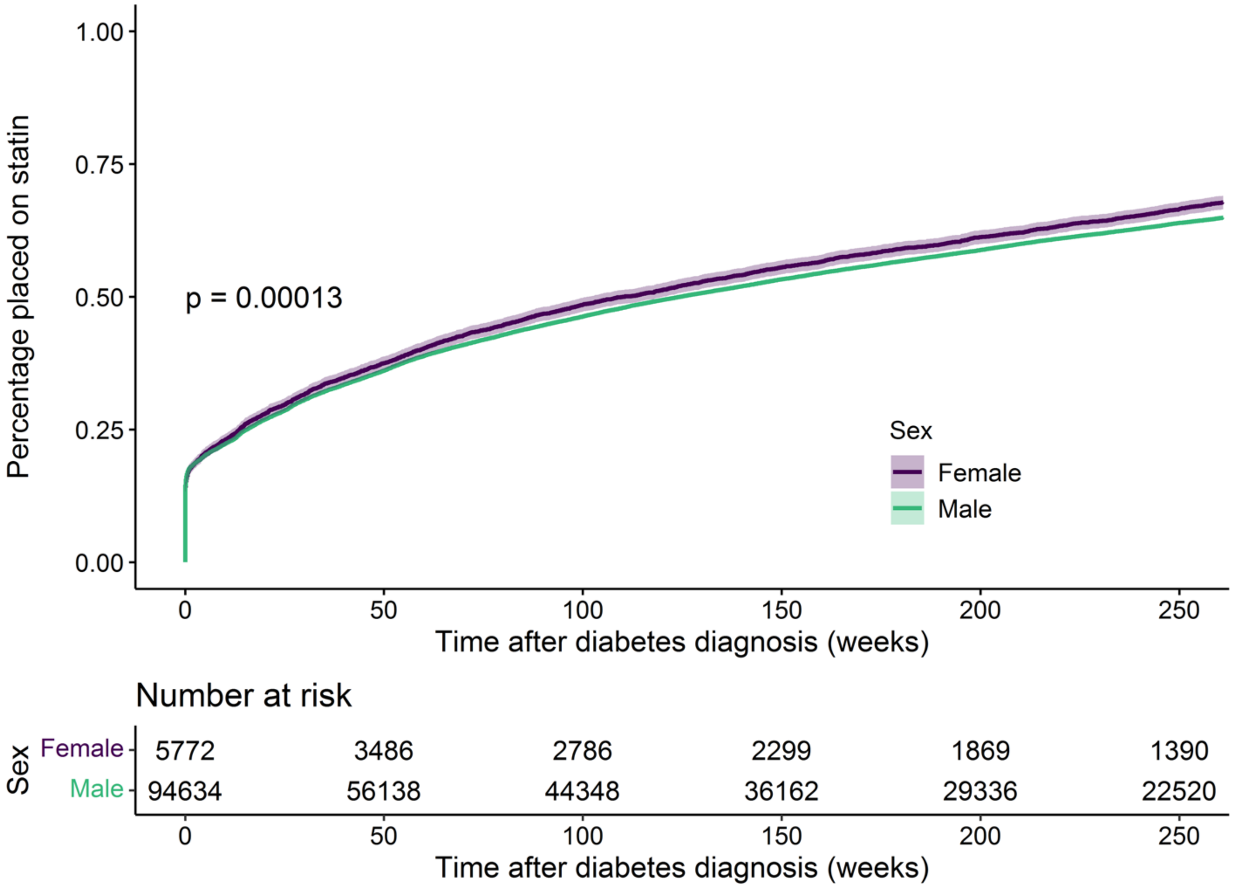

**eFigure 4b.** Forest plot of HR for statin initiation within 5 years for women compared to men after covariate adjustment.

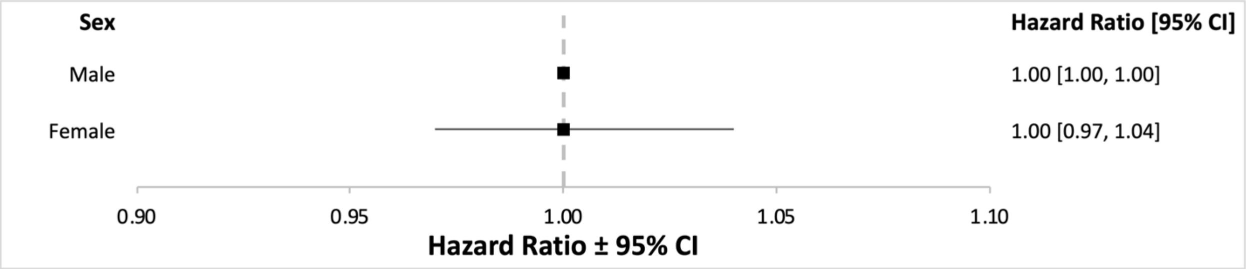

**eFigure 4c.** Crude cumulative incidence of time to target LDL reduction following initial diabetes diagnosis, stratified by sex.

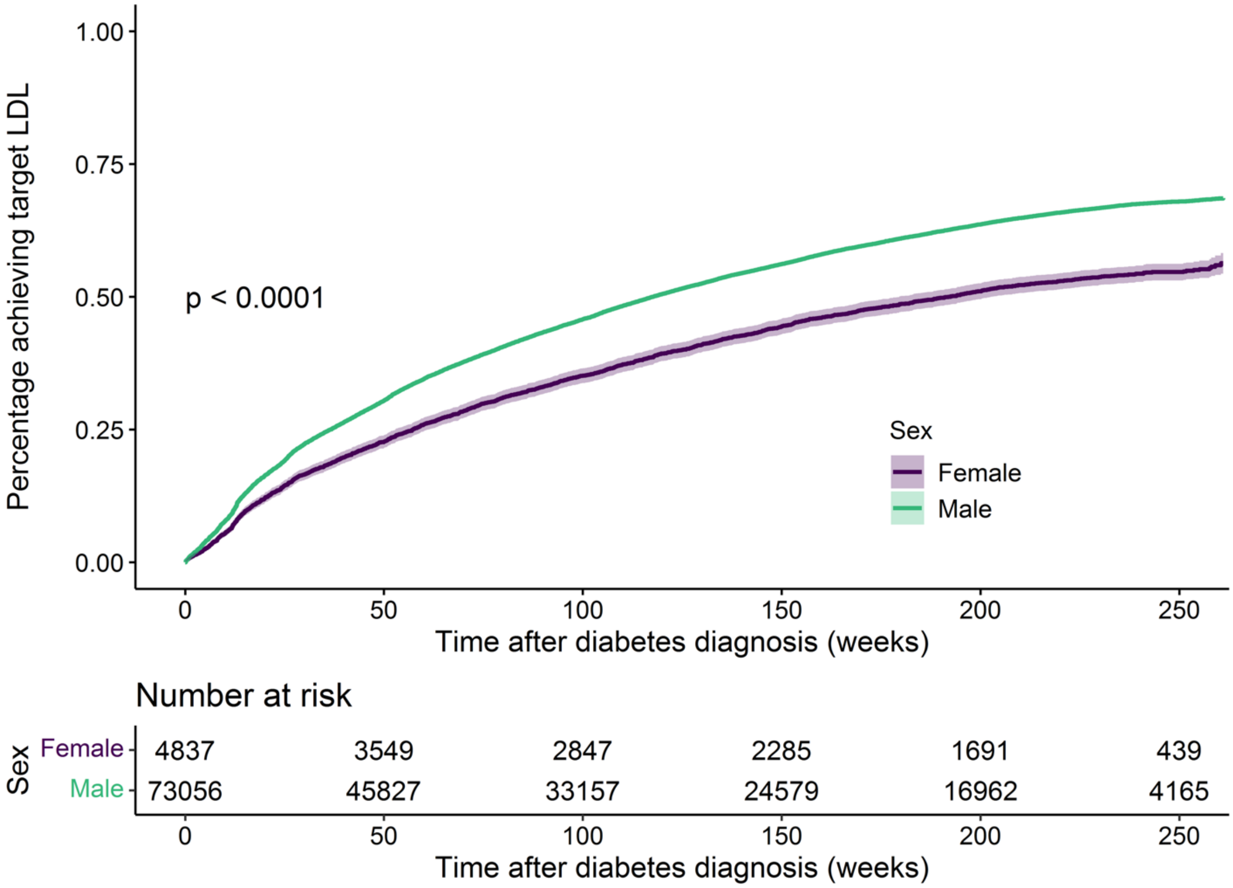

**eFigure 4d.** Forest plot of HR for target LDL reduction within 5 years for women compared to men after covariate adjustment.

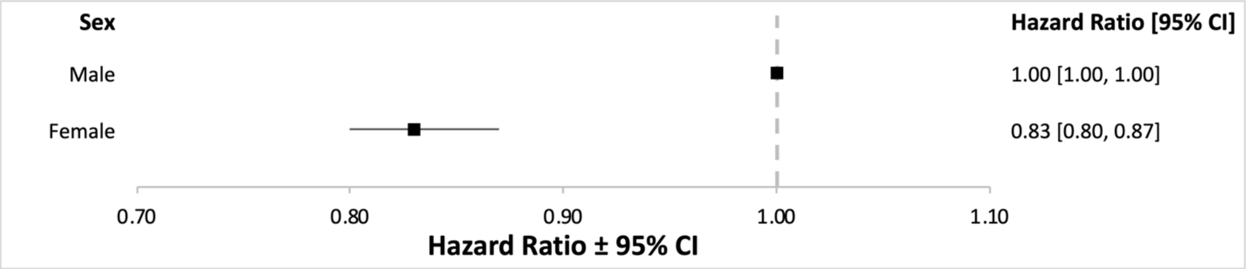

**eFigure 5a.** Crude cumulative incidence of initiation of statin therapy following initial diabetes diagnosis, stratified by race/ethnicity.

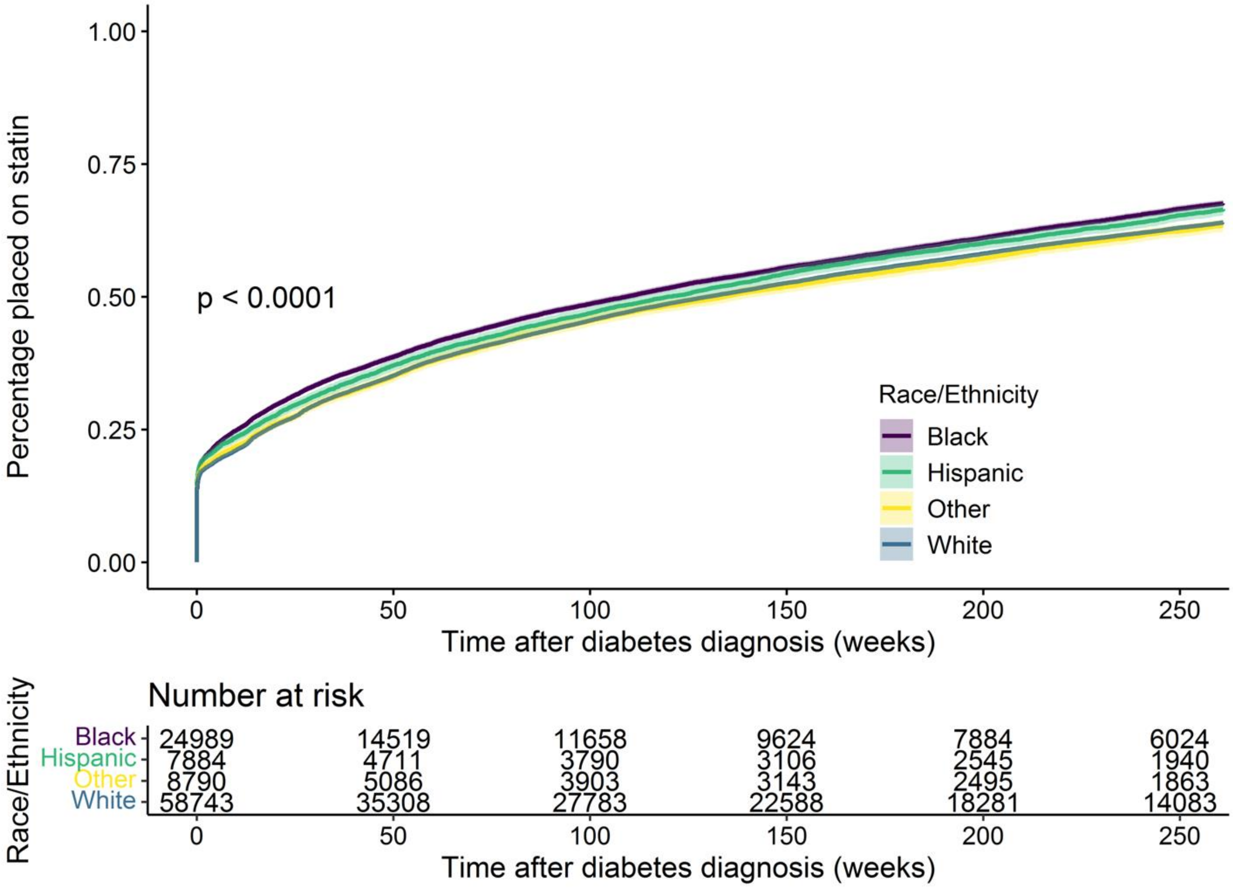

**eFigure 5b.** Forest plot of HR for statin initiation within 5 years for Black, Hispanic, and "other" race/ethnicity compared to white individuals after covariate adjustment.

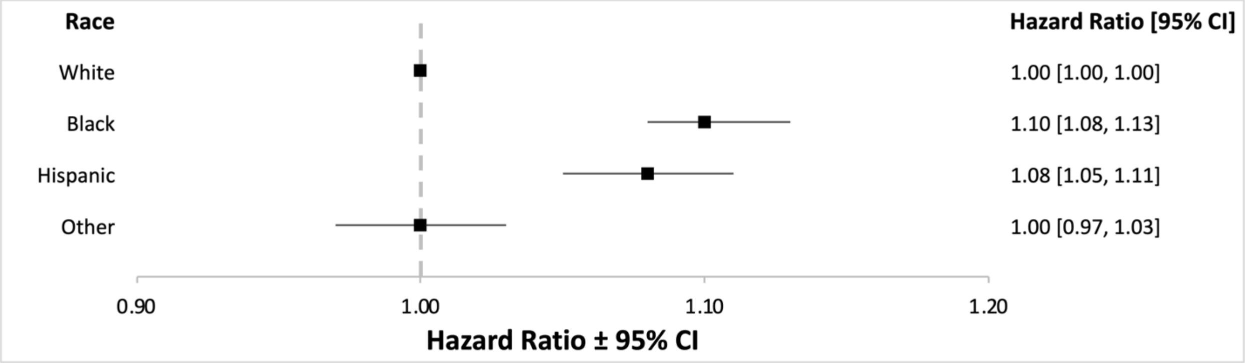

**eFigure 5c.** Crude cumulative incidence of time to target LDL reduction following initial diabetes diagnosis, stratified by race/ethnicity.

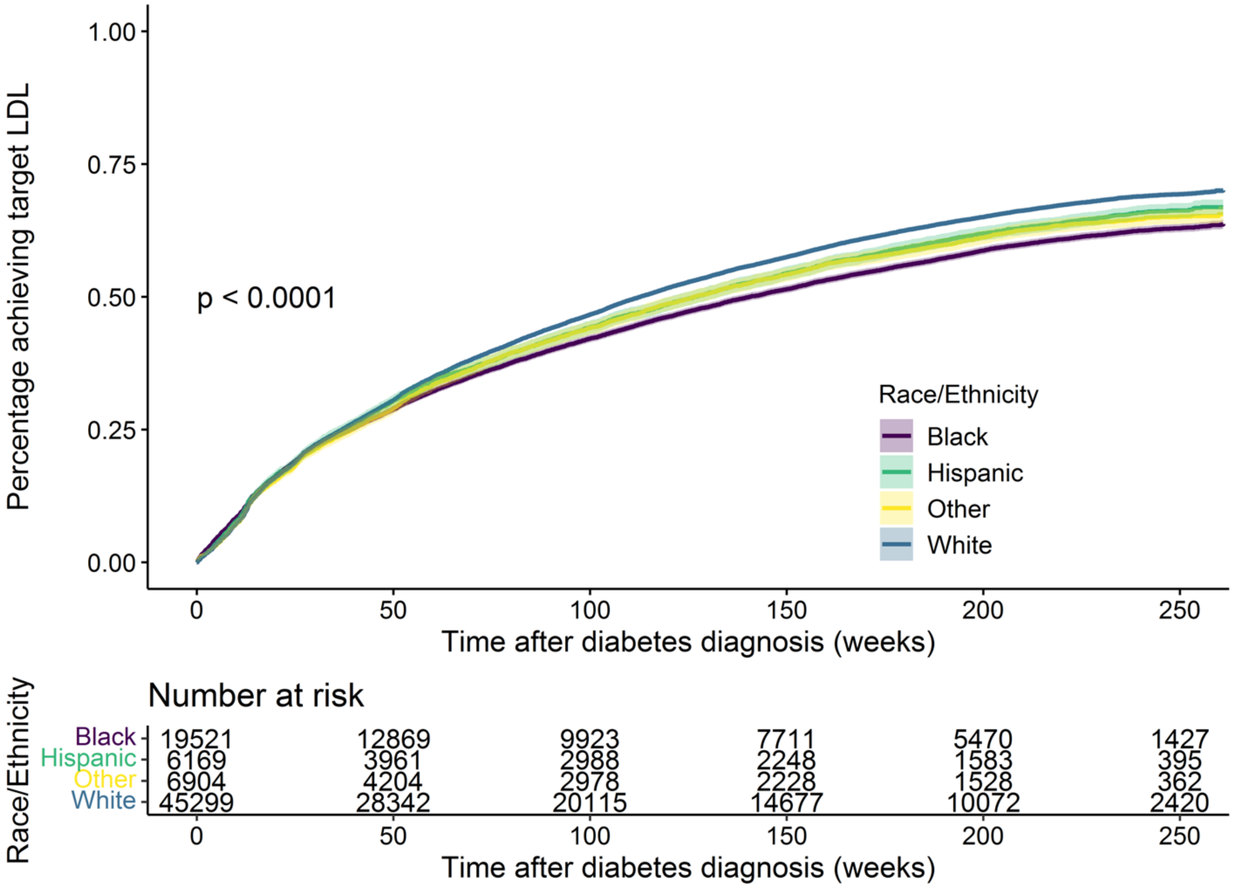

**eFigure 5d.** Forest plot of HR for target LDL reduction within 5 years for Black, Hispanic, and "other" race/ethnicity compared to white individuals after covariate adjustment.

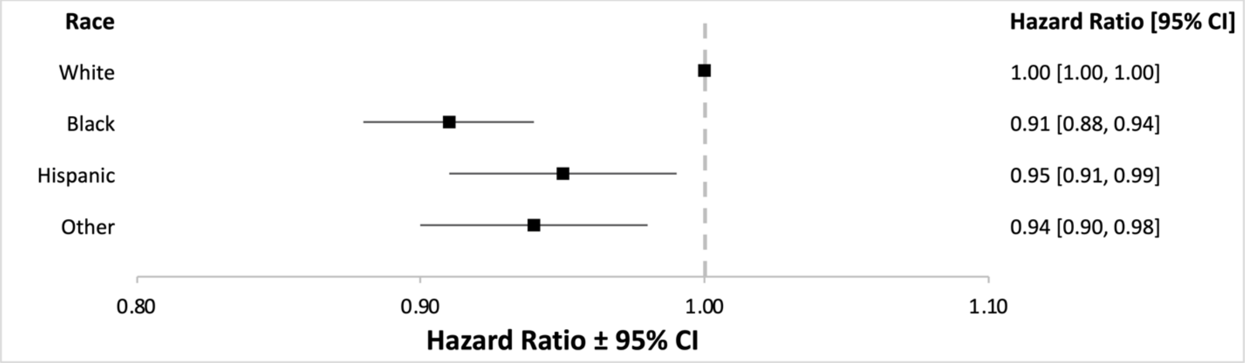
